# Epidemiological Association of Cannabinoid- and Drug- Exposures and Sociodemographic Factors with Limb Reduction Defects Across USA 1989-2016: A Geotemporospatial and Causal Inference Study

**DOI:** 10.1101/2020.09.01.20186163

**Authors:** Albert Stuart Reece, Gary Kenneth Hulse

**Affiliations:** Division of Psychiatry, University of Western Australia, Crawley, Western Australia 6009, Australia; School of Medical and Health Sciences, Edith Cowan University, Joondalup, Western Australia, 6027, Australia

**Keywords:** cannabis, cannabinoid, Δ9-tetrahydrocannabinol, cannabigerol, cannabis-related teratology, limb reduction defects

## Abstract

Reports of major limb defects after prenatal cannabis exposure (PCE) in animals and of human populations in Hawaii, Europe and Australia raise the question of whether the increasing use of cannabis in USA might be spatiotemporally associated with limb reduction rates (LRR) across USA. Geotemporospatial analysis conducted in R. LRR was significantly associated with cannabis use and THC potency and demonstrated prominent cannabis-use quintile effects. In final lagged geospatial models interactive terms including cannabinoids were highly significant and robust to adjustment. States in which cannabis was not legalized had a lower LRR (4.28 v 5.01 /10,000 live births, relative risk reduction = −0.15, (95%C.I. −0.25, −0.02), P=0.021). 37-63% of cases are estimated to not be born alive; their inclusion strengthened these associations. Causal inference studies using inverse probabilty-weighted robust regression and e-values supported causal epidemiological pathways. Findings apply to several cannabinoids, are consistent with pathophysiological and causal mechanisms, are exacerbated by cannabis legalization and demonstrate dose-related intergenerational sequaelae.

**Highlights:**

- Limb reduction rates (LRR) were associated with cannabis use, and THC potency
- These relationships were robust to adjustment for ethic and economic covariates
- They were maintained at geospatiotemporal regression
- LRR elevated as stillborn and aborted cases were considered
- Criteria of causality was fulfilled

## Introduction

Limb reductions (LR) are rare and dramatic defects which were first described several thousand years ago in the literature of antiquity (Bermejo-Sanchez, Cuevas et al. 2011, Bermejo-Sanchez, Cuevas et al. 2011). More recently they received prominence as the hallmark and initial indication of the teratogenic action of the drug thalidomide (Bermejo-Sanchez, Cuevas et al. 2011, Bermejo-Sanchez, Cuevas et al. 2011). LR includes both absence of proximal limb elements (intercalary segments, phocomelia) as well as complete limb absence limbs (amelia). LR occurs at a mean rate of 4.20/10,000. Given CDC data indicate 3,791,715 US births in 2018 this suggests over 1,600 LR cases annually (CDC, Centers for Disease Control et al. 2019). Leading studies from the International Clearing House of Birth Defects Surveillance and Research (ICBDSR) noted that little was known about the causes of these disorders (Bermejo-Sanchez, Cuevas et al. 2011, Bermejo-Sanchez, Cuevas et al. 2011). Sometimes LR arise as part of exceedingly rare congenital syndromes or together with multiple congenital anomalies, however their most common presentation is as an isolated disorder. ICBDSR noted that intrauterine vascular catastrophes have been shown to cause some cases with clear evidence of placental arteritis, vascular inflammation, subacute thrombosis and vascular and placental fibrosis seen in some cases (Hoyme, Jones et al. 1982, Bermejo-Sanchez, Cuevas et al. 2011, Bermejo-Sanchez, Cuevas et al. 2011). The upper limbs are known to be affected about twice as often as the lower. On the basis of minimal family inheritance isolated LR is not thought to have a genetic basis. Hotspots for both phocomelia and amelia have been reported, particularly in Victoria, Australia (Bermejo-Sanchez, Cuevas et al. 2011, Bermejo-Sanchez, Cuevas et al. 2011). It should be noted that not all cases are born alive. This is important as most registries list the numbers of cases as rates per 10,000 live births so that cases which occur as still births and cases for which early termination of pregnancy for the anomaly (ETOPFA) is performed can account for 37-63% of the total numbers as described below. This becomes an important issue analytically.

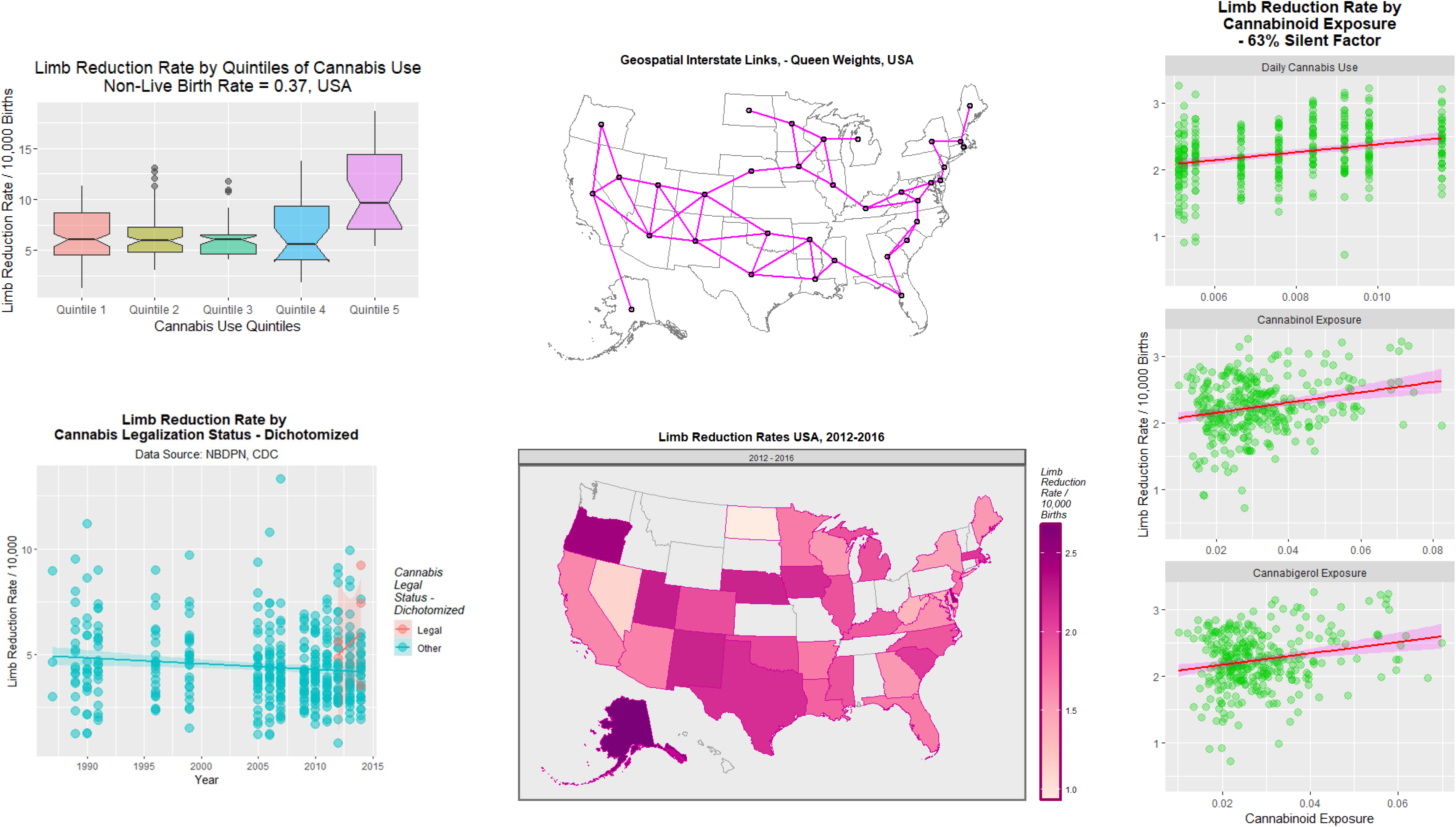

The embryology of limb development is complex and fascinating with limbs developing along each of the three spatial axes as a complex interplay of transcription factor and inducer gradients and an ordered and sequential cascade of molecular and signalling events. The emergence of the limb vessel which supplies the emerging structure is central to the maintenance of the whole process. It therefore becomes easy to understand how environmental impacts at critical stages could severely impact this sophisticated and coordinated sequential process.

A remarkably prescient paper from Hawaii found a greatly elevated rate of upper limb reduction deformity amongst patients exposed to cannabis either together with other drugs, or by itself (Forrester and Merz 2007). On the basis of 7 and 3 cases exposed amongst 115 total cases, rate ratios of 23.27 (95%C.I. 9.15-49.50) and 21.90 (4.45-65.63) were reported. Our hypothesis that rates of LR have a positive association with cannabis use was based on the previous Hawaiian findings and was formulated prior to the commencement of this work. Since both drug use and birth defect data was available for USA that nation formed our study setting.

## Methods

### Data

Birth defect data was taken from the annual reports of National Birth Defect Prevention Network organized by Centers for Disease Control Atlanta Georgia (National Birth Defects Prevention Network 2018). These reports are a collation of reports from state-based birth defects registries and normally report the dat in five year moving average style. The central year of this period was taken as the nominal year of the report. Data on the annual number of births in each state was taken from the CDC Wonder births registries (CDC, Centers for Disease Control et al. 2019). US Census data for populations, age distributions, ethnicity and median household income was accessed from US Census Bureau via tidycensus package from R. Drug use data was taken from the National Survey of Drug Use and Health (NSDUH) conducted annually by the Substance Abuse and Mental Health Services Administration (SAMHSA) (Substance Abuse and Mental Health Administration, Department of Health and Human Services et al. 2018). NSDUH is a survey which is carefully structured to be representative of the non-institutionalized US adult population. Cannabinoid (Δ9-tetrahydrocannabinol (Δ9THC), cannabigerol (CBG), cannabichromene, cannabinol, and cannabidiol) concentrations were taken from those reported in Federal Seizures by the Drug Enforcement Agency (ElSohly, Ross et al. 2000, Forrester and Merz 2007)

### Derived Variables

Quintiles were calculated based on the interval rather than the population distribution and were calculated with the cut_interval tool from ggplot2 in R. SAMHSA report different rates of intensity of cannabis use by days used last month by ethnicity at the national level. These data were used to calculate mean numbers of days cannabis was smoked. This figure was multiplied by the state cannabis use rate and the THC potency of cannabis in that year to derive a state based cannabis ethnic index referred to in the Tables as an ethnic “score.” The last month cannabis use rates, abbreviated to “mrjmon” in NSDUH, was multiplied by the cannabinoid concentration to derive an estimate of state-based levels of exposure to individual cannabinoids. The only longitudinal series of early termination for anomaly (ETOPFA) we were able to identify was the Western Australian Developmental Anomalies Registry (WARDA) (Women and Newborn Health Service, Department of Health et al. 2015). This series was used to calculate fractional maximal ETOPFA rates for each year to convert live birth anomaly rates to total rates.

### Statistics

Data was processed in “R-Studio” 1.2.1335 based on “R” 3.6.1 from CRAN. Variables were log transformed guided by the Shapiro test. Data were manipulated and matched in R-package dplyr, graphs were drawn in sf and ggplot2, geofacetting was done with geofacet, linear regression was performed in base, panel regression was performed in plm, two-step regression was performed in AER, spatial weights were prepared in spdep and spatial regression was performed in splm. For linear and panel regression missing data was casewise deleted. For spatial regression missing data was imputed by temporal kriging (mean substitution) mas described. LR datapoints more than 10 standard deviations from the mean were dropped. P<0.05 was considered significant.

Inverse probability weights (IPW) were calculated on the balanced kriged data using the ipw package in R. Causal inference was conducted using robust regression from the R-package survey and mixed effects regression from package nlme both using IPW weights. E-Values were used to estimated the strength of association an unknown confounder would require with both the predictor (mrjmon) and the outcome (Case rate) to explain away the effects, and were calculated using the package EValue in R.

### Data Sharing Statement

Key data including software code in R and a Data Dictionary key has been made available in the Mendeley data repository at this URL: http://dx.doi.org/10.17632/gtk7w24yvs.1.

### Ethics

This research was approved by the Human Ethics Research Committee of the University of Western Australia June 7^th^ 2019, (RA/4/20/4724).

## Results

437 data points relating to LR rate (LRR) were retrieved from NBDPN from 1986-1988 to 2012-2016 is shown in Supplementary Table 1. Prior to 2007-2011 LRR was listed separately for upper and lower limbs. After that time all limb defects were grouped under a single heading. Data from the earlier period were summed to make it comparable with the data from the later time period. For analytical purposes the middle year of each quoted time period is considered to be the nominal year of reference. A datapoint for Oklahoma 2005-2009 was omitted as it lay beyond 13.7 standard deviations outside the mean.

The median (+S.E.M.) LRR over the whole period of the NBDPN dataset was 4.20 (95%C.I. 2.51, 5.89) / 10,000 live births. The median figure for 2012-2016 was 4.10 (3.88, 4.31).

This data is also shown as a panel plot in Supplementary Figure 1, and as a geofacetted panel plot with each state in approximately its geographical location, in Supplementary Figure 2. The data is presented map graphically in Figure 1. Missing data points are evident.

**Figure 1.:**
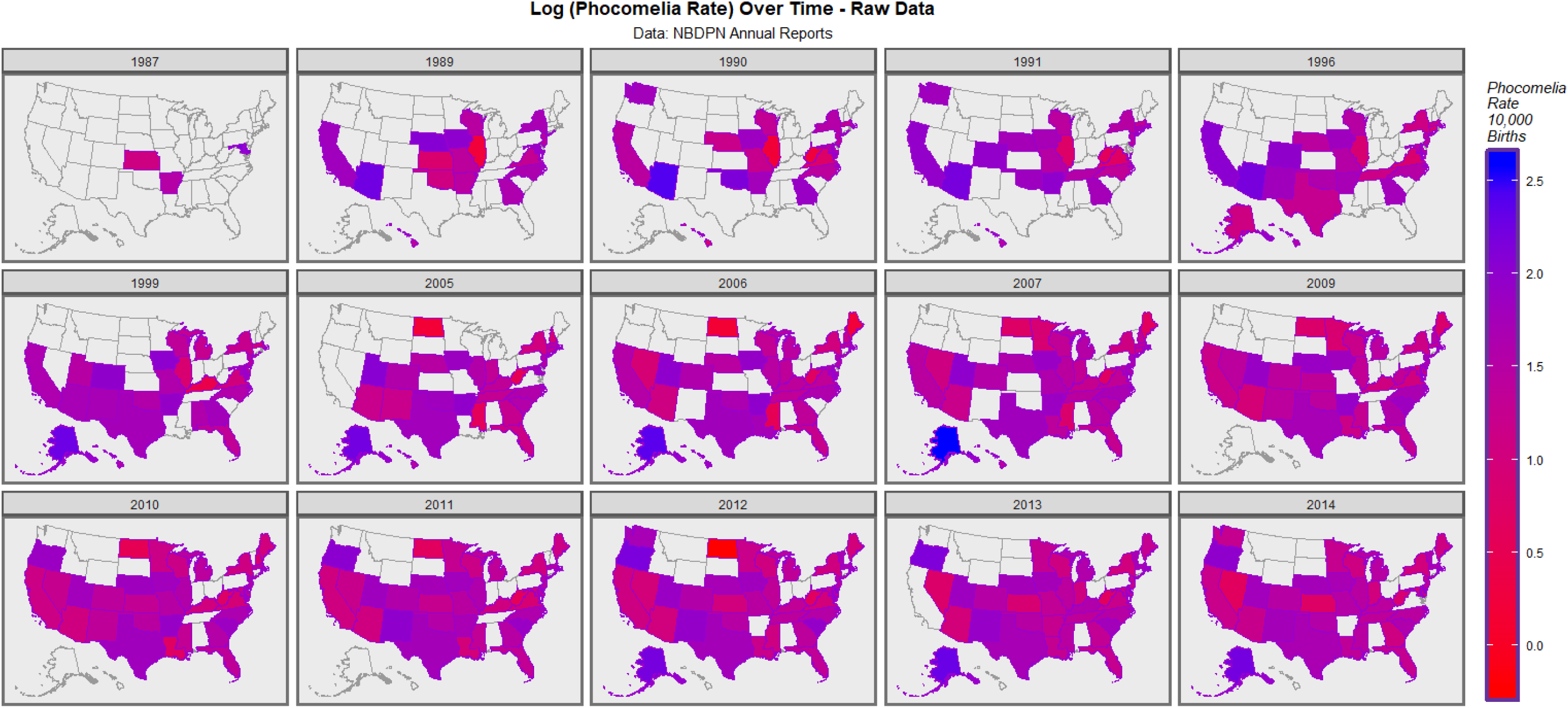
Map-graph of the limb reduction rate across USA over time. Raw data plot.

States were divided into quintiles based on their last month cannabis use rate in the 2017 NSDUH report as shown in Supplementary Table 2. Quintile 5 was made up of the states of Colorado, Alaska, Vermont, Puerto Rico and Quintile 4 of Maine, Rhode Island, Oregon, New Hampshire. The corresponding quintile median (+ S.E.M.) values for LRR are shown in Supplementary Table 3.

Figure 2 presents these data in an introductory manner. Panel A presents the LRR data over time. The time-trend appears essentially flat. Panel B presents it as a function of last month cannabis use. The trend appears to be rising. When these data are charted by quintile of cannabis use the highest quintile appears to be well above the others (Panel C). This is emphasized in Panel D where the highest quintile is charted alone and the lower four are grouped together. Panel E charts the LRR against cannabis exposure by cannabis use quintile. The highest quintile appears to be above the others. This is illustrated in Panel F where the highest quintile is shown compared to the remainder. Panel G presents a boxplot of LRR by quintile. Panel H presents a dichotomized plot of the highest quintile against the remainder.205

**Figure 2.:**
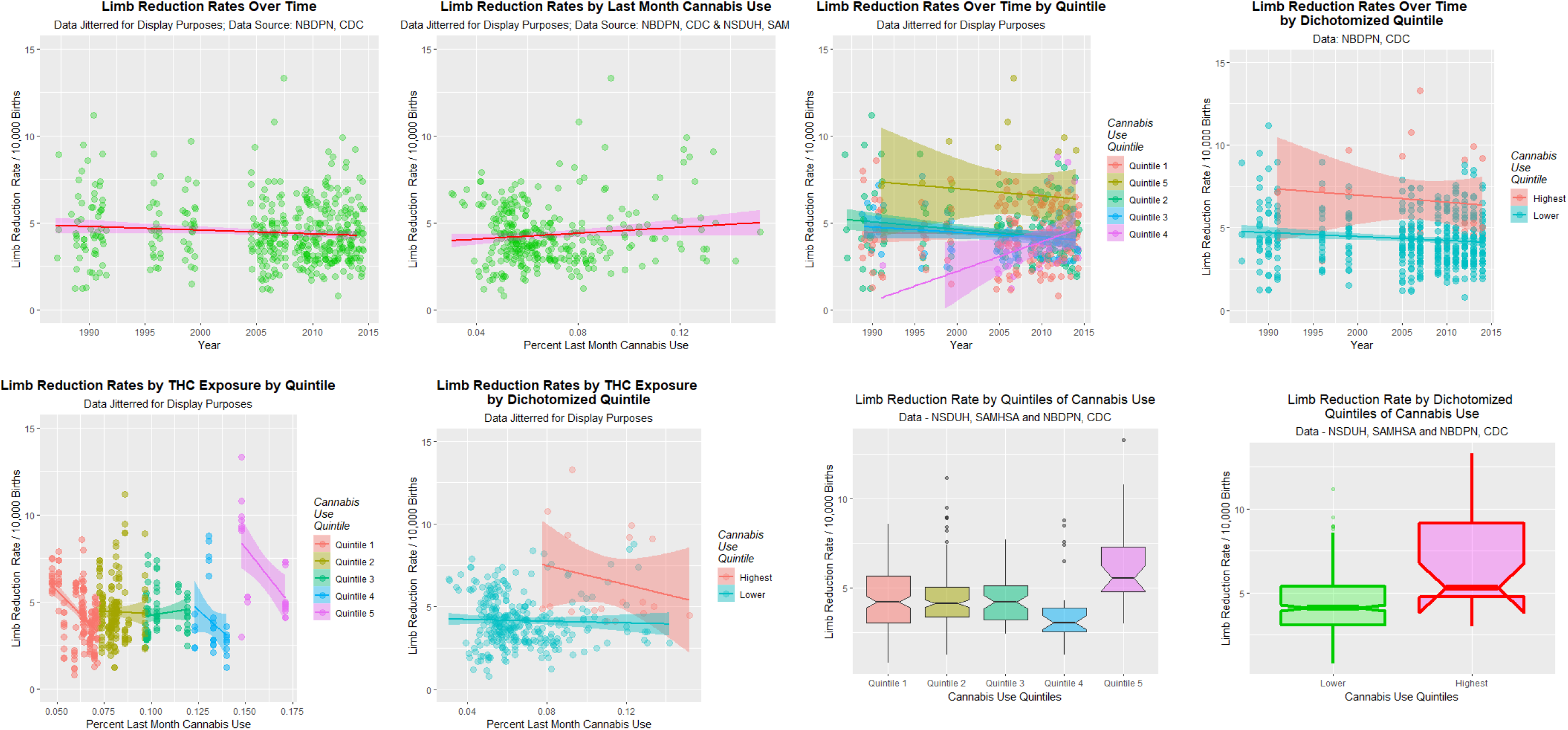
Univariate limb reduction trends. A: Trends over time. B. Trends by Cannabis use. C. Time trends by cannabis use quintile. D: Time trends by cannabis use dichotomized quintiles: highest quintile v. others. E: Limb reduction rates by cannabis use, by cannabis use quintile. F: Limb reduction rates by dichotomized cannabis use quintile by cannabis use. G: Boxplot of limb reduction rate by cannabis use quintile. Note jump from fourth to fifth quintile. H: Boxplot of limb reduction rate by dichotomized cannabis use quintiles, highest v. others.

The data are analyzed formally with results presented in Supplementary Table 4. The time trend is confirmed to not be significant, the quintile effect is highly significant (β-est.=0.434 (0.283, 0.585), P=2.9×10^-8^), the dichotomized quintile effect is highly significant (β-est.=0.416 (0.273, 0.559), P=2.2×10^-8^), the monthly cannabis use effect is significant (β-est.=8.507 (−0.040, 17.054), P=0.0492) and the THC exposure effect is also highly significant (from β-est.=10.637 (3.704, 17.569), P=0.0029).

Supplementary Figure 3 presents the mean national trends for daily or near daily use of cannabis (20-30 days per month) and for the numbers using cannabis whilst pregnant.

Figure 3 presents graphically data looking at the log LRR plotted against (A) drugs, (B) cannabinoids and (C) ethnicity. There is little relationship against drug use, with a suggestion of a declining effect with alcohol abuse or dependence shown in the first panel. Contrariwise with monthly cannabis use, Δ9THC exposure, cannabichromene, cannabinol and cannabigerol exposure there is a suggestion of a rising effect with exposure. The ethnicity plot shows a falling relationship with Caucasian-American ancestry, but a rising relationship with Hispanic-American and American Indian / Alaska Native identification.

**Figure 3.:**
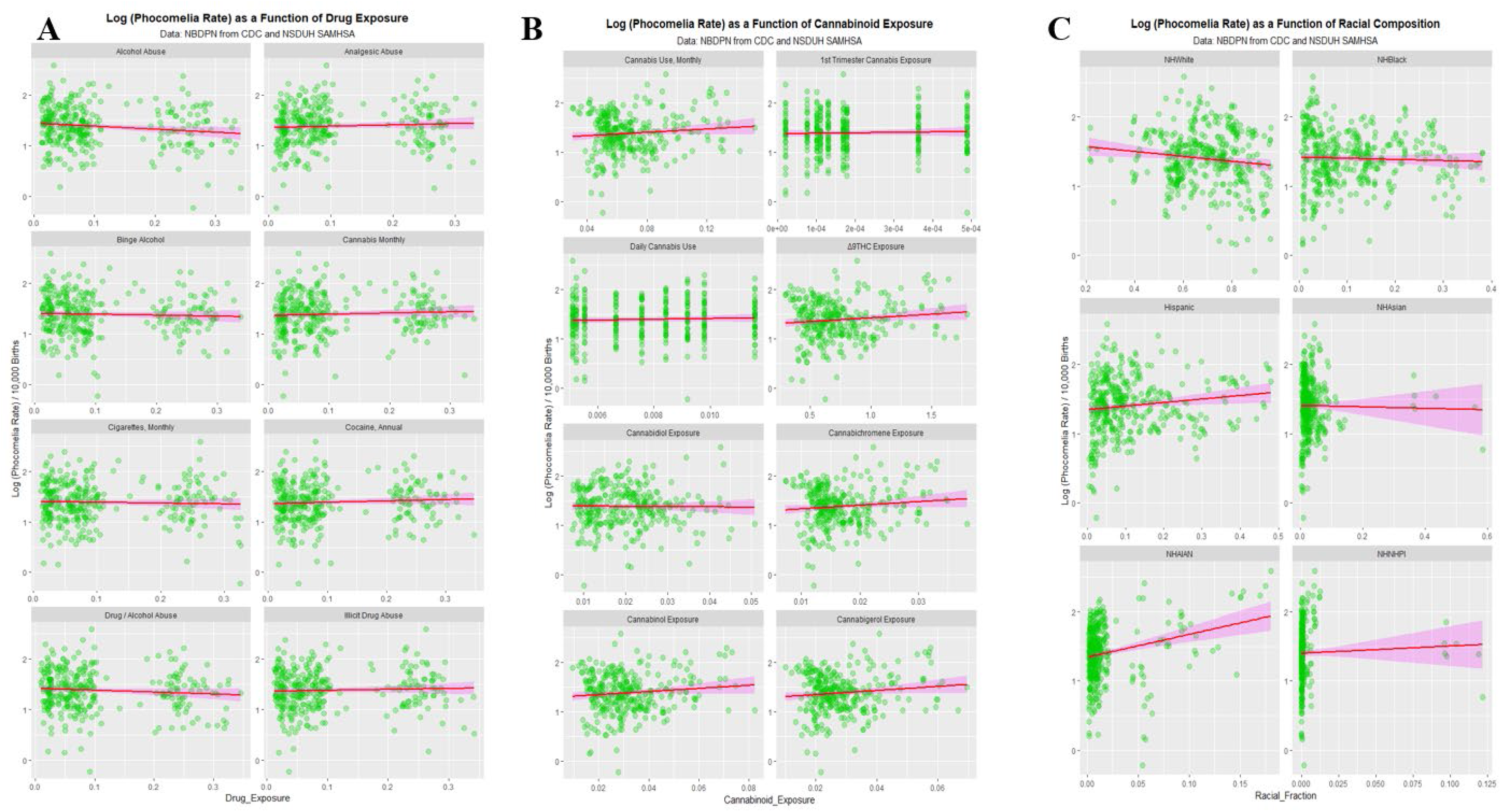
Univariate exposure plots. Limb reduction rates (raw data) as a function of (A) drug exposure, (B) cannabinoid exposure and (C) census-identified ethnicity.

The effects with cannabinoids are formalized in Supplementary Table 5 which shows significant relationships between LRR and Δ9THC, cannabigerol, cannabichromene and cannabinol.

Supplementary Figure 4 shows that the LRR is not impacted by median household income (abbreviation MHY).

These data are well suited to panel regression a technique which was developed within econometrics and which tolerates missing values. The results are shown in Supplementary Table 6. The top half of this table presents regressions done in each domain of socioeconomics, ethnicity and drug exposure. MHY is confirmed not to be significant. Non-Hispanic American Indian / Alaska Native ancestry (NHAIAN) is alone significant amongst the races. When drug exposure, ethnicity and income are combined terms including cannabis exposure (from P=0.0084) and ethnicity are significant. When the model is lagged to two years only drugs, including cannabis, are significant.

It was of interest to conduct spatial regression on these data. As spatial regression algorithms do not permit missing data it became necessary to impute the missing values. Supplementary Table 7 illustrates this from temporal kriging (mean substitution over time). Limiting the data to the period 2005-2014, when it was relatively complete and the drug dataset from NSDUH was also complete, 27 kriged data points (9.3%) were added to the 288 NBDPN dataset to provide the kriged dataset of N=315 for spatial analysis.

Figure 4 shows this kriged data map-graphically. Figure 5 shows the neighbour links which were converted into the spatial weights matrix for the spatial regression.

**Figure 4.:**
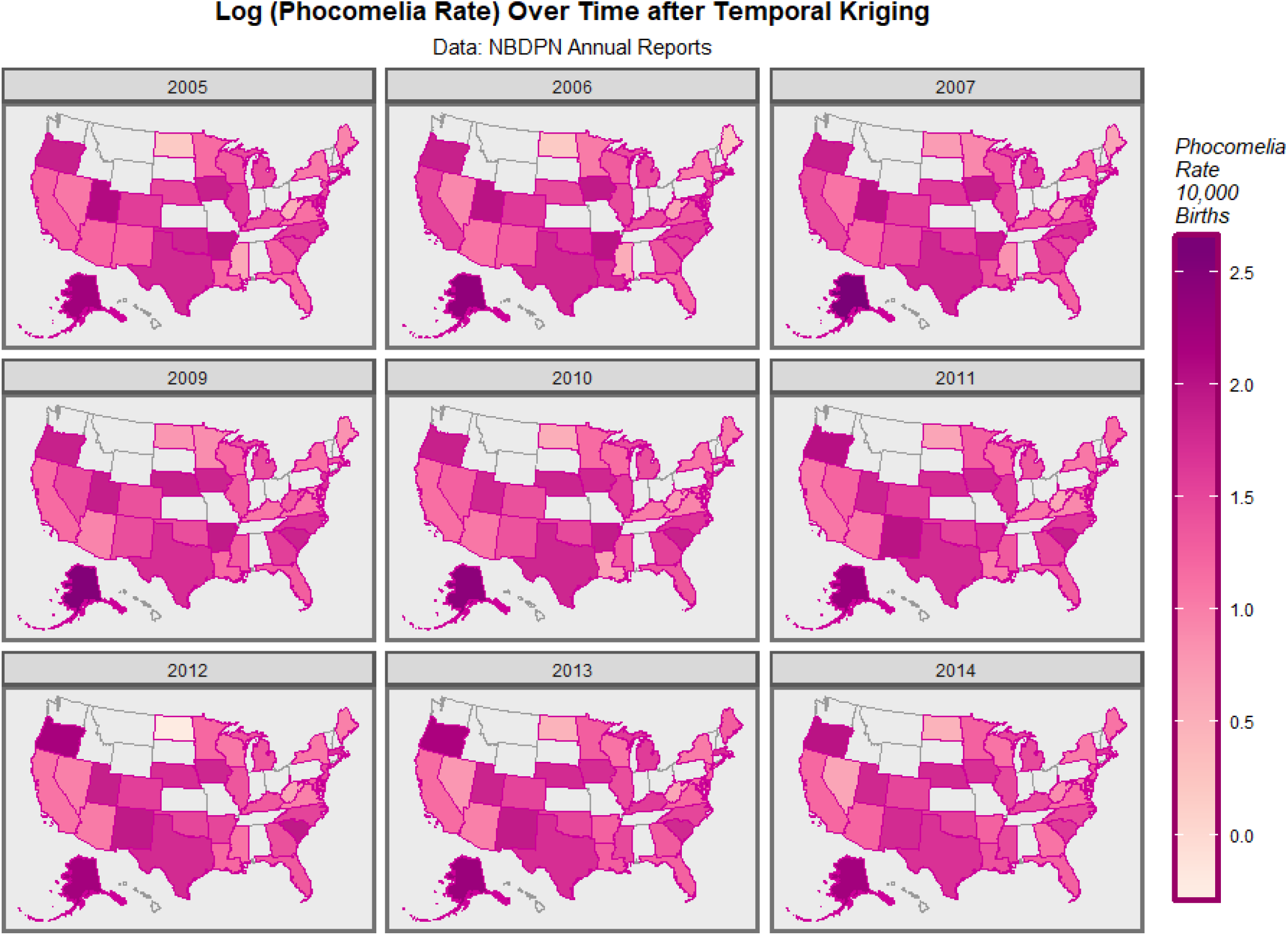
Map-graph of the limb reduction rate across USA over time after temporal kriging 2005-2014.

**Figure 5.:**
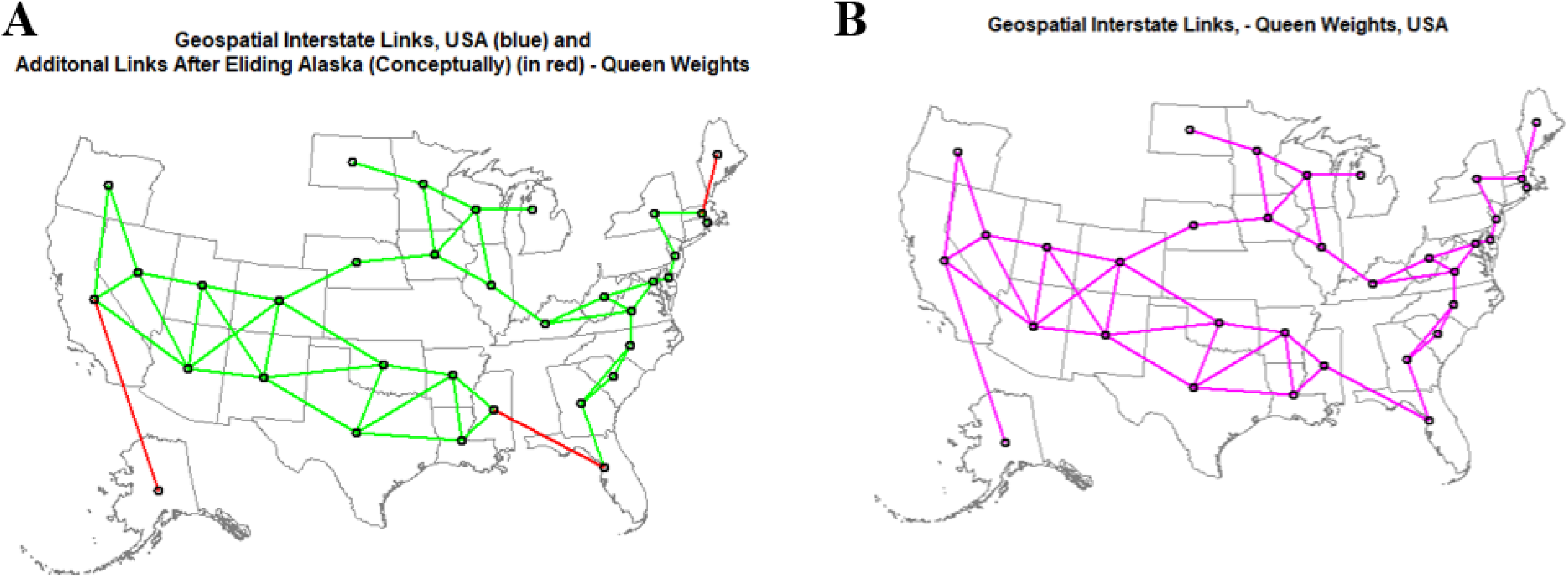
Geospatial links and weights. (A) Editing of polynomial-derived spatial links. (B) Final links used in spatial weights matrix.

Table 1 presents the results of the spatial analysis. Instrumental and lagged variables are shown in the first column of the Table. When considering ethnicity, Hispanic ethnicity is significant. Median household income (MHY) is not significant. In a combined model with all three domains, drugs including cannabis (from β-est.= −4.122 (−6.760, −1.484), P=0.002) and Hispanic ethnicity are significant.

**Table 1.:**
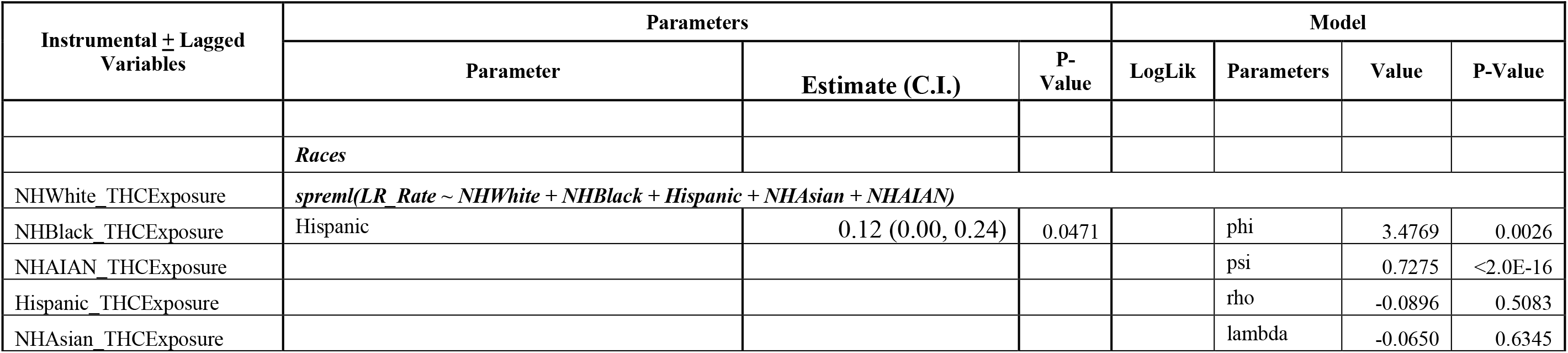
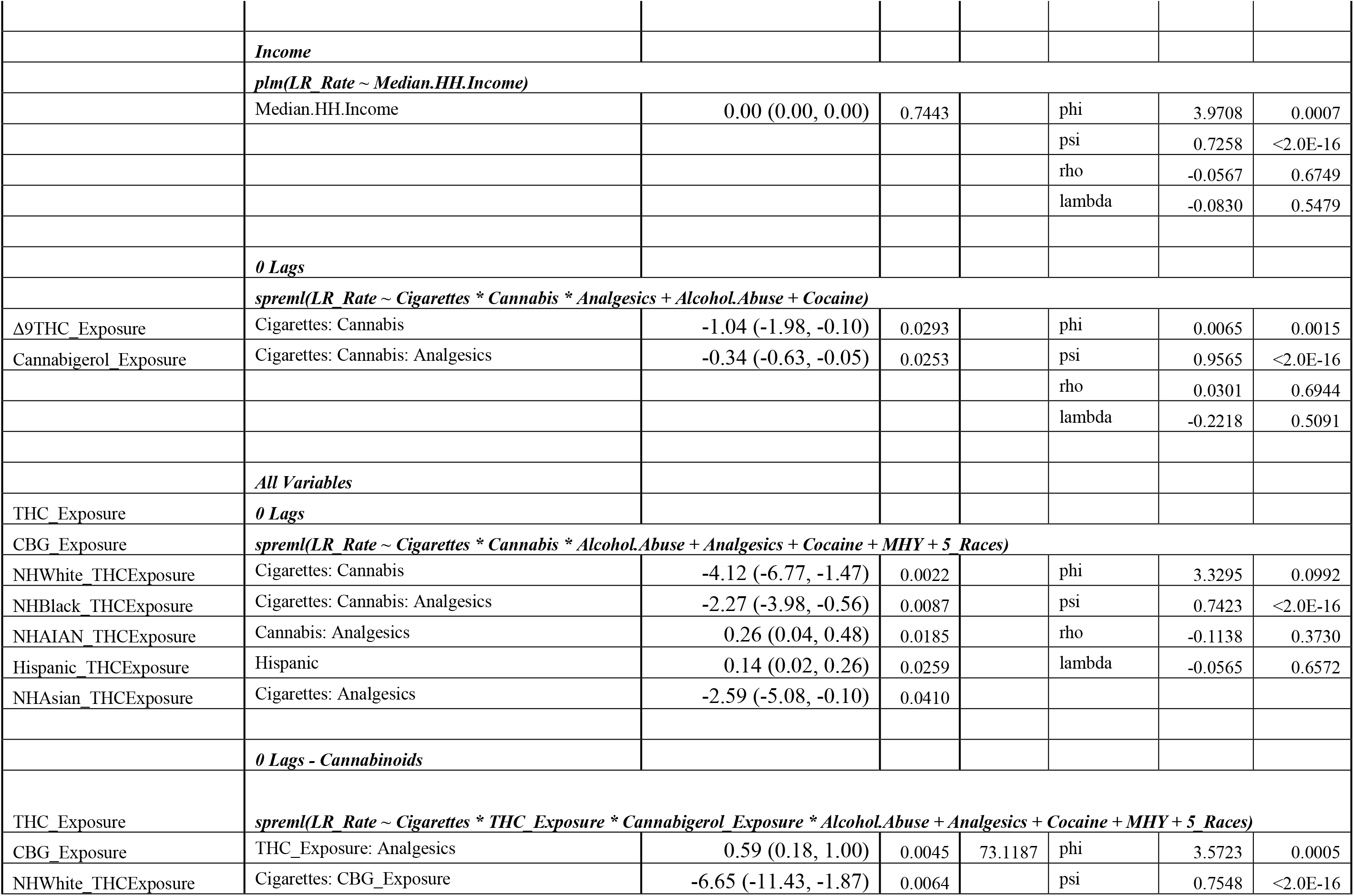
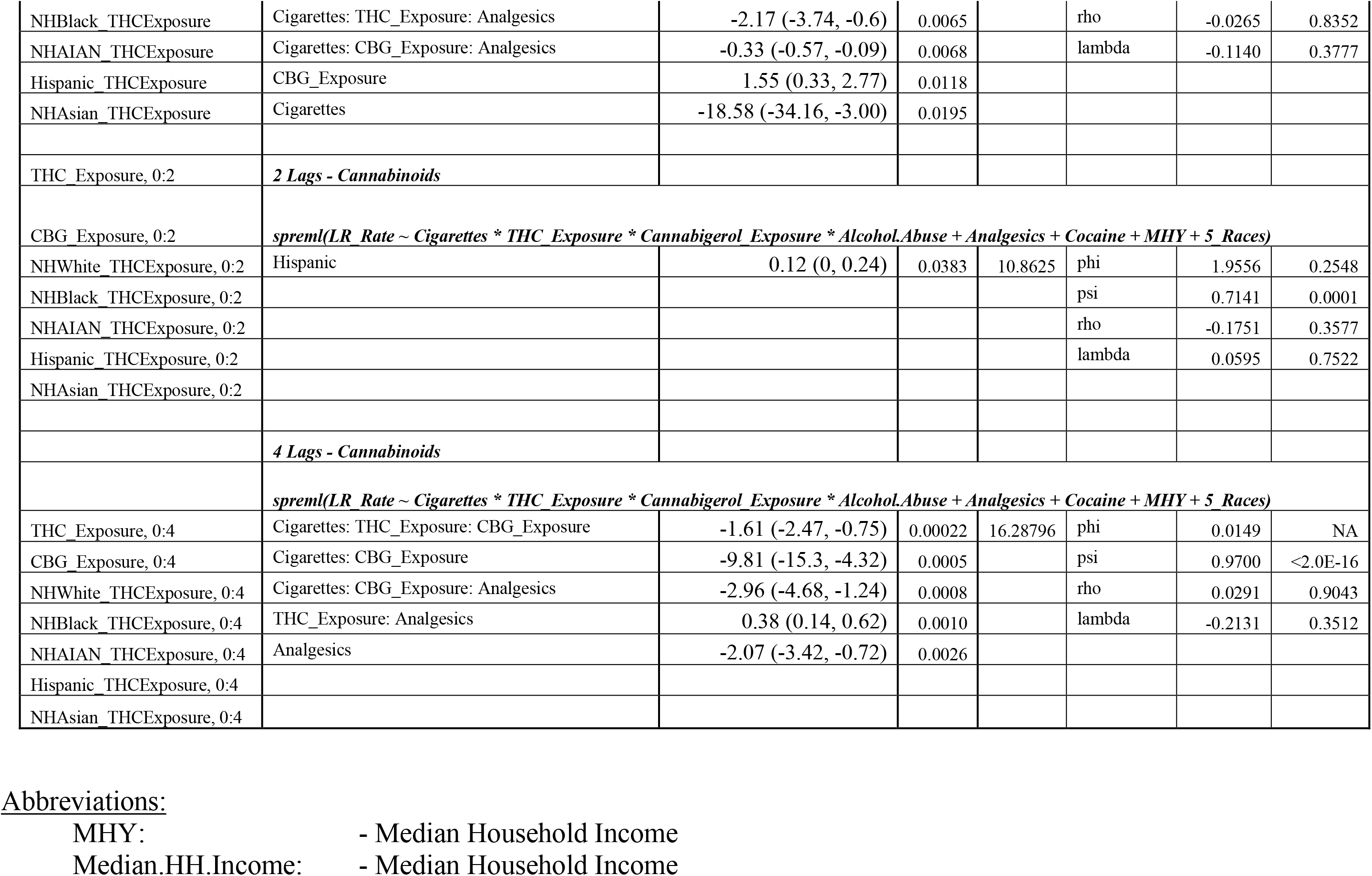
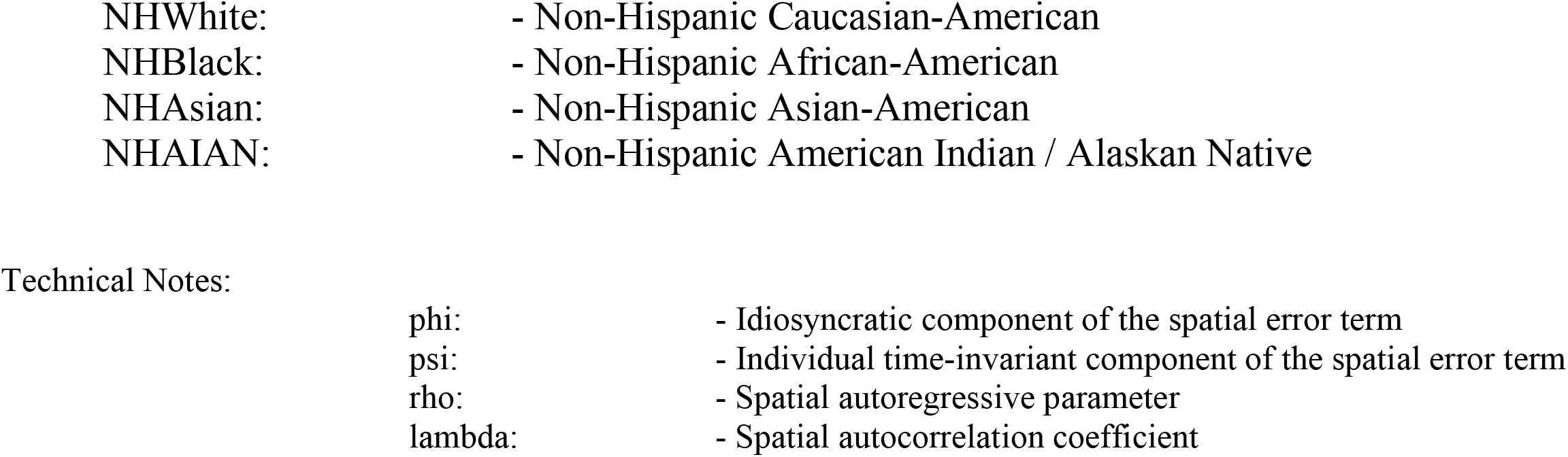
Geospatial spreml Regression of LRR on Drugs, Cannabinoids, Race and Income.

When an interaction between (estimates of) the cannabinoids Δ9THC and CBG were used in place of cannabis the results shown in the lower part of this table were derived. In an unlagged model Δ9THC exposure was significant (from β-est.=0.589 (0.183, 0.995), P=4.5×10^-3^). In a model lagged to four years which better accounts for the moving average style of data published by NBDPN, the Δ9THC: CBG interaction was significant (from β-est.= −1.606 (−2.459, −0.753), P=2.2×10^-4^).

Supplementary Table 8 re-formats data from the tables in the ICBDSR references limiting consideration to only registries reporting positive values for ETOPFA or stillbirths (SB), a procedure suggested by leading public health schools (Mokdad, Dwyer-Lindgren et al. 2017, Roth, Dwyer-Lindgren et al. 2017, Dwyer-Lindgren, Bertozzi-Villa et al. 2018). It shows that 37% of cases from the US registries from Texas, Georgia and Utah were not accounted for in liver birth figures. Worldwide the equivalent figure was 63%. This combined ETOPFA+SB figure is referred to as a “hidden factor” or “silent factor” in our Figures and Tables.

The WARDA series (Supplementary Table 9) was used to convert the reported live birth anomaly rate to a total rate inclusive of ETOPFA’s and stillbirths. The time trend for the estimates of the full dataset appears in Supplementary Figure 5, the time-quintile plot in Supplementary Figure 6, the quintile analysis in Supplementary Figure 7, the dichotomized quintile analysis in Supplementary Figure 8, the map-graphical illustrationin Supplementary Figure 9 and the map-graph for the kriged data in Supplementary Figure 10. In each case apparently significant changes are shown.

Supplementary Figures 11-15 perform a similar role for an ETOPFA/SB factor of 63% as is applicable internationally. Even more striking changes are noted.

Supplementary Figure 16 compares the univariate effects of no hidden factor adjustment, 37% and 63% adjustment on the relationships of the LRR with drug covariates. Adjustment appears to accentuate the changes described above.

Figure 6 shows the effects of these adjustments on cannabinoid covariates, and they again seem to be accentuated.Log (Phocomelia Rate) as a Function of Cannabinoid Exposure

**Figure 6.:**
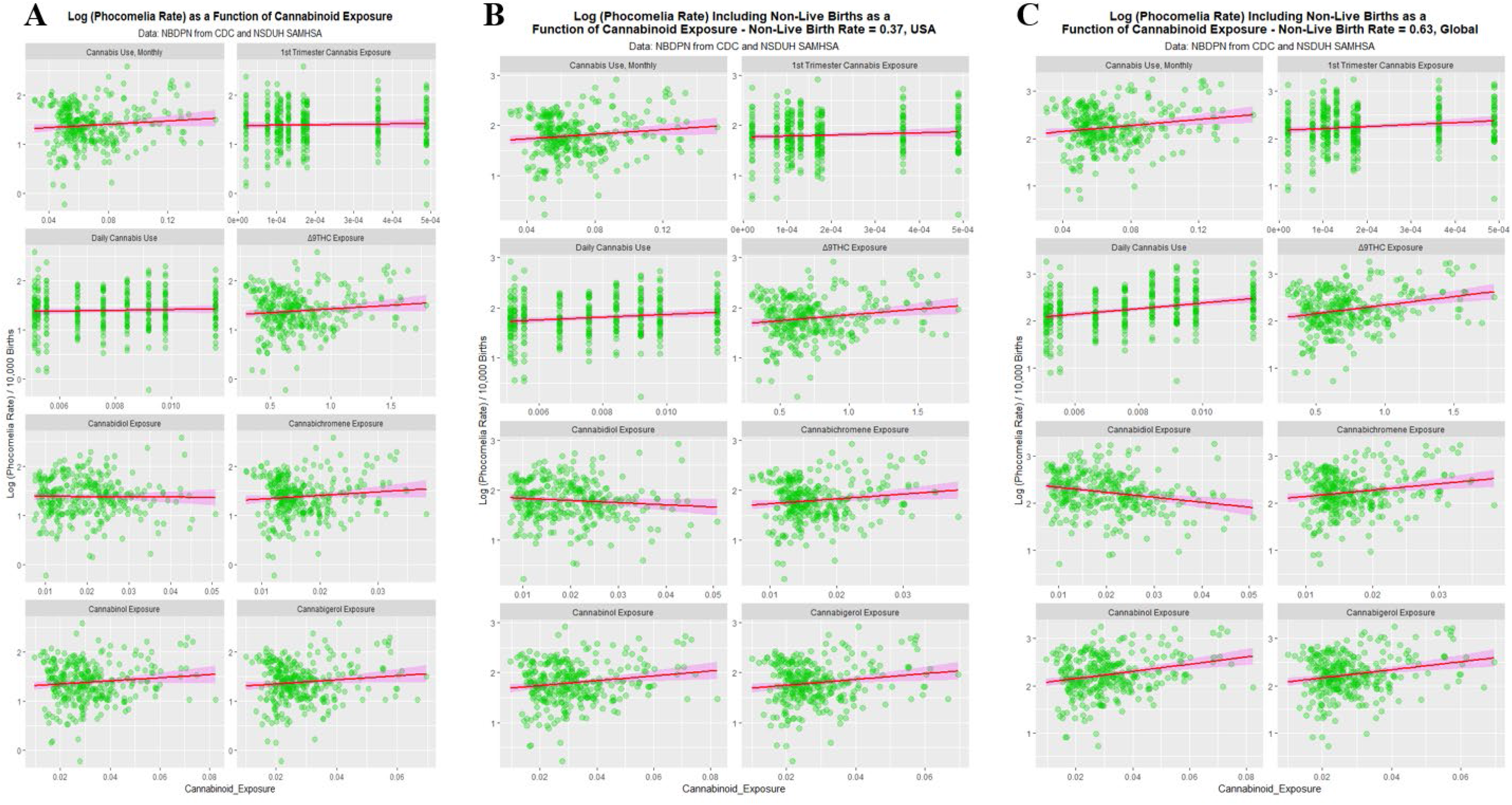
Association plots for cannabinoids with silent factors of (A) 0%, (B) 37% and (C) 63% corresponding to raw data, USA and International data respectively

Supplementary Figure 17 does this for ethnicity with similar effects.

It is conceivable that the type of case finding conducted by the state registries might impact case rates. Supplementary Figure 18 compares the case rates by registry case practice and finds interestingly that the case rates in registries with passive case rates are higher than those in registries practising active or mixed case finding. Compared to passive case finding active and mixed case finding, at a silent factor of zero, were associated with rate reduction (β-estimate= −0.27 (−0.41, −0.14), P=0.0001 and −0.16 (−0.360, −0.33), P=0.0152; model Adj. R^2^ = 0.0846, F=7.935, df=2,148, P=0.0005).

Table 2 and Supplementary Table 10 quantitate these effects by geospatial regression for 37% and 63% hidden factor adjustment. In each case cannabinoids are highly significant (from β-est.=7.474 (3.172, 11.776), P=7×10^-4^ and β-est.=7.190 (2.817, 11.563), P=0.001).

**Table 2.:**
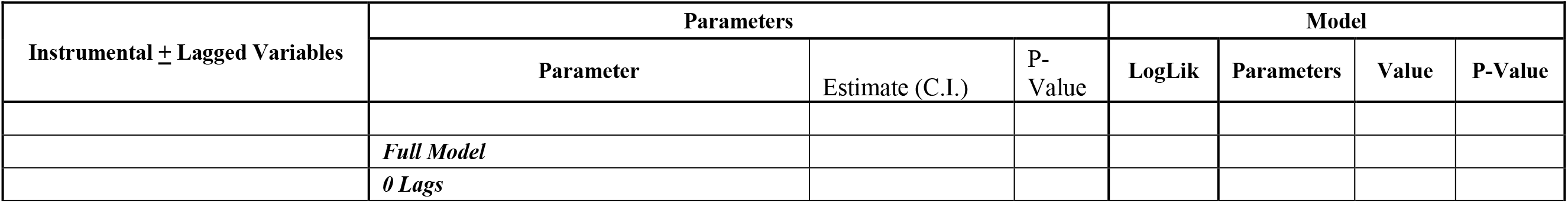
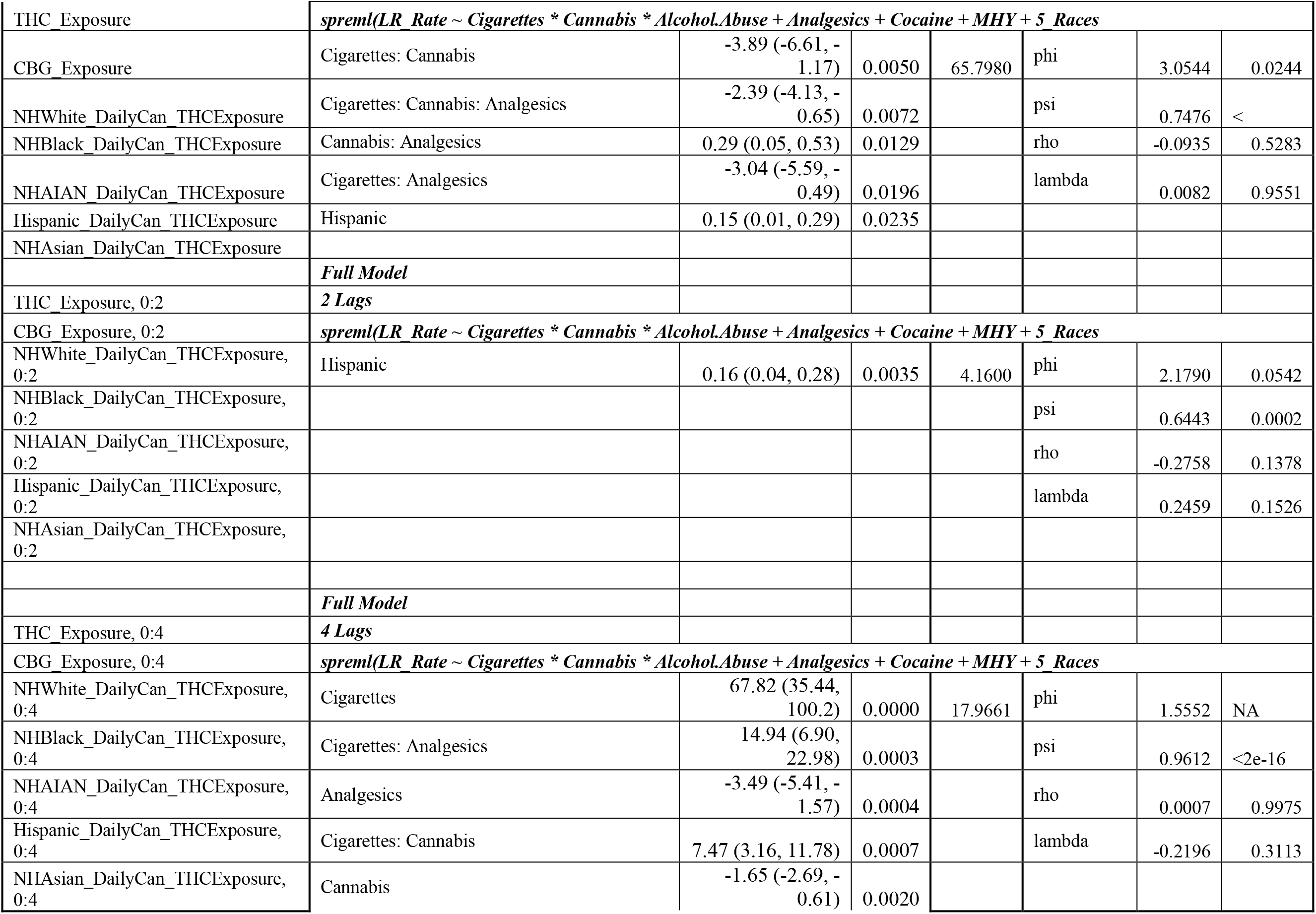
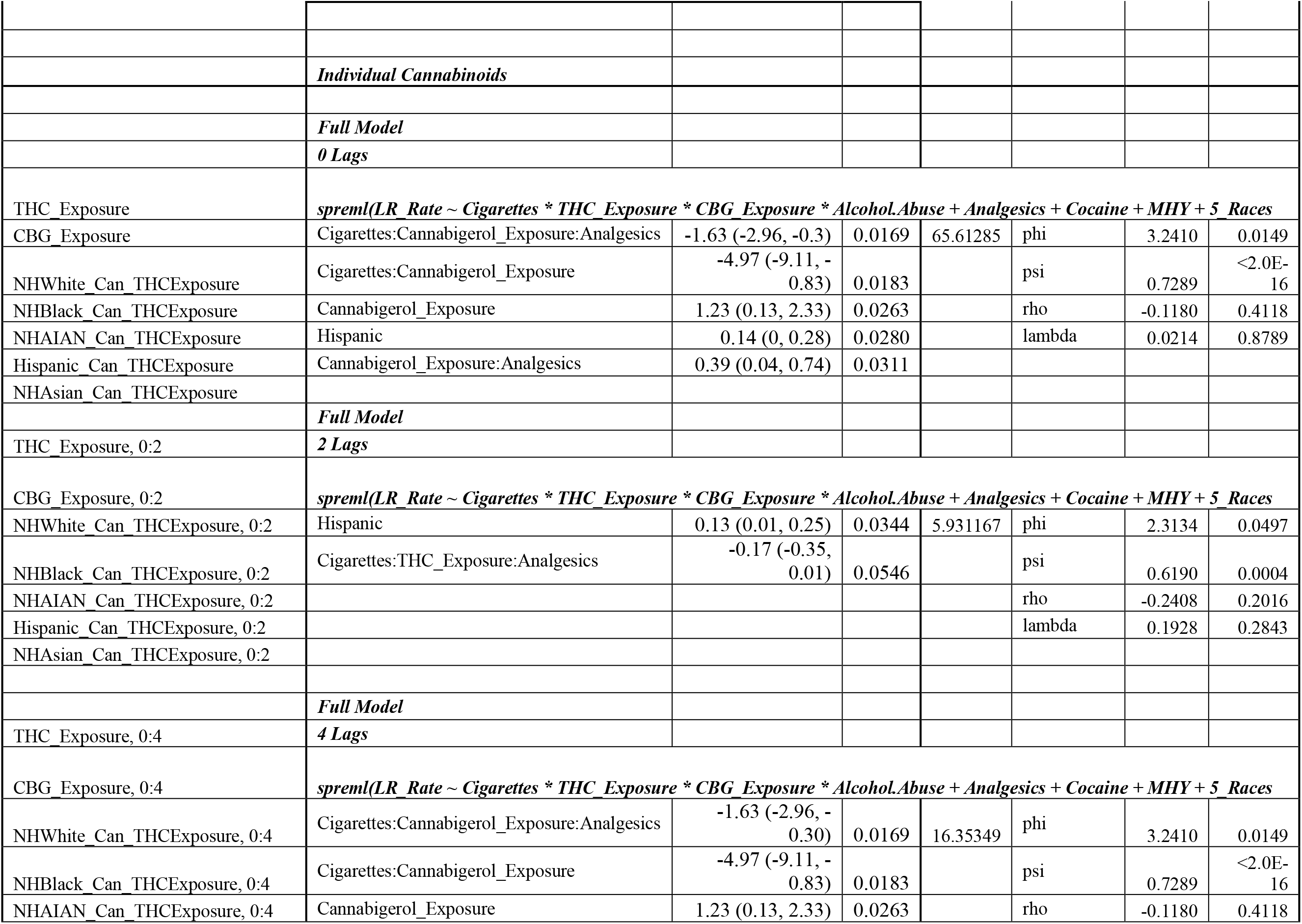
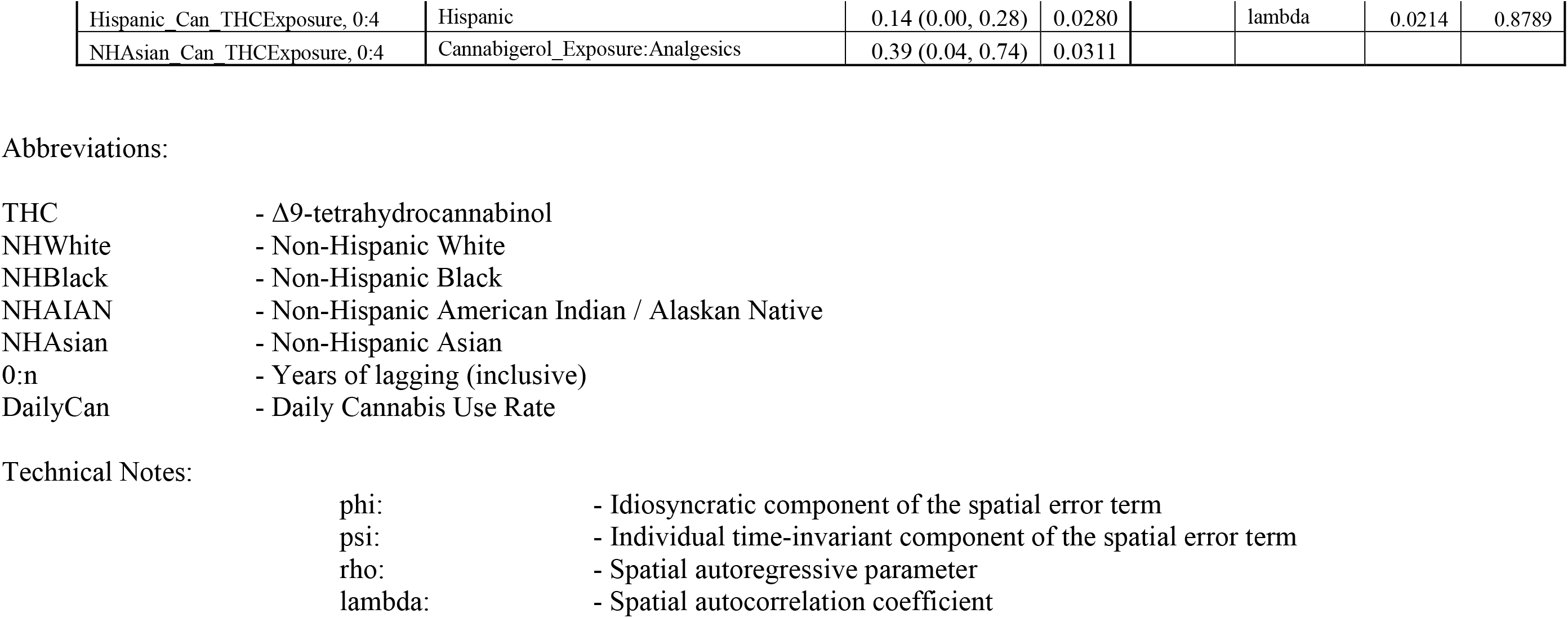
Geospatial spreml Regression of LRR on Drugs, Cannabinoids, Race and Income, USA Silent Factor = 37%.

Figure 7A presents all of the official NBDPN data on the ethnic rates of LR. May zeroes are entered in the data, doubtless most of them meaninglessly or due to low number counts. However low rates are not zero rates as well argued by University of Washington researchers (Mokdad, Dwyer-Lindgren et al. 2017, Roth, Dwyer-Lindgren et al. 2017, Dwyer-Lindgren, Bertozzi-Villa et al. 2018). Following their approach we therefore omit the zeros. Figure 7B shows that this method brings out the high rates in the Non-Hispanic American Indian / Alaska Native (NHAIAN) group. There is no significant trend with time. When the data are adjusted for the US and international hidden factors the appearances shown in Figure 7C and 7D are revealed.

**Figure 7.:**
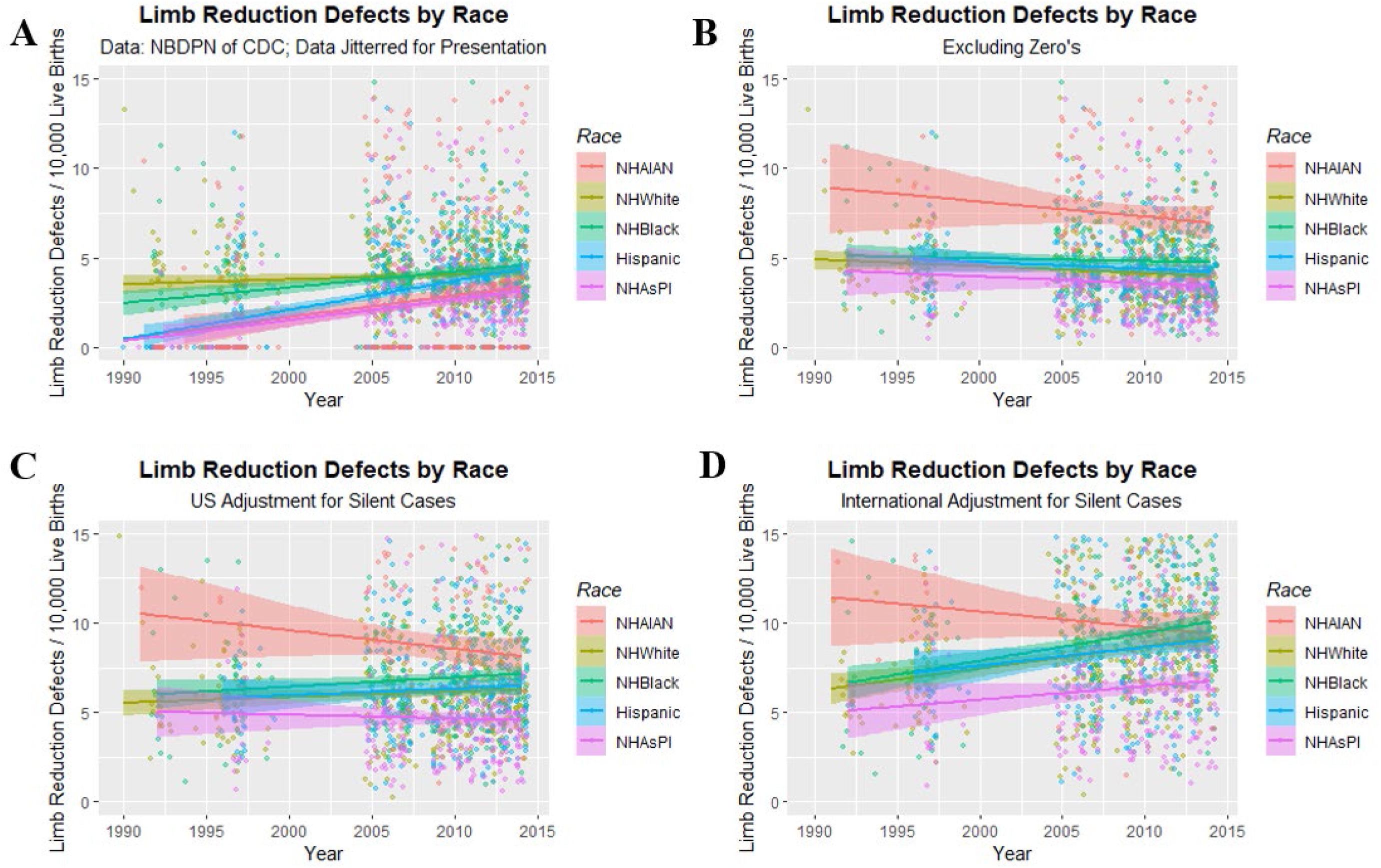
Limb reduction rates by ethnic background (NBDPN data). (A) Limb reduction rates over time by ethnicity. (B) Limb reduction rates by ethnicity over time omitting uninformative zero’s from data. (C) Same as (B) but adjusted for US “silent case” rate (37%) arising from ETOPFA’s and stillbirths. (D) Same as (B) but adjusted for global “silent case” rate (63%) arising from ETOPFA’s and stillbirths.

Supplementary Table 11 presents two-step regression results performed in R::AER using the Asian / Pacific Islander group as the controls and the unadjusted LRR as the dependent variable. Very highly significant results are shown with the effects for NHAIAN and Non-Hispanic African-Americans significant (from β-est.=0.999 (0.885, 1.113), P<2.2×10^-16^, and β-est.=0.449 (0.357, 0.541), P<2.2×10^-16^). However when the days of use of cannabis by each ethnicity and the THC potency of the cannabis smoked are used as interactive instrumental variables this effect completely disappears. When the full complement of instrumental variables is employed in the exhaustive model only the NHAIAN group is significant.

This important result shows that whilst racial factors are apparently strong, they are completely accounted for by substance and particularly cannabinoid exposure and are thus not robust to adjustment.

Figure 8 presents the effects of cannabis legalization policies on LRR. The legalized status appears to have a higher LRR than comparators. Panel B dichotomizes the legal status into legalized status compared to the others.

**Figure 8.:**
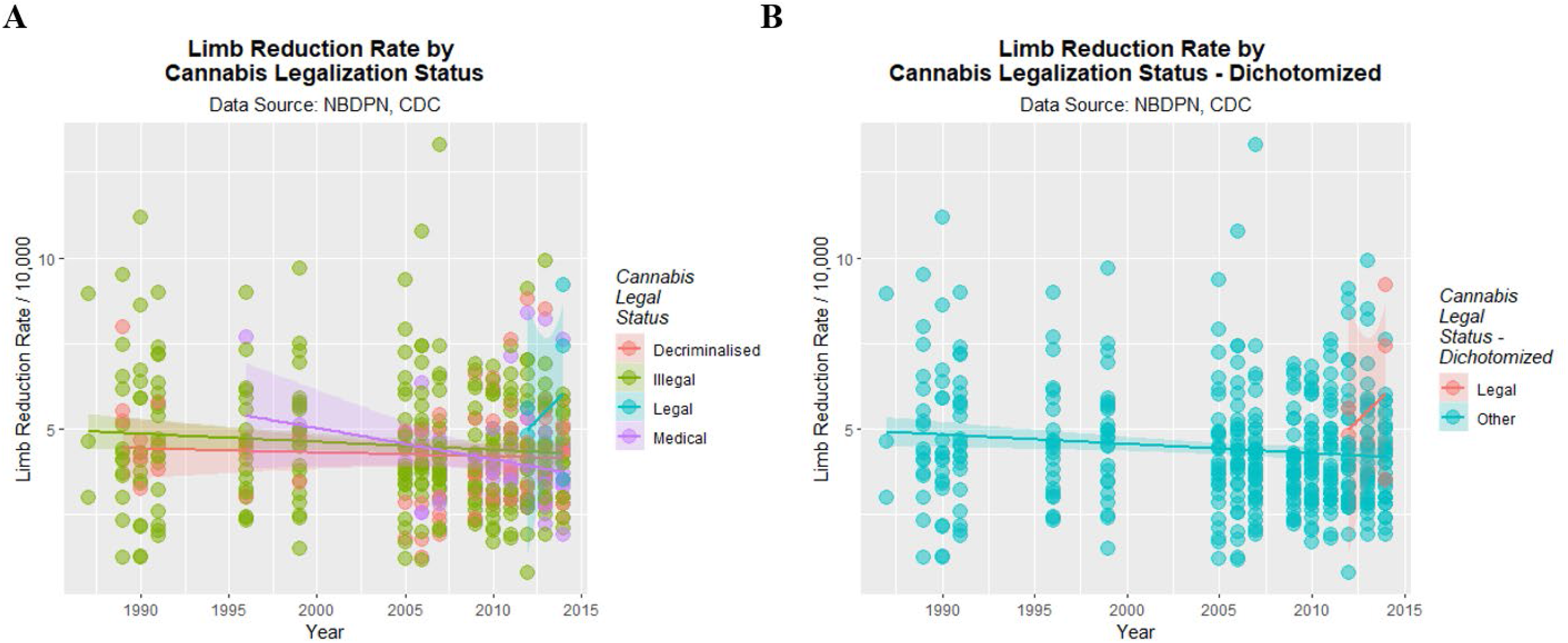
Limb reduction rates by (A) cannabis legal status and (B) cannabis legal status dichotomized into legalized vs. other legal statuses. Data not corrected for silent factors.

Table 3 quantifies these effects with linear regression with silent factors of 0%, 36% and 63%. Increasingly significant changes are noted. Supplementary Table 12 presents the numbers of cases and controls in each of the legal categories dichotomized to legal states vs. others.

**Table 3.:** Time: Status Interactions by Silent Factor for Dichotomized Legal Status.

| *Silent Factor* | Parameter |  |  | Model Parameters |  |  |  |
| --- | --- | --- | --- | --- | --- | --- | --- |
|  | Parameter | Estimate (C.I.) | P-Value | Adj. R-Squared | F | dF | P-value |
|  | *lm(LR Rate ~ Legal Status)* |  |  |  |  |  |  |
| **0%** | Year:Status_Legal | 0.0007 (0.0001, 0.0013) | 0.0396 | 0.0139 | 4.055 | 2,432 | 0.0180 |
| **37%** | Year:Status_Legal | 0.0011 (0.0001, 0.0021) | 0.0236 | 0.0381 | 9.587 | 2,432 | 8.4E-05 |
| **63%** | Year:Status_Legal | 0.0020 (0.0006, 0.0034) | 0.0071 | 0.1951 | 53.590 | 2,432 | <2.0E-16 |

Table 4A presents the increased relative risk (RR) associated with cannabis legalization and Table 4B presents the relative risk reduction associated with non-cannabis legalized status. As can be seen in Table 4A the elevated RR of LRR is 1.17 (1.02, 1.34), 1.26 (1.13, 1.40) and 1.39 (1.289, 1.51) for 0%, 36% and 63% silent factors respectively. The table also presents attributable risks in the exposed, attributable risks in the population and applicable P-values. Table 4B presents the inverse of these results as relative risk reductions (RRR).

**Table 4.:** Relative Risk Calculations for: (A) Risk Elevation by Legal Status and (B) Risk Reduction by Not Legal Status.

|  | LR+ | LR- | Total | Risk | Incr RR | AFE | AFP | P-values |
| --- | --- | --- | --- | --- | --- | --- | --- | --- |
| ***0% Silent Factor*** |  |  |  |  |  |  |  |  |
| Legal_States | 215 | 428,805 | 429,020 | 0.0005 |  |  |  |  |
| Others | 14,398 | 33,630,778 | 33,645,176 | 0.0004 | 1.17 (1.02 to 1.34) | 14.61 (2.30 to 25.36) | 0.21 (0.02 to 0.41) | 0.0212 |
| ***36% Silent Factor*** |  |  |  |  |  |  |  |  |
| Legal_States | 338 | 428,682 | 429,020 | 0.0008 |  |  |  |  |
| Others | 21,106 | 33,624,070 | 33,645,176 | 0.0006 | 1.26 (1.13 to 1.40) | 20.38 (11.35 to 28.49) | 0.32 (0.15 to 0.49) | 3.1E-05 |
| ***63% Silent Factor*** |  |  |  |  |  |  |  |  |
| Legal_States | 566 | 428,454 | 429,020 | 0.0013 |  |  |  |  |
| Others | 31,954 | 33,613,222 | 33,645,176 | 0.0009 | 1.39 (1.28 to 1.51) | 28.01 (21.78 to 33.75) | 0.49 (0.34 to 0.63) | 6.7E-15 |

**(B) Relative Risk Reduction Accompanying Absence of Cannabis Legalization**
|  | LR+ | LR- | Total | Risk | RRR | AFE | AFP | P-values |
| --- | --- | --- | --- | --- | --- | --- | --- | --- |
| ***0% Silent Factor*** |  |  |  |  |  |  |  |  |
| Others | 14,398 | 33,630,778 | 33,645,176 | 0.0004 |  |  |  |  |
| Legal_States | 215 | 428,805 | 429,020 | 0.0005 | 0.15 (0.02 to 0.25) | -17.11 (-33.98 to -2.36) | -16.85 (-33.43 to -2.34) | 0.0212 |
| **36% Silent Factor** |  |  |  |  |  |  |  |  |
| Others | 21,106 | 33,624,070 | 33,645,176 | 0.0006 |  |  |  |  |
| Legal_States | 338 | 428,682 | 429,020 | 0.0008 | 0.20 (0.11 to 0.28) | -25.59 (-39.83 to -12.80) | -25.19 (-39.15 to -12.63) | 3.1E-05 |
| **63% Silent Factor** |  |  |  |  |  |  |  |  |
| Others | 31,954 | 33,613,222 | 33,645,176 | 0.0009 |  |  |  |  |
| Legal_States | 566 | 428,454 | 429,020 | 0.0013 | 0.28 (0.21 to 0.34) | -38.91 (-50.94 to -27.84) | -38.23 (-49.99 to -27.40) | 6.7E-15 |

Abbreviations:
- Limb Reductions
- No Limb Reductions (Otherwise normal births)
Incr
- Increased
- Relative Risk
RRR
- Relative Risk Reduction
AFE
- Attributable Fraction in Exposed

As the kriged dataset is balanced it lends itself to analysis by the techniques of formal causal inference including the calculation of inverse probability weights (IPW). IPW were therefore calculated on the kriged data with a 37% silent factor.

Table 5 shows the results of robust regression conducted in the package survey in R using IPW weights. The final model from a purely additive model, a model using a four way interaction between tobacco: binge alcohol: cannabis: analgesics, and a five-way interactive model between tobacco: binge alcohol: Δ9THC: cannabigerol: analgesics are shown. Median household income, ethnicity and ethnic cannabis use scores are covariates. As shown in the second model eight terms including cannabis appear including four terms relating to ethnic cannabis use. Eight terms including cannabinoids also appear in the final model.

**Table 5.:**
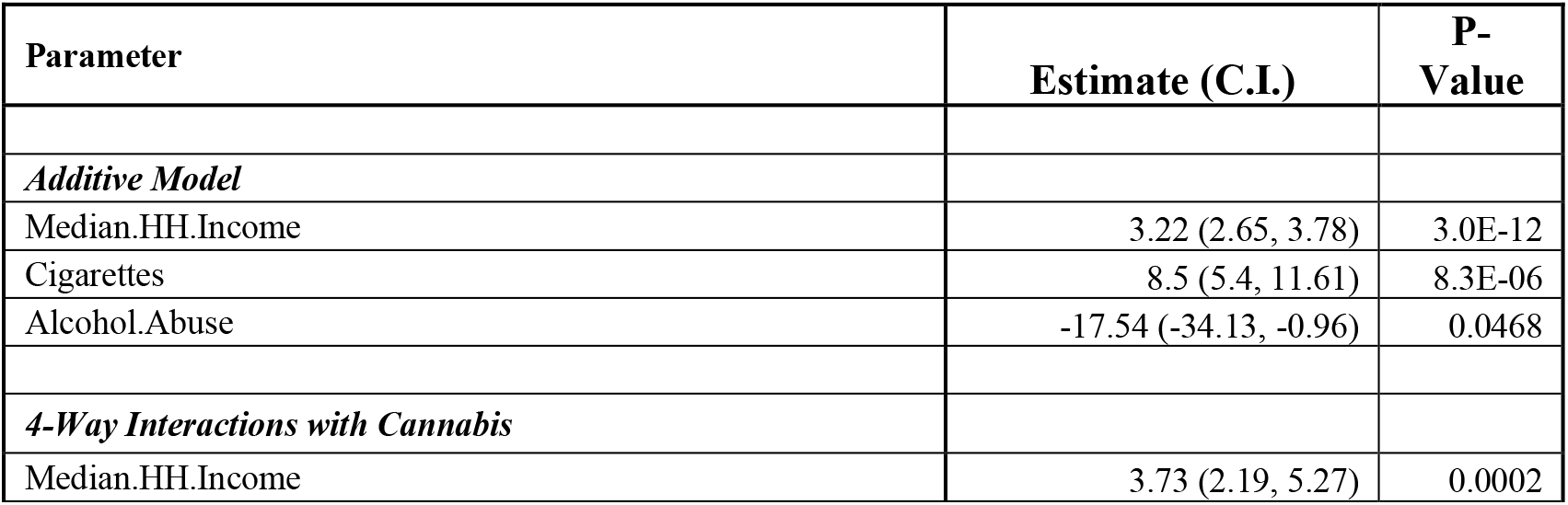
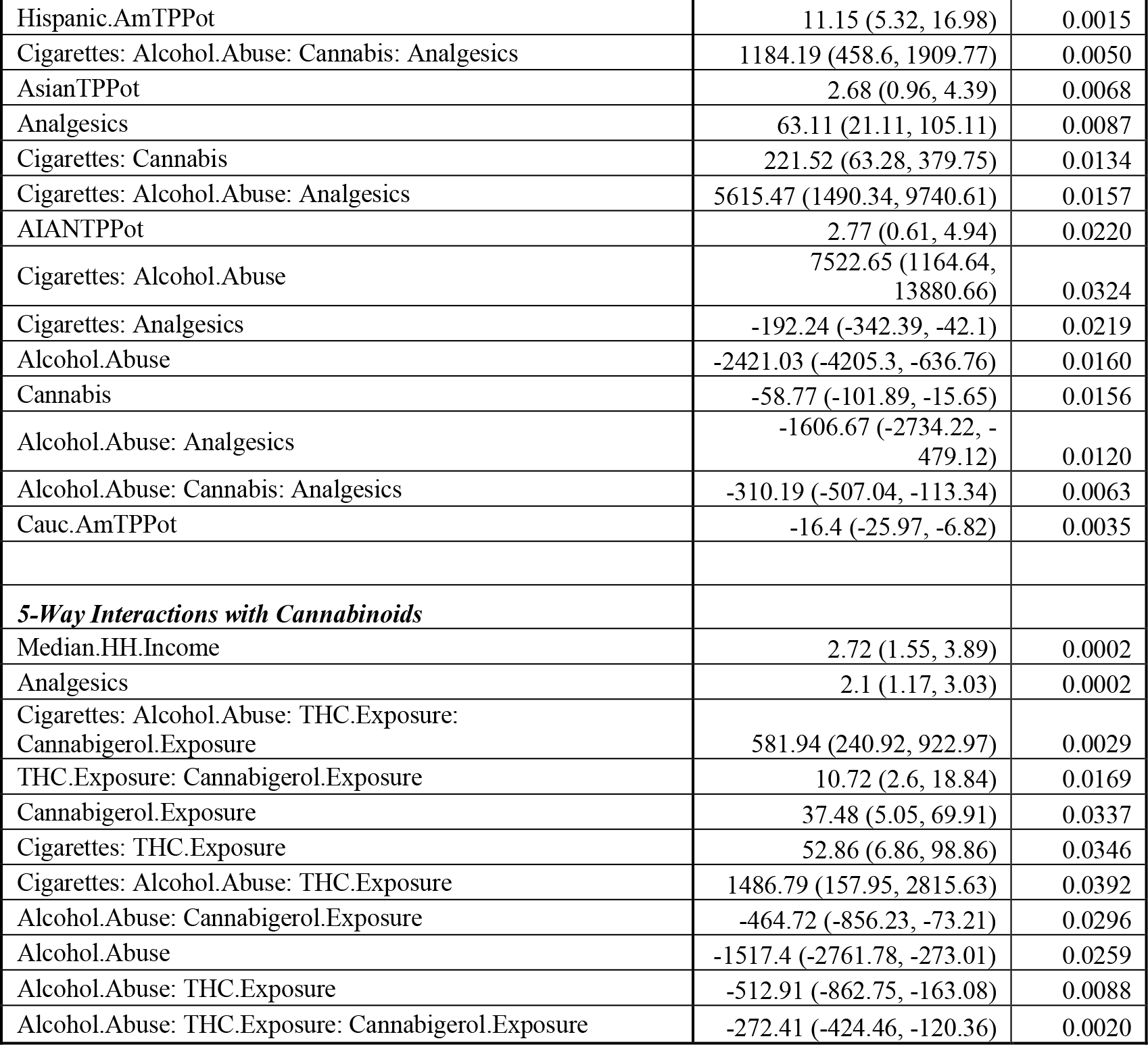
Robust Generalized Linear Regression Utilizing Inverse Probability Weights.

| Parameter | Estimate (C.I.) | P-Value |
| --- | --- | --- |
| *Additive Model* |  |  |
| Median.HH.Income | 3.22 (2.65, 3.78) | 3.0E-12 |
| Cigarettes | 8.5 (5.4, 11.61) | 8.3E-06 |
| Alcohol.Abuse | -17.54 (-34.13, -0.96) | 0.0468 |
| *4-Way Interactions with Cannabis* |  |  |
| Median.HH.Income | 3.73 (2.19, 5.27) | 0.0002 |
| Hispanic.AmTPPot | 11.15 (5.32, 16.98) | 0.0015 |
| Cigarettes: Alcohol.Abuse: Cannabis: Analgesics | 1184.19 (458.6, 1909.77) | 0.0050 |
| AsianTPPot | 2.68 (0.96, 4.39) | 0.0068 |
| Analgesics | 63.11 (21.11, 105.11) | 0.0087 |
| Cigarettes: Cannabis | 221.52 (63.28, 379.75) | 0.0134 |
| Cigarettes: Alcohol.Abuse: Analgesics | 5615.47 (1490.34, 9740.61) | 0.0157 |
| AIANTPPot | 2.77 (0.61, 4.94) | 0.0220 |
| Cigarettes: Alcohol.Abuse | 7522.65 (1164.64, 13880.66) | 0.0324 |
| Cigarettes: Analgesics | -192.24 (-342.39, -42.1) | 0.0219 |
| Alcohol.Abuse | -2421.03 (-4205.3, -636.76) | 0.0160 |
| Cannabis | -58.77 (-101.89, -15.65) | 0.0156 |
| Alcohol.Abuse: Analgesics | -1606.67 (-2734.22, -479.12) | 0.0120 |
| Alcohol.Abuse: Cannabis: Analgesics | -310.19 (-507.04, -113.34) | 0.0063 |
| Cauc.AmTPPot | -16.4 (-25.97, -6.82) | 0.0035 |
| **5-Way Interactions with Cannabinoids** |  |  |
| Median.HH.Income | 2.72 (1.55, 3.89) | 0.0002 |
| Analgesics | 2.1 (1.17, 3.03) | 0.0002 |
| Cigarettes: Alcohol.Abuse: THC.Exposure: Cannabigerol.Exposure | 581.94 (240.92, 922.97) | 0.0029 |
| THC.Exposure: Cannabigerol.Exposure | 10.72 (2.6, 18.84) | 0.0169 |
| Cannabigerol.Exposure | 37.48 (5.05, 69.91) | 0.0337 |
| Cigarettes: THC.Exposure | 52.86 (6.86, 98.86) | 0.0346 |
| Cigarettes: Alcohol.Abuse: THC.Exposure | 1486.79 (157.95, 2815.63) | 0.0392 |
| Alcohol.Abuse: Cannabigerol.Exposure | -464.72 (-856.23, -73.21) | 0.0296 |
| Alcohol.Abuse | -1517.4 (-2761.78, -273.01) | 0.0259 |
| Alcohol.Abuse: THC.Exposure | -512.91 (-862.75, -163.08) | 0.0088 |
| Alcohol.Abuse: THC.Exposure: Cannabigerol.Exposure | -272.41 (-424.46, -120.36) | 0.0020 |

A similar exercise is conducted with IPW-weighted mixed effects regression with results shown in Supplementary Table 13. The models were of similar structure used to those employed for robust regression in Table 5. The benefit of using these models was that mixed effects models have a model standard deviation which can be entered into e-value calculations, which is absent from robust models generated in the survey package. As shown in Table 5 terms including cannabis or cannabinoids are significant and appear in final models in the additive, four-way cannabis, and five-way cannabinoid models from very high levels of statistical significance.

Finally e-values were calculated from some of the major results above and are illustrated in Table 6. It is clear that many of the e-values are very large, implying that some unmeasured confounder would require a large degree of co-association with both the outcome and the exposure (mrjmon) to impact the results.

**Table 6.:** Selected E-Values.

| Parameter | Table | Regression Coefficient (C.I.) | R.R. (C.I.) | eValues |
| --- | --- | --- | --- | --- |
| ***Linear Regression*** |  |  |  |  |
| LR over Time | eTable 4 | -0.21 (-0.043, 0.00012) | 0.9892 (0.9785, 1.00001) | 1.12, 1.00 |
| LR by Monthly Cannabis Use | eTable 4 | 8.51 (3.11, 4.32) | 95.17 (1.04, 8685.75) | 189.85, 1.25 |
| LR by Cannabis Use Quintile | eTable 4 | 0.43 (0.28, 0.59) | 2.72 (1.92, 3.83) | 4.87, 3.25 |
| LR by Cannabis Use Quintile Dichotomized | eTable 4 | 0.42 (0.27, 0.56) | 2.59 (1.86, 3.59) | 4.62, 31.4 |
| ***Legal Relationship*** |  |  |  |  |
| Cannabis Legalization, Silent Factor = 0% | Table 4 | 1.17 (1.02, 1.34) | 1.17 (1.02, 1.34) | 1.62, 1.16 |
| Cannabis Legalization, Silent Factor = 37% | Table 4 | 1.26 (1.13, 1.40) | 1.26 (1.13, 1.40) | 1.83, 1.51 |
| Cannabis Legalization, Silent Factor = 63% | Table 4 | 1.39 (1.28, 1.51) | 1.39 (1.28, 1.51) | 2.13, 1.88 |
| ***Geotemporospatial Regression - 37% Silent Factor*** | Table 2 |  |  |  |
| *No Lag* | Table 2 |  |  |  |
| Cannabis: Analgesics | Table 2 | 0.28 (0.06, 0.51) | 2.11 (1.17, 3.82) | 3.65, 1.62 |
| *4 Lags* |  |  |  |  |
| Cannabis: Tobacco | Table 2 | 7.47 (3.17, 11.78) | 4.44E+19 (2.30E+08, 8.58E+30) | 8.89E+19, 4.61E+08 |
| *Individual Cannabinoids* | Table 2 |  |  |  |
| *No Lags* | Table 2 |  |  |  |
| Cannabigerol | Table 2 | 1.23 (0.15, 2.32) | 23.85 (1.46, 389.22) | 47.19, 2.28 |
| Cannabigerol: Analgesics | Table 2 | 0.38 (0.035, 0.74) | 2.70 (1.09, 6.66) | 4.85, 1.42 |
| *4 Years Lag* | Table 2 |  |  |  |
| Tobacco: THC | Table 2 | 9.14 (4.22, 14.07) | 4.31E+31 (4.30E+14, 4.34E+48) | 8.62E+31, 6.61E+14 |
| Cannabigerol: Analgesics | Table 2 | 0.61 (0.26, 0.96) | 133.81 (8.21, 2.18E+03) | 267.13, 15.91 |
| ***Mixed Effects iptw Regression*** |  |  |  |  |
| *Additive Model* | eTable 13 |  |  |  |
| Afric.American.THc.Exposure | eTable 13 | 0.61 (0.57, 0.65) | 1.000 (1.000, 1.000) | 1.0048, 1.0046 |
| *Interactive Model* |  |  |  |  |
| NHWhite.THc.Exposure | eTable 13 | 4.06 (3.7, 4.42) | 1.005 (1.004, 1.006) | 1.024, 1.022 |
| Cigarettes: Cannabis: Analgesics | eTable 13 | 480.35 (337.18, 623.51) | 1.065 (1.045, 1.085) | 1.32, 1.26 |
| Cigarettes: Cannabis | eTable 13 | 1462.8 (1017.39, 1908.22) | 1.21 (1.14, 1.28) | 1.72, 1.54 |
| Alcohol.Abuse: Cannabis: Analgesics | eTable 13 | 1102.5 (626.32, 1578.68) | 1.16 (1.09, 1.23) | 1.57, 1.39 |
| Alcohol.Abuse: Cannabis | eTable 13 | 3224.18 (1743.59, 4704.78) | 1.53 (1.26, 1.85) | 2.42, 1.83 |

## Discussion

### Statement of Principal Findings

The key results of the study were that LRR was significantly associated with cannabis use, and in a geospatiotemporal context with exposure to the cannabinoids Δ9THC and cannabigerol, although there was no time trend. Moreover these changes were robust to adjustment for common sociodemographic covariates including socioeconomic and ethnographic factors. There was a marked quintile effect with an apparently sharp jump from the fourth to fifth quintile. The marked effects of race were completely dissipated by adjustment for cannabinoid exposure. 40-60% of the cases are not accounted for in live birth rates. When either a US or an international adjustment correction is applied to these numbers the changes described for the raw rates become intensified and more significant. Importantly the legal paradigm relating to cannabis regulation was shown to be highly significant with a relative risk reduction (RRR) of 15% (95%C.I. 0.02 to 0.25) for states where cannabis had not been legalized increasing to 28% (C.I. 0.21 to 0.34) protection for a 63% international silent factor protecting from 38% (27.40 to 49.99%) of the population attributable caseload (also at 63% silent factor). Formal assessment of the potentially causal nature of the cannabis – LRR relationship by IPW-weighted robust and mixed effects regression and the calculation of e-values, confirmed that the association fulfilled formal criteria for a causal relationship.

### Consistency with Other Reports

Our results are supported by the prior results from Hawaii (Forrester and Merz 2007) and also apparently by reports from German obstetric hospitals and French birth defect registries where outbreaks of LR have been described (Agence France-Presse in Paris 2018, Willsher K. 2018, Robinson M. 2019). The odds ratio reported in France was 58-times elevation (Agence France-Presse in Paris 2018, Willsher K. 2018) which is within the confidence interval reported from Hawaii (95%C.I. 4.45, 65.63). Similarly French cows in these areas are more frequently born without forelimbs. Interestingly, both France and Germany along with a number of other EU counties allow cannabinoids to enter the food chain whereas Switzerland does not allow this practice. Public health enquiries in this regard are on-going. Nearby Switzerland has not seen any such increase in cases.

### Possible Explanations for Study Findings

It is important to note that cannabis can act by several cellular mechanisms to restrict cell growth and inhibit cell division (McClean and Zimmerman 1976, Zimmerman and Raj 1980, Tahir and Zimmerman 1991) and can act genomically and epigenomically (Zimmerman A.M. and Zimmerman S. 1987, Zimmerman and Zimmerman 1990). Its mitochondrial inhibitory actions (Bartova and Birmingham 1976, Hebert-Chatelain, Reguero et al. 2014, Wolff, Schlagowski et al. 2015, Hebert-Chatelain, Desprez et al. 2016) carry serious downstream genotoxic and epigenotoxic implications by reducing cellular energy charge for DNA-dependent reactions (Canto, Menzies et al. 2015), by limiting the availability of numerous chemical moieties which underpin epigenomic regulation and by inducing mitonuclear stress response cascade (Canto, Menzies et al. 2015). Cannabinoid receptors exist at high density on vascular endothelium (Yamaji, Sarker et al. 2003, van Diepen, Schlicker et al. 2008, Bukiya A.N., Jackson S. et al. 2014, Gasperi, Evangelista et al. 2014, Pacher, Steffens et al. 2018) and can induce arteritis (Pacher, Steffens et al. 2018) be pro-coagulant and anti-prostacyclin (Wang, Yuan et al. 2011, Murphy, Itchon-Ramos et al. 2018) and has been associated with human vascular aging (Reece A.S., Norman et al. 2016). Moreover it has been linked with gastroschisis in many studies (Reece A. S. and Hulse G.K. 2019, Reece A. S. and Hulse G.K. 2019) which is a congenital defect now considered to have a vasculopathic basis (Hoyme, Higginbottom et al. 1981, Van Allen and Smith 1981, Werler, Mitchell et al. 2009, Lubinsky 2014). These data imply that cannabis could be acting via the vasculopathic pathway outlined in the Introduction. Thus various mechanistic pathways exist to underpin the geospatial and causal epidemiological results reported in this study.

### Absolute and Relative Strengths and Weaknesses of the Study

This study has various strengths including its use of population-derived indices such as ethnicity and median household income and a national birth anomalies dataset. It adjusts for estimates of ETOPFA for which hard data does not exist and considers the impact of such data on the findings described. Our study is the first to apply the analytical techniques of geospatial regression and causal inference to this topic, and the first to our knowledge to investigate the impact of drug exposure on this major teratological outcome in the geospatial context. Study limitations are those related to its ecological design. We did not have access to individual case data and were not able to directly correlate exposure with outcomes. Moreover we did not have access to fine geospatial resolution birth defect data such as was recently published by CDC (Short, Stallings et al. 2019). Uncontrolled confounding is always a possibility in such investigations but seems most unlikely to account for the described association in view of the large e-values calculated. In view of the public health importance of the issue and the numbers of exposed individuals we feel that the area needs further research at both the basic sciences and finer spatial epidemiological level.

### Generalizability

In that the USA is the world’s leading nation on many metrics and provides the best publicly available data on both drug exposure and birth defects, we feel that the present findings are likely to be generalizable to similar contexts in other western nations.

### Implications for Policy and Future Directions

Our interpretation of these results is that cannabis exposure has a strong geospatial link to LRR which is robust to adjustment for socioeconomic and sociodemographic factors and fulfils the criteria for causality. These results are particularly concerning given prior reports of a wide spectrum of cardiovascular, neurological and other anomalies reported as being cannabis-related from various locations including Hawaii, Colorado, Canada, France, Switzerland and Australia (Forrester and Merz 2007, Report of the Queensland Perinatal Maternal and Perinatal Quality Council and Queensland Health 2018, Reece A. S. and Hulse G.K. 2019, Reece A.S. and Hulse G.K. 2019, Reece A. S. and Hulse G.K. 2020). The causal nature of this relationship concerns us greatly. We feel that such results should be part of a program of improved public awareness in relation to the risks associated with the use of diverse cannabinoids and especially far-reaching intergenerational implications.

## Data Availability

Data Availability Statement. Key data including software code in R and a Data Dictionary key has been made available in the Mendeley data repository at this URL: http://dx.doi.org/10.17632/gtk7w24yvs.1.

## Acknowledgements

All authors had full access to all the data in the study and take responsibility for the integrity of the data and the accuracy of the data analysis.

## Contributorship Statement

ASR assembled the data, designed and conducted the analyses, and wrote the first manuscript draft. GKH provided technical and logistic support, co-wrote the paper, assisted with gaining ethical approval, provided advice on manuscript preparation and general guidance to study conduct. ASR had the idea for the article, performed the literature search, wrote the first draft and is the guarantor for the article.

## Supplementary Material - Table of Contents

| **Item** | **Tables** | **Page** |
| --- | --- | --- |
| Supplementary Table 1 | Raw LR Data | 2 |
| Supplementary Table 2 | Cannabis Use Quintiles | 6 |
| Supplementary Table 3 | Median Values Full and Kriged Datasets | 10 |
| Supplementary Table 4 | Introductory Regressions | 11 |
| Supplementary Table 5 | Cannabinoid Regressions | 12 |
| Supplementary Table 6 | Panel Regressions | 13 |
| Supplementary Table 7 | Kriged Data | 15 |
| Supplementary Table 8 | ICBDSR Data Re-Visited | 17 |
| Supplementary Table 9 | WARDA Data on ETOPFA Rates | 21 |
| Supplementary Table 11 | Spatial Regression with International Adjustment Factor for Silent Cases – 63% | 23 |
| Supplementary Table 12 | Two-Step Instrumental Regression by Race | 25 |
| Supplementary Table 13 | Mixed Effects Regression with IPW Weights | 28 |
|  | **Figures** |  |
| Supplementary Figure 1 | Limb Reduction Rate by State – Facetted Plot | 30 |
| Supplementary Figure 2 | Limb Reduction Rate by State – Geo-facetted Plot | 31 |
| Supplementary Figure 3 | Daily Cannabis Use – General and in Pregnancy | 32 |
| Supplementary Figure 4 | LRR- Income Association | 33 |
| Supplementary Figure 5 | LR Non-Live Birth Rate – Scaled with USA Silent Factor, 37% | 34 |
| Supplementary Figure 6 | LRR Over Time by Quintile Corrected for USA Silent Factor | 35 |
| Supplementary Figure 7 | LRR Boxplots by Quintile Corrected for USA Silent Factor | 36 |
| Supplementary Figure 8 | LRR Boxplots by Dichotomized Quintile Corrected for USA Silent Factor | 37 |
| Supplementary Figure 9 | Map-Graphs of LRR Corrected for USA Silent Factor – Raw Data | 38 |
| Supplementary Figure 10 | Map-Graphs of LRR Corrected for USA Silent Factor – Kriged Data | 39 |
| Supplementary Figure 11 | LR Non-Live Birth Rate Over Time – Scaled with International Silent Factor, 63% | 40 |
| Supplementary Figure 12 | LR Non-Live Birth Rate Boxplots – Scaled with International Silent Factor, 63% | 41 |
| Supplementary Figure 13 | LR Non-Live Birth Rate Over Time by Quintiles – Scaled with International Silent Factor | 42 |
| Supplementary Figure 14 | Map-Graphs of LRR Over Time – Adjusted with International Silent Factor – Raw Data | 43 |
| Supplementary<br>Figure 15 | Map-Graphs of LRR Over Time – Adjusted with International Silent Factor – Kriged Data | 44 |
| Supplementary<br>Figure 16 | Drug Exposure Plots for Hidden Factors of 0%, 37% and 63% | 45 |
| Supplementary<br>Figure 17 | Ethnic Composition Plots for Hidden Factors of 0%, 37% and 63% | 46 |
| Supplementary<br>Figure 18 | Effects of Registry Ascertainment and Case Finding Practices | 47 |

**Supplementary Table 1.:**
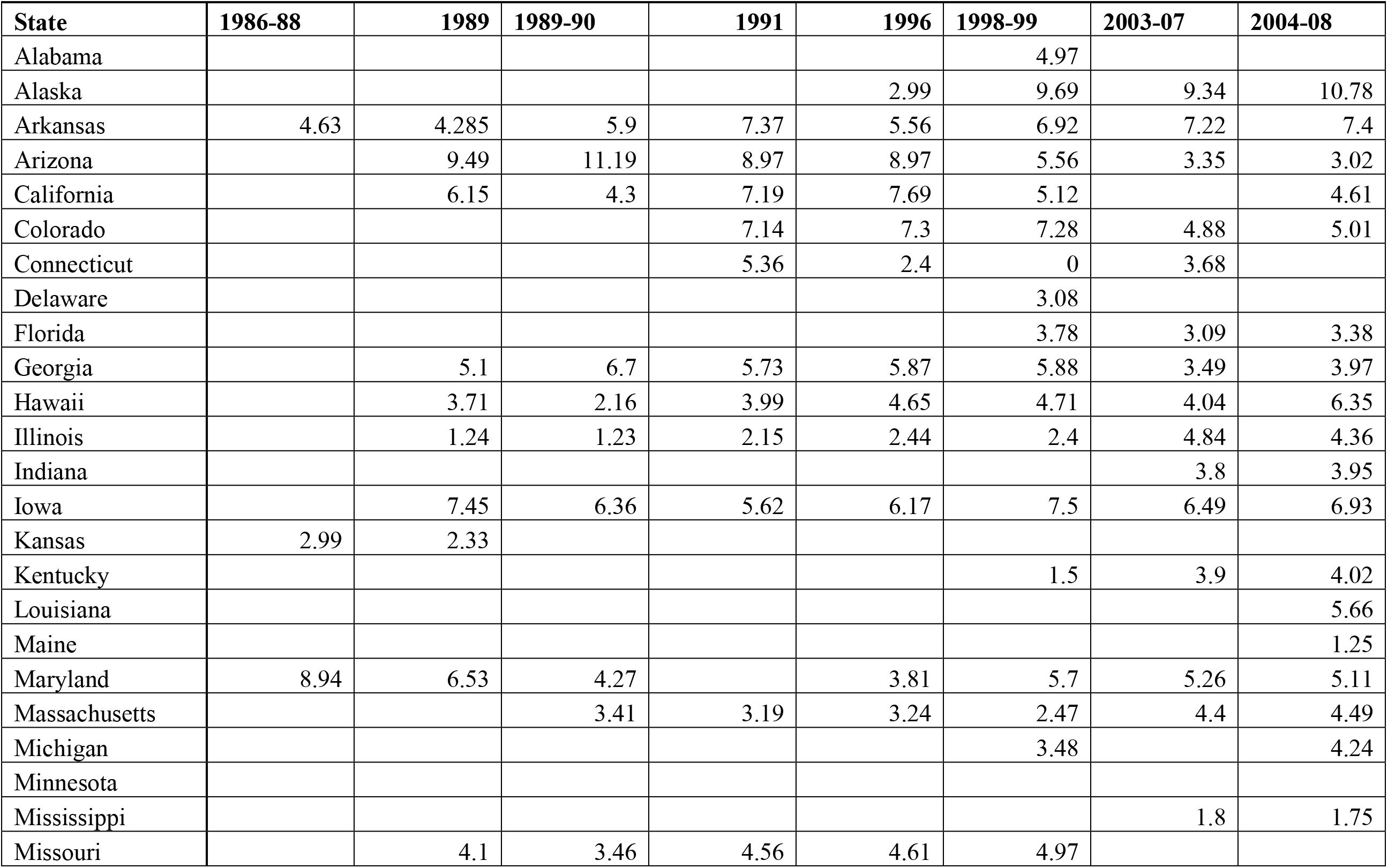
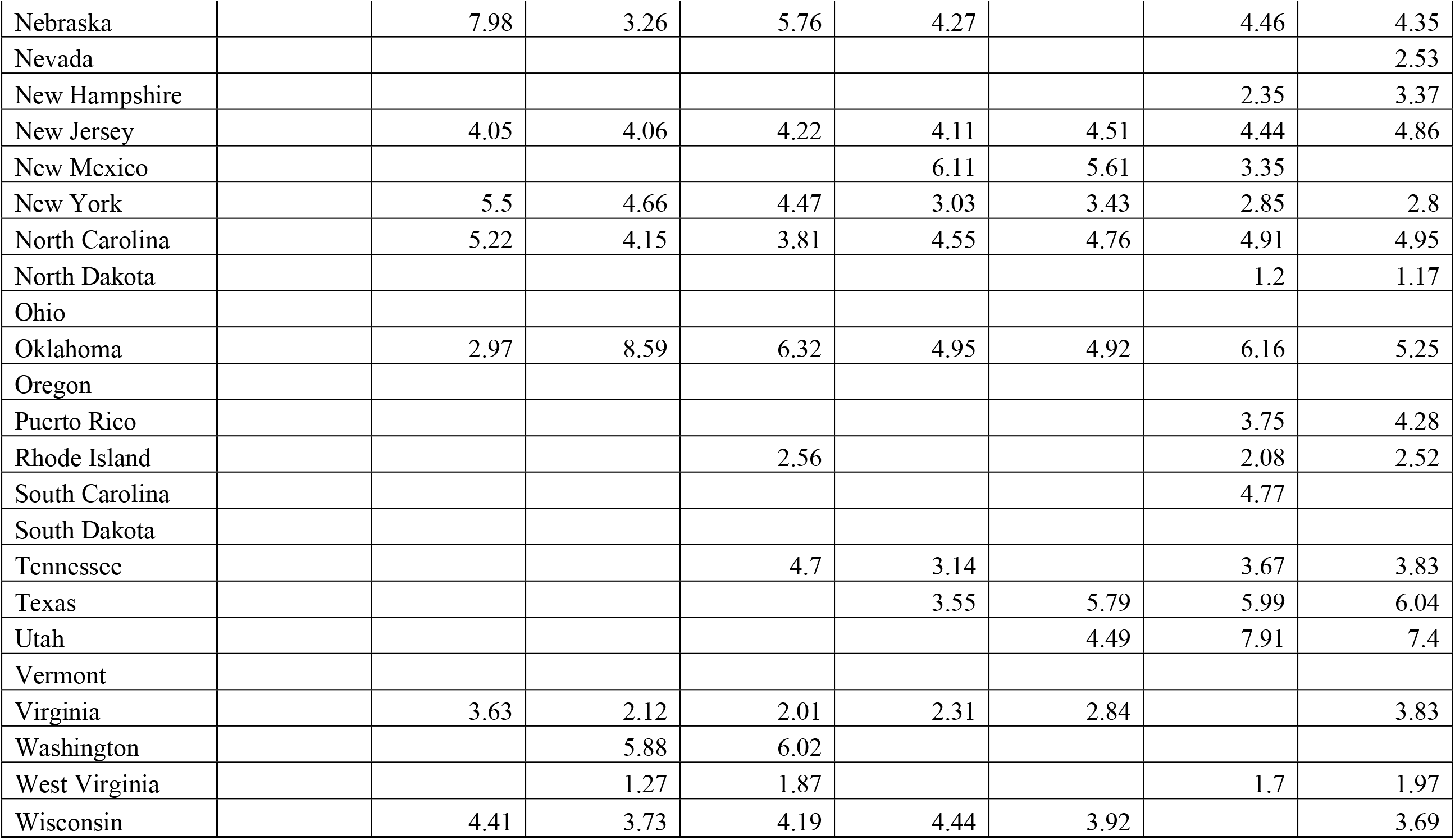
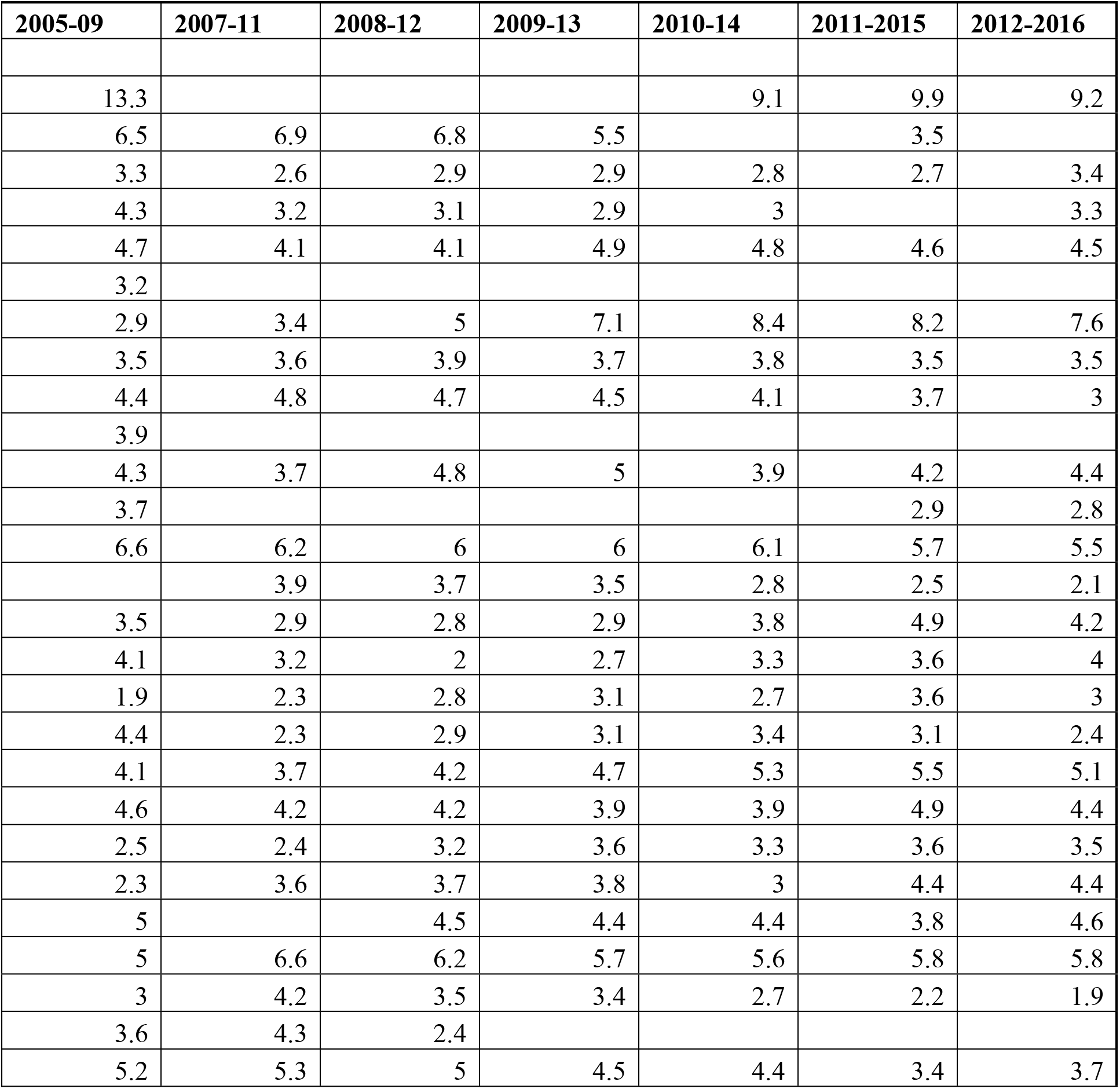
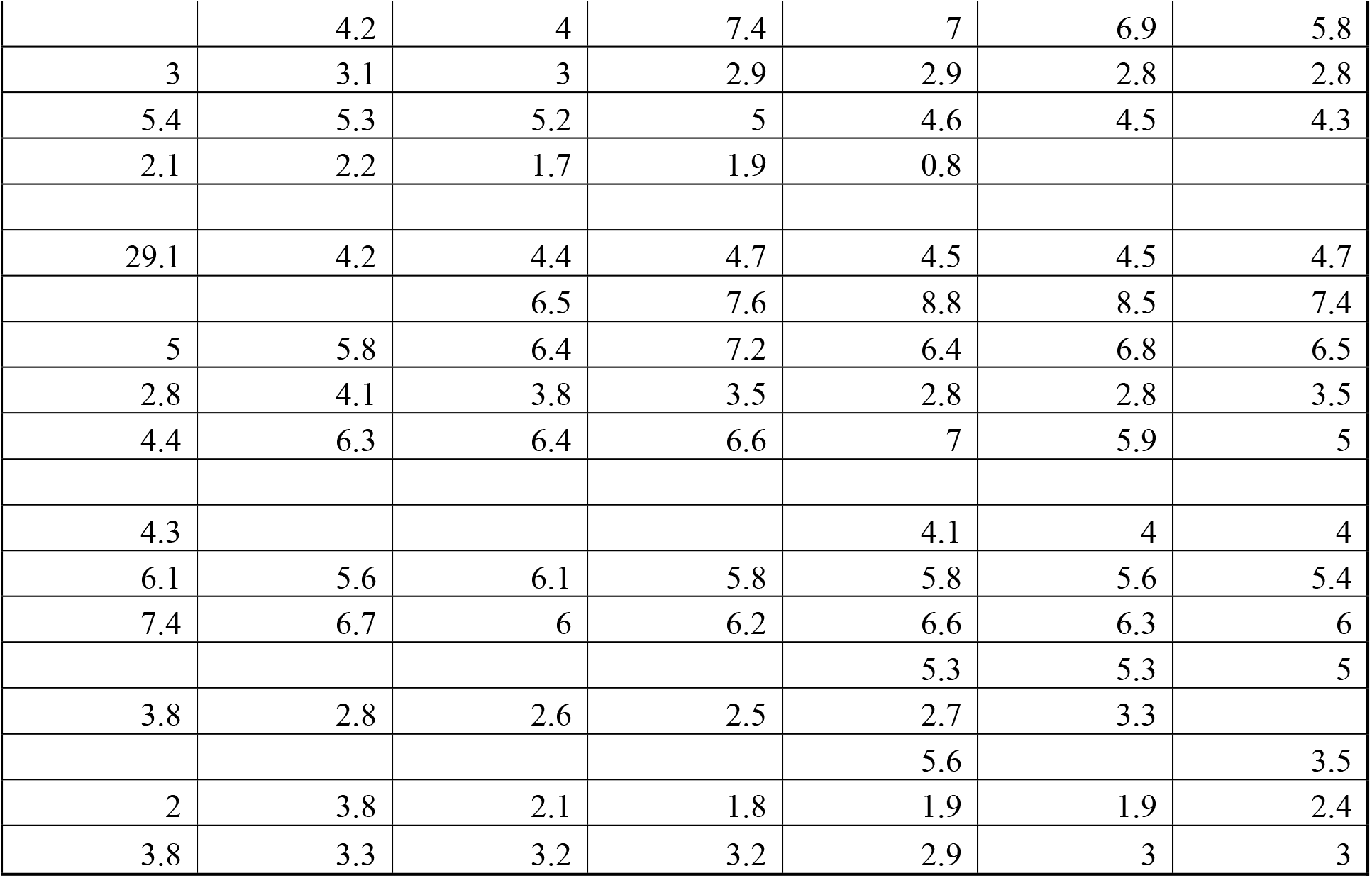
Raw Limb Reduction Data.

**Supplementary Table 2.:** Cannabis Use Quintiles.

| State | Year | mrjmon | QuinCI | Quin | QuinCN |
| --- | --- | --- | --- | --- | --- |
| Colorado | 2015 | 0.171216 | (0.146,0.171] | Quintile 5 | (0.113,0.171] |
| Vermont | 2015 | 0.150801 | (0.146,0.171] | Quintile 5 | (0.113,0.171] |
| Alaska | 2015 | 0.147986 | (0.146,0.171] | Quintile 5 | (0.113,0.171] |
| Maine | 2015 | 0.139832 | (0.122,0.146] | Quintile 4 | (0.113,0.171] |
| Rhode Island | 2015 | 0.132805 | (0.122,0.146] | Quintile 4 | (0.113,0.171] |
| Oregon | 2015 | 0.130385 | (0.122,0.146] | Quintile 4 | (0.113,0.171] |
| New Hampshire | 2015 | 0.123054 | (0.122,0.146] | Quintile 4 | (0.113,0.171] |
| District of Columbia | 2015 | 0.121021 | (0.0968,0.122] | Quintile 3 | (0.113,0.171] |
| Massachusetts | 2015 | 0.119083 | (0.0968,0.122] | Quintile 3 | (0.113,0.171] |
| Washington | 2015 | 0.114177 | (0.0968,0.122] | Quintile 3 | (0.113,0.171] |
| Montana | 2015 | 0.113184 | (0.0968,0.122] | Quintile 3 | (0.0817,0.113] |
| New Mexico | 2015 | 0.103081 | (0.0968,0.122] | Quintile 3 | (0.0817,0.113] |
| Michigan | 2015 | 0.102894 | (0.0968,0.122] | Quintile 3 | (0.0817,0.113] |
| California | 2015 | 0.098151 | (0.0968,0.122] | Quintile 3 | (0.0817,0.113] |
| New York | 2015 | 0.097454 | (0.0968,0.122] | Quintile 3 | (0.0817,0.113] |
| Connecticut | 2015 | 0.097221 | (0.0968,0.122] | Quintile 3 | (0.0817,0.113] |
| Maryland | 2015 | 0.096645 | (0.072,0.0968] | Quintile 2 | (0.0817,0.113] |
| Indiana | 2015 | 0.087862 | (0.072,0.0968] | Quintile 2 | (0.0817,0.113] |
| Arizona | 2015 | 0.085918 | (0.072,0.0968] | Quintile 2 | (0.0817,0.113] |
| South Carolina | 2015 | 0.082666 | (0.072,0.0968] | Quintile 2 | (0.0817,0.113] |
| Missouri | 2015 | 0.081698 | (0.072,0.0968] | Quintile 2 | (0.0743,0.0817] |
| Hawaii | 2015 | 0.081108 | (0.072,0.0968] | Quintile 2 | (0.0743,0.0817] |
| Delaware | 2015 | 0.081095 | (0.072,0.0968] | Quintile 2 | (0.0743,0.0817] |
| Illinois | 2015 | 0.080563 | (0.072,0.0968] | Quintile 2 | (0.0743,0.0817] |
| Georgia | 2015 | 0.078868 | (0.072,0.0968] | Quintile 2 | (0.0743,0.0817] |
| Florida | 2015 | 0.078732 | (0.072,0.0968] | Quintile 2 | (0.0743,0.0817] |
| Nevada | 2015 | 0.078481 | (0.072,0.0968] | Quintile 2 | (0.0743,0.0817] |
| Ohio | 2015 | 0.078167 | (0.072,0.0968] | Quintile 2 | (0.0743,0.0817] |
| Pennsylvania | 2015 | 0.078005 | (0.072,0.0968] | Quintile 2 | (0.0743,0.0817] |
| Minnesota | 2015 | 0.07738 | (0.072,0.0968] | Quintile 2 | (0.0743,0.0817] |
| North Carolina | 2015 | 0.074308 | (0.072,0.0968] | Quintile 2 | (0.064,0.0743] |
| Kansas | 2015 | 0.073762 | (0.072,0.0968] | Quintile 2 | (0.064,0.0743] |
| Arkansas | 2015 | 0.072504 | (0.072,0.0968] | Quintile 2 | (0.064,0.0743] |
| Kentucky | 2015 | 0.071327 | [0.0472,0.072] | Quintile 1 | (0.064,0.0743] |
| New Jersey | 2015 | 0.070252 | [0.0472,0.072] | Quintile 1 | (0.064,0.0743] |
| West Virginia | 2015 | 0.069969 | [0.0472,0.072] | Quintile 1 | (0.064,0.0743] |
| South Dakota | 2015 | 0.069882 | [0.0472,0.072] | Quintile 1 | (0.064,0.0743] |
| Wisconsin | 2015 | 0.068192 | [0.0472,0.072] | Quintile 1 | (0.064,0.0743] |
| Virginia | 2015 | 0.067445 | [0.0472,0.072] | Quintile 1 | (0.064,0.0743] |
| Louisiana | 2015 | 0.064257 | [0.0472,0.072] | Quintile 1 | (0.064,0.0743] |
| Nebraska | 2015 | 0.063974 | [0.0472,0.072] | Quintile 1 | [0.0472,0.064] |
| Tennessee | 2015 | 0.063807 | [0.0472,0.072] | Quintile 1 | [0.0472,0.064] |
| Oklahoma | 2015 | 0.063326 | [0.0472,0.072] | Quintile 1 | [0.0472,0.064] |
| Idaho | 2015 | 0.063242 | [0.0472,0.072] | Quintile 1 | [0.0472,0.064] |
| Wyoming | 2015 | 0.062962 | [0.0472,0.072] | Quintile 1 | [0.0472,0.064] |
| Texas | 2015 | 0.060034 | [0.0472,0.072] | Quintile 1 | [0.0472,0.064] |
| North Dakota | 2015 | 0.059227 | [0.0472,0.072] | Quintile 1 | [0.0472,0.064] |
| Alabama | 2015 | 0.053685 | [0.0472,0.072] | Quintile 1 | [0.0472,0.064] |
| Mississippi | 2015 | 0.053619 | [0.0472,0.072] | Quintile 1 | [0.0472,0.064] |
| Utah | 2015 | 0.051131 | [0.0472,0.072] | Quintile 1 | [0.0472,0.064] |
| Iowa | 2015 | 0.047242 | [0.0472,0.072] | Quintile 1 | [0.0472,0.064] |

**Alphabetical Listing of Cannabis Use Quintiles**
| State | Year | mrjmon | QuinCI | Quin | QuinCN |
| --- | --- | --- | --- | --- | --- |
| Alabama | 2015 | 0.053685 | [0.0472,0.072] | Quintile 1 | [0.0472,0.064] |
| Alaska | 2015 | 0.147986 | (0.146,0.171] | Quintile 5 | (0.113,0.171] |
| Arizona | 2015 | 0.085918 | (0.072,0.0968] | Quintile 2 | (0.0817,0.113] |
| Arkansas | 2015 | 0.072504 | (0.072,0.0968] | Quintile 2 | (0.064,0.0743] |
| California | 2015 | 0.098151 | (0.0968,0.122] | Quintile 3 | (0.0817,0.113] |
| Colorado | 2015 | 0.171216 | (0.146,0.171] | Quintile 5 | (0.113,0.171] |
| Connecticut | 2015 | 0.097221 | (0.0968,0.122] | Quintile 3 | (0.0817,0.113] |
| Delaware | 2015 | 0.081095 | (0.072,0.0968] | Quintile 2 | (0.0743,0.0817] |
| District of Columbia | 2015 | 0.121021 | (0.0968,0.122] | Quintile 3 | (0.113,0.171] |
| Florida | 2015 | 0.078732 | (0.072,0.0968] | Quintile 2 | (0.0743,0.0817] |
| Georgia | 2015 | 0.078868 | (0.072,0.0968] | Quintile 2 | (0.0743,0.0817] |
| Hawaii | 2015 | 0.081108 | (0.072,0.0968] | Quintile 2 | (0.0743,0.0817] |
| Idaho | 2015 | 0.063242 | [0.0472,0.072] | Quintile 1 | [0.0472,0.064] |
| Illinois | 2015 | 0.080563 | (0.072,0.0968] | Quintile 2 | (0.0743,0.0817] |
| Indiana | 2015 | 0.087862 | (0.072,0.0968] | Quintile 2 | (0.0817,0.113] |
| Iowa | 2015 | 0.047242 | [0.0472,0.072] | Quintile 1 | [0.0472,0.064] |
| Kansas | 2015 | 0.073762 | (0.072,0.0968] | Quintile 2 | (0.064,0.0743] |
| Kentucky | 2015 | 0.071327 | [0.0472,0.072] | Quintile 1 | (0.064,0.0743] |
| Louisiana | 2015 | 0.064257 | [0.0472,0.072] | Quintile 1 | (0.064,0.0743] |
| Maine | 2015 | 0.139832 | (0.122,0.146] | Quintile 4 | (0.113,0.171] |
| Maryland | 2015 | 0.096645 | (0.072,0.0968] | Quintile 2 | (0.0817,0.113] |
| Massachusetts | 2015 | 0.119083 | (0.0968,0.122] | Quintile 3 | (0.113,0.171] |
| Michigan | 2015 | 0.102894 | (0.0968,0.122] | Quintile 3 | (0.0817,0.113] |
| Minnesota | 2015 | 0.07738 | (0.072,0.0968] | Quintile 2 | (0.0743,0.0817] |
| Mississippi | 2015 | 0.053619 | [0.0472,0.072] | Quintile 1 | [0.0472,0.064] |
| Missouri | 2015 | 0.081698 | (0.072,0.0968] | Quintile 2 | (0.0743,0.0817] |
| Montana | 2015 | 0.113184 | (0.0968,0.122] | Quintile 3 | (0.0817,0.113] |
| Nebraska | 2015 | 0.063974 | [0.0472,0.072] | Quintile 1 | [0.0472,0.064] |
| Nevada | 2015 | 0.078481 | (0.072,0.0968] | Quintile 2 | (0.0743,0.0817] |
| New Hampshire | 2015 | 0.123054 | (0.122,0.146] | Quintile 4 | (0.113,0.171] |
| New Jersey | 2015 | 0.070252 | [0.0472,0.072] | Quintile 1 | (0.064,0.0743] |
| New Mexico | 2015 | 0.103081 | (0.0968,0.122] | Quintile 3 | (0.0817,0.113] |
| New York | 2015 | 0.097454 | (0.0968,0.122] | Quintile 3 | (0.0817,0.113] |
| North Carolina | 2015 | 0.074308 | (0.072,0.0968] | Quintile 2 | (0.064,0.0743] |
| North Dakota | 2015 | 0.059227 | [0.0472,0.072] | Quintile 1 | [0.0472,0.064] |
| Ohio | 2015 | 0.078167 | (0.072,0.0968] | Quintile 2 | (0.0743,0.0817] |
| Oklahoma | 2015 | 0.063326 | [0.0472,0.072] | Quintile 1 | [0.0472,0.064] |
| Oregon | 2015 | 0.130385 | (0.122,0.146] | Quintile 4 | (0.113,0.171] |
| Pennsylvania | 2015 | 0.078005 | (0.072,0.0968] | Quintile 2 | (0.0743,0.0817] |
| Rhode Island | 2015 | 0.132805 | (0.122,0.146] | Quintile 4 | (0.113,0.171] |
| South Carolina | 2015 | 0.082666 | (0.072,0.0968] | Quintile 2 | (0.0817,0.113] |
| South Dakota | 2015 | 0.069882 | [0.0472,0.072] | Quintile 1 | (0.064,0.0743] |
| Tennessee | 2015 | 0.063807 | [0.0472,0.072] | Quintile 1 | [0.0472,0.064] |
| Texas | 2015 | 0.060034 | [0.0472,0.072] | Quintile 1 | [0.0472,0.064] |
| Utah | 2015 | 0.051131 | [0.0472,0.072] | Quintile 1 | [0.0472,0.064] |
| Vermont | 2015 | 0.150801 | (0.146,0.171] | Quintile 5 | (0.113,0.171] |
| Virginia | 2015 | 0.067445 | [0.0472,0.072] | Quintile 1 | (0.064,0.0743] |
| Washington | 2015 | 0.114177 | (0.0968,0.122] | Quintile 3 | (0.113,0.171] |
| West Virginia | 2015 | 0.069969 | [0.0472,0.072] | Quintile 1 | (0.064,0.0743] |
| Wisconsin | 2015 | 0.068192 | [0.0472,0.072] | Quintile 1 | (0.064,0.0743] |
| Wyoming | 2015 | 0.062962 | [0.0472,0.072] | Quintile 1 | [0.0472,0.064] |

**Supplementary Table 3.:** Median Values Full and Kriged Datasets.

| Quintile | Full Dataset 1988-2016 | Kriged Dataset 2003-2016 |
| --- | --- | --- |
| | Median ± S.E.M. | Median ± S.E.M. |
| Quintile 1 | 6.06 (5.61, 6.51) | 9.2 (8.49, 9.91) |
| Quintile 2 | 5.93 (5.5, 6.36) | 9.41 (8.68, 10.14) |
| Quintile 3 | 6.03 (5.5, 6.56) | 8.89 (7.91, 9.87) |
| Quintile 4 | 5.55 (4.26, 6.84) | 8.67 (6.51, 10.83) |
| Quintile 5 | 10.61 (8.41, 12.81) | 16.36 (12.95, 19.77) |

**Supplementary Table 4.:** Introductory Regressions.

| Parameter | Parameter |  | Model Parameters |  |  |  |
| --- | --- | --- | --- | --- | --- | --- |
|  | Estimate (C.I.) | Pr(>\|t\|) | R-Squared | F | dF | P |
| *lm(LR_Rate ~ Year)* |  |  |  |  |  |  |
| Year | -0.02 (-0.0396, -0.0004) | 0.0513 | 0.0065 | 3.821 | 1,433 | 0.0513 |
| *lm(LR_Rate ~ Quintile)* |  |  |  |  |  |  |
| Quintile 5 | 0.43 (0.27, 0.59) | 2.9E-08 | 0.0759 | 9.915 | 4,430 | 1.1E-07 |
| *lm(LR_Rate ~ Dichotomized_Quintiles)* |  |  |  |  |  |  |
| Highest Quintile | 0.42 (0.28, 0.56) | 2.2E-08 | 0.0677 | 32.49 | 1,433 | 2.2E-08 |
| *lm(LR_Rate ~ Cannabis_Use)* |  |  |  |  |  |  |
| Cannabis_Monthly | 8.51 (0.06, 16.96) | 0.0492 | 0.0091 | 3.898 | 1,314 | 0.0492 |
| *lm(LR_Rate ~ Cannabis_Use \* THC_Potency)* |  |  |  |  |  |  |
| Cannabis: THC | 10.64 (3.7, 17.58) | 0.0029 | 0.0310 | 4.356 | 3,312 | 0.0050 |
| THC_Concentration | -0.7 (-1.17, -0.23) | 0.0037 |  |  |  |  |
| Cannabis_Monthly | -109.27 (-186.69, -31.85) | 0.0060 |  |  |  |  |

**Supplementary Table 5.:**
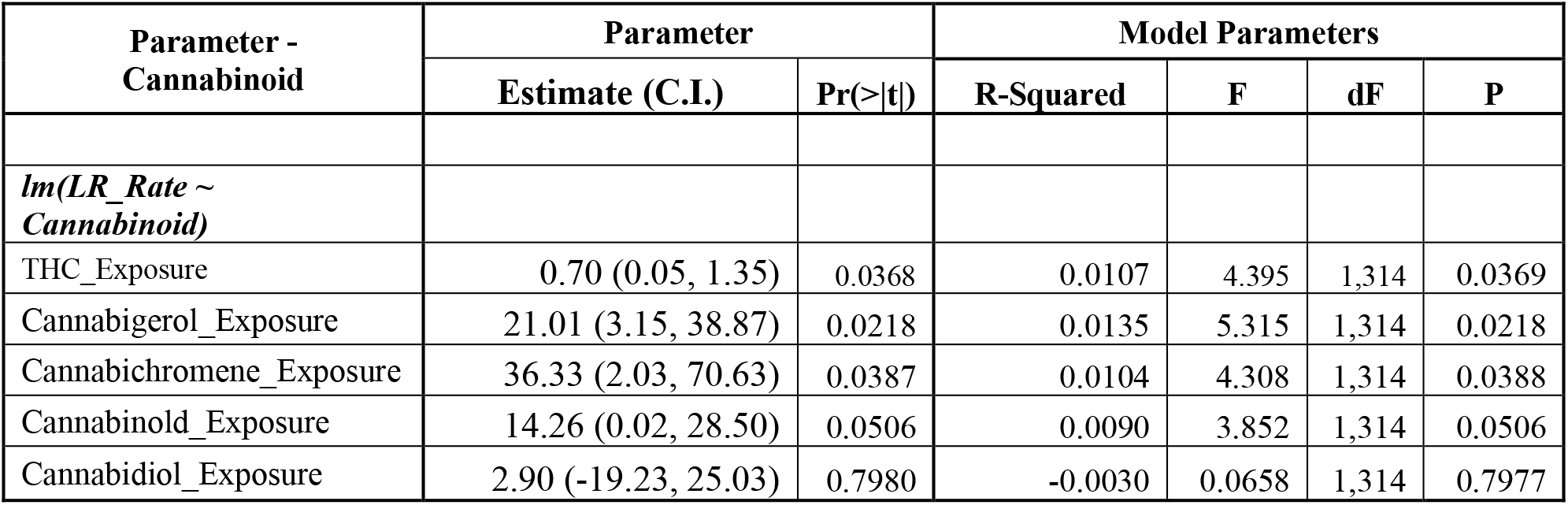
Cannabinoid Regressions.

**Supplementary Table 6.:**
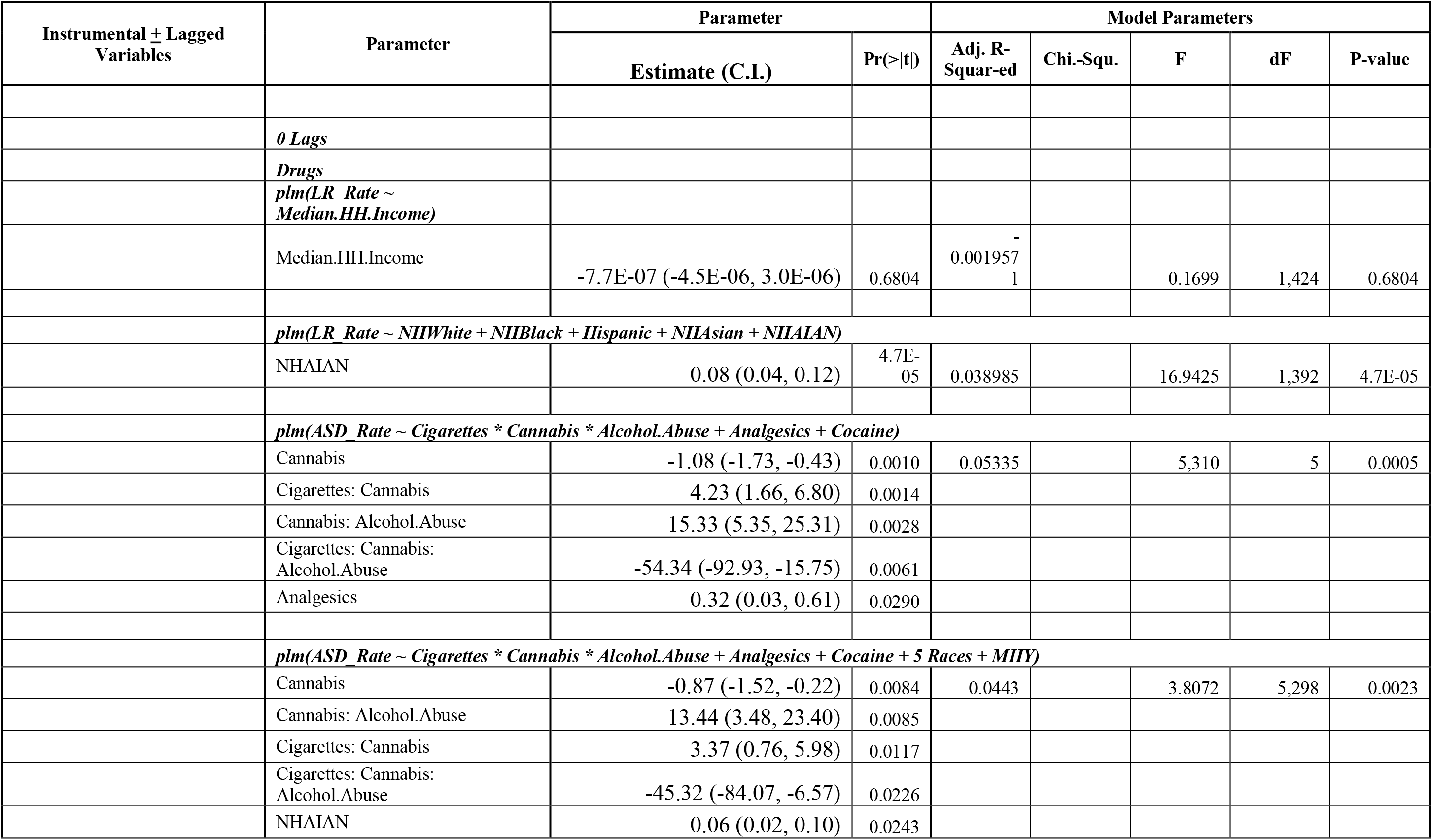
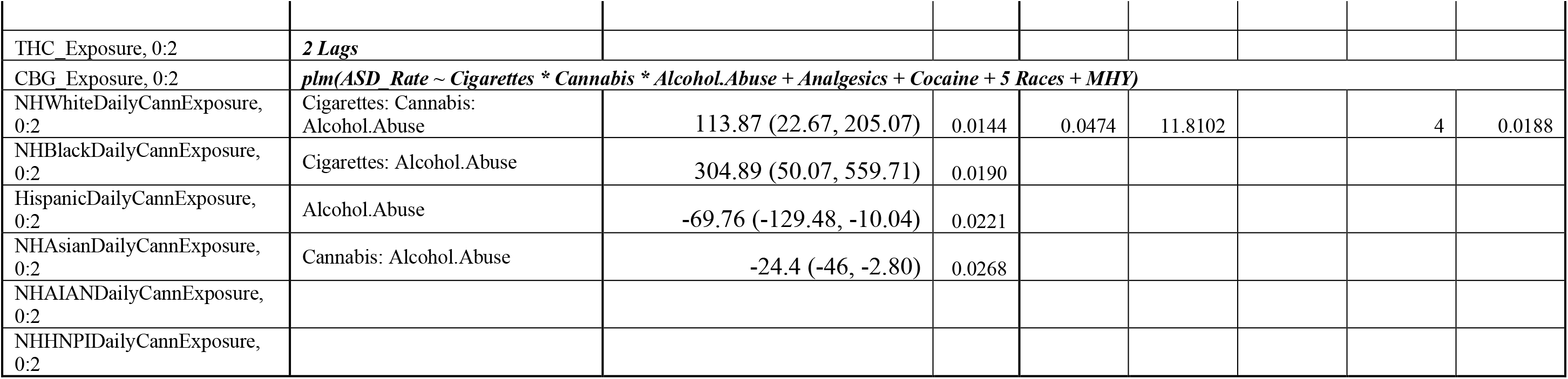
Panel Regressions.

**Supplementary Table 7.:**
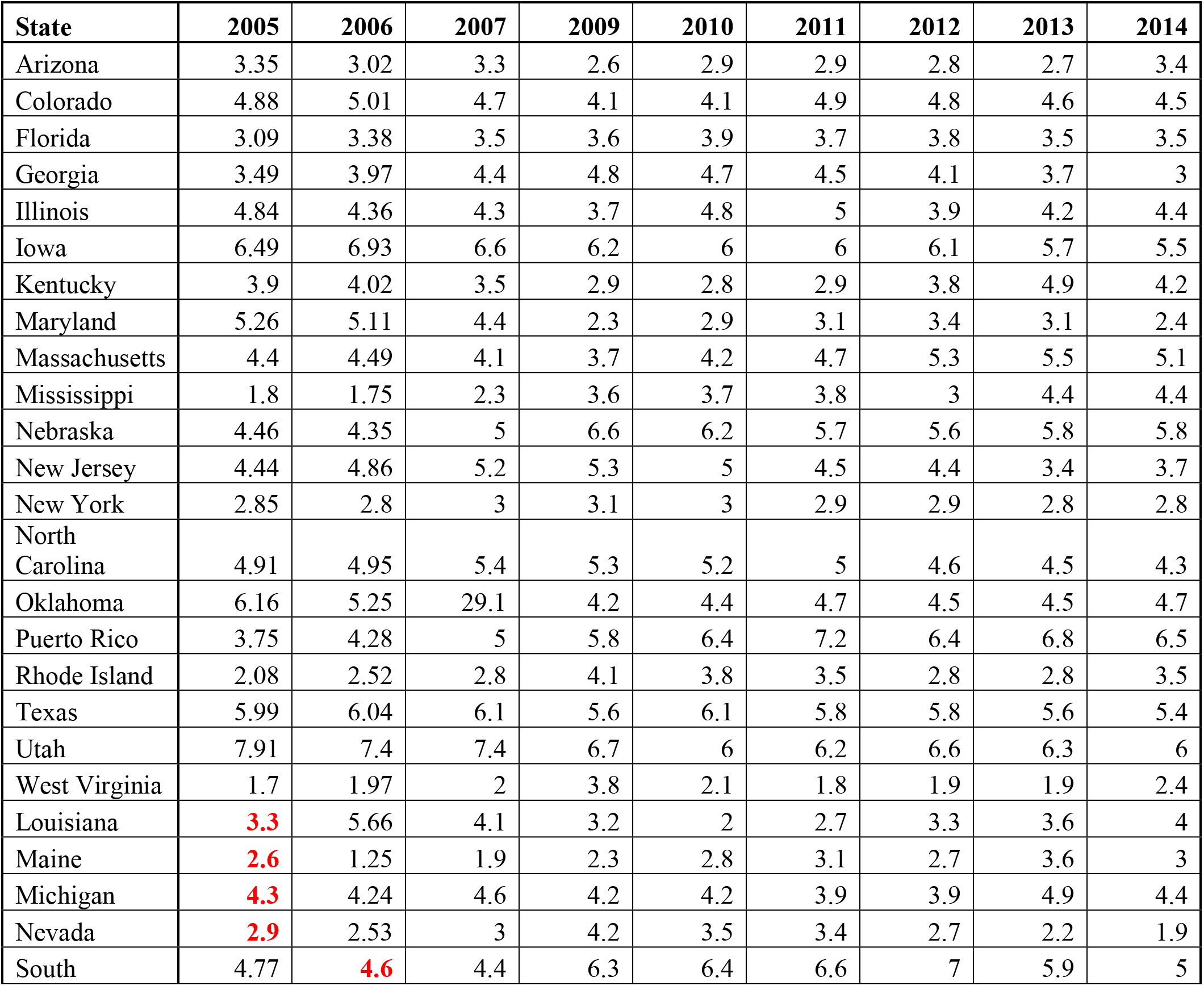
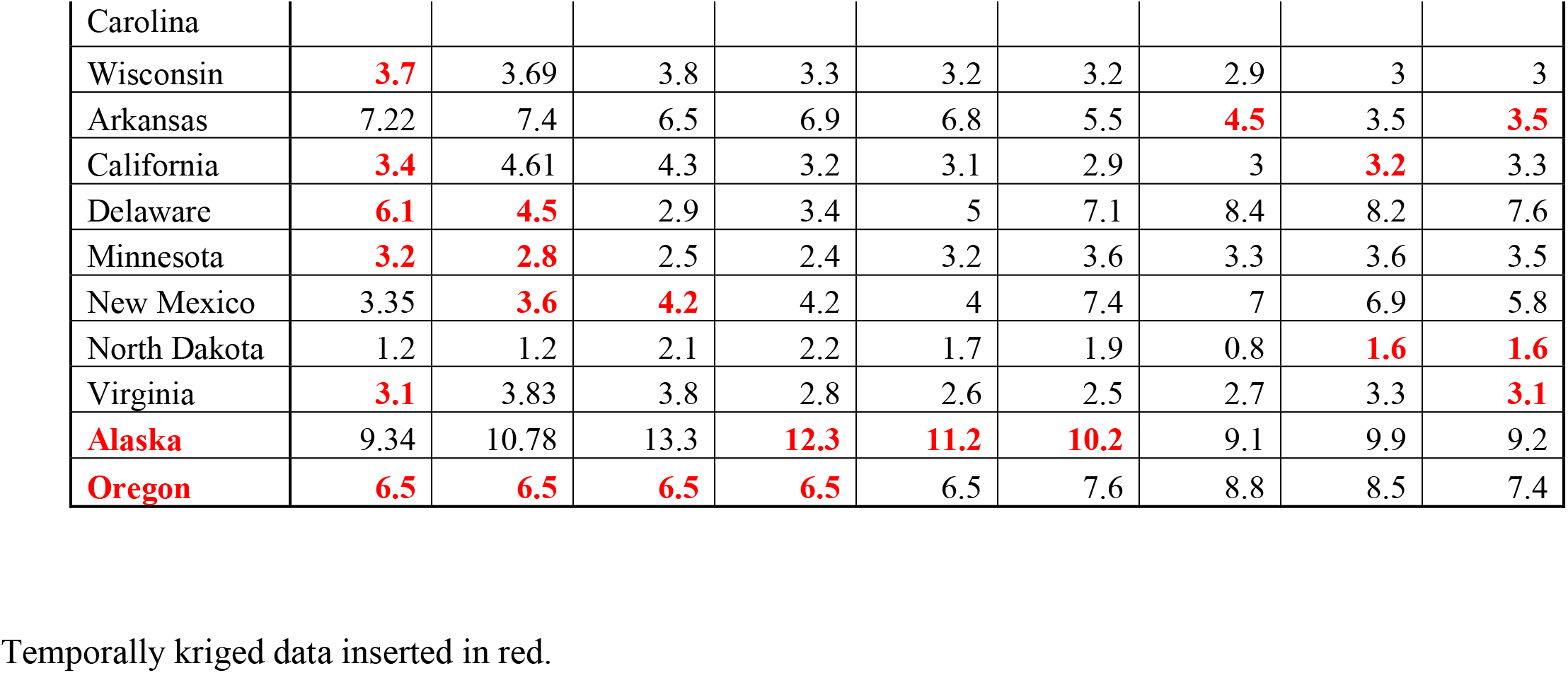
Kriged Data.

**Supplementary Table 8.:**
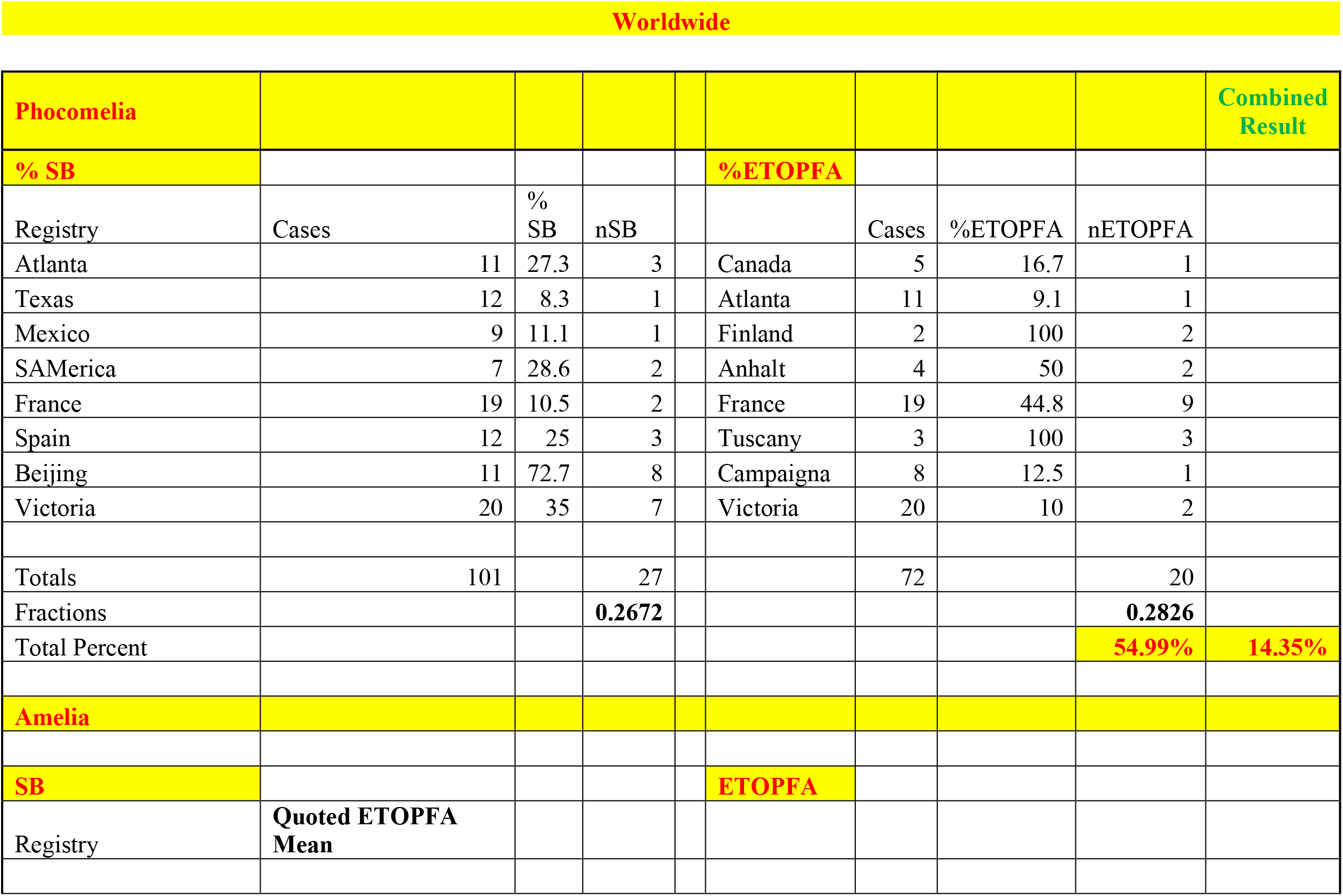
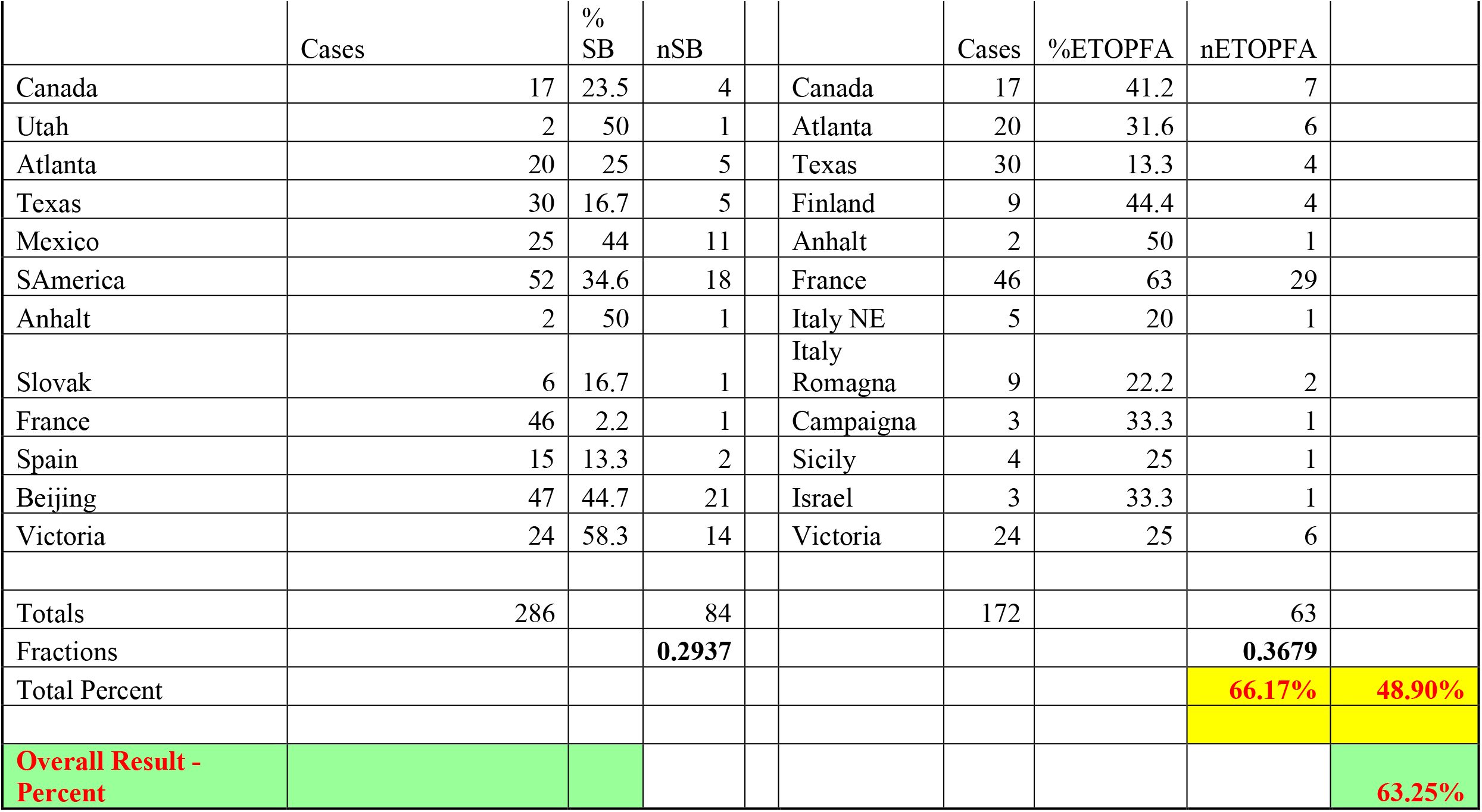
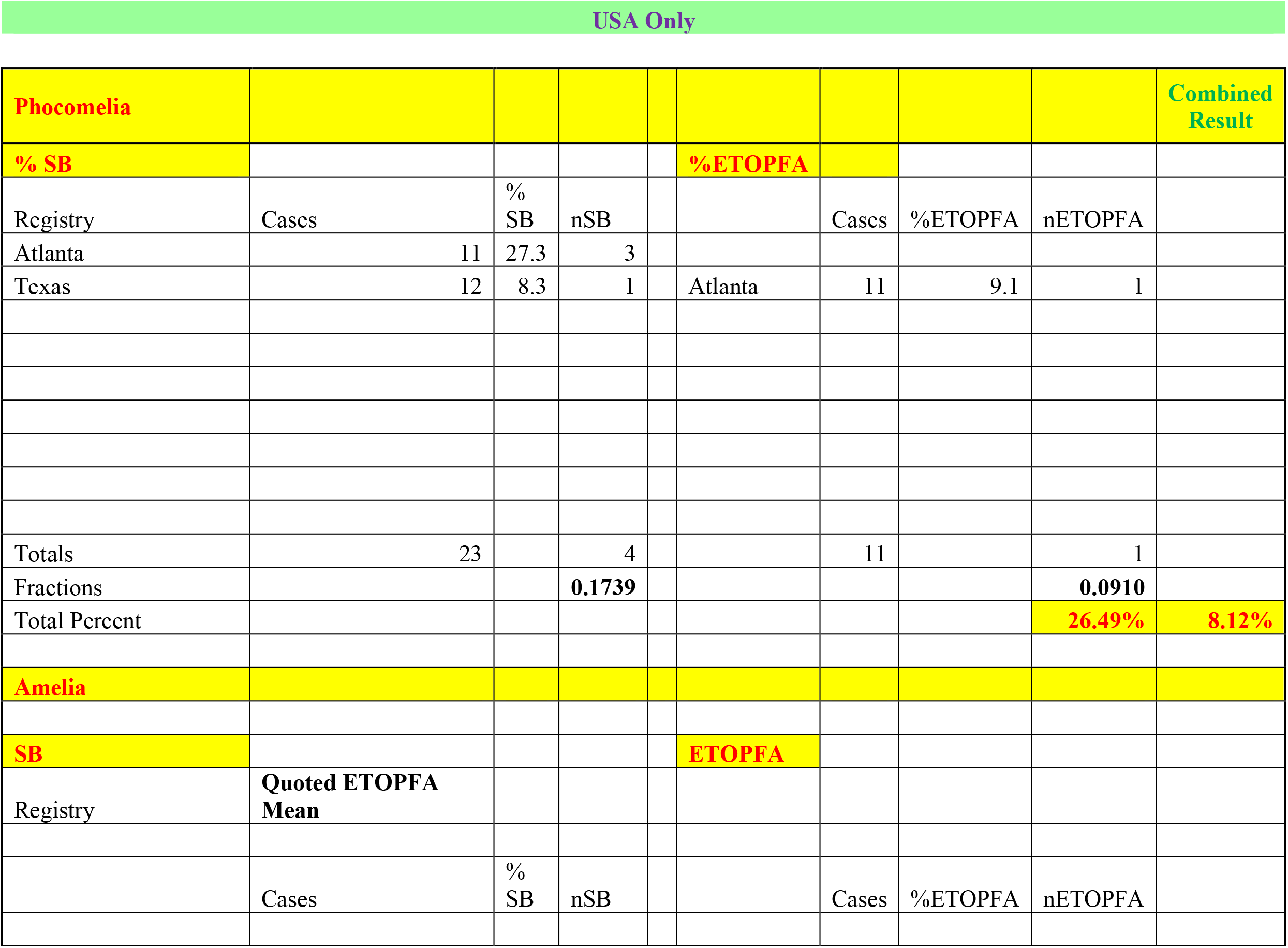
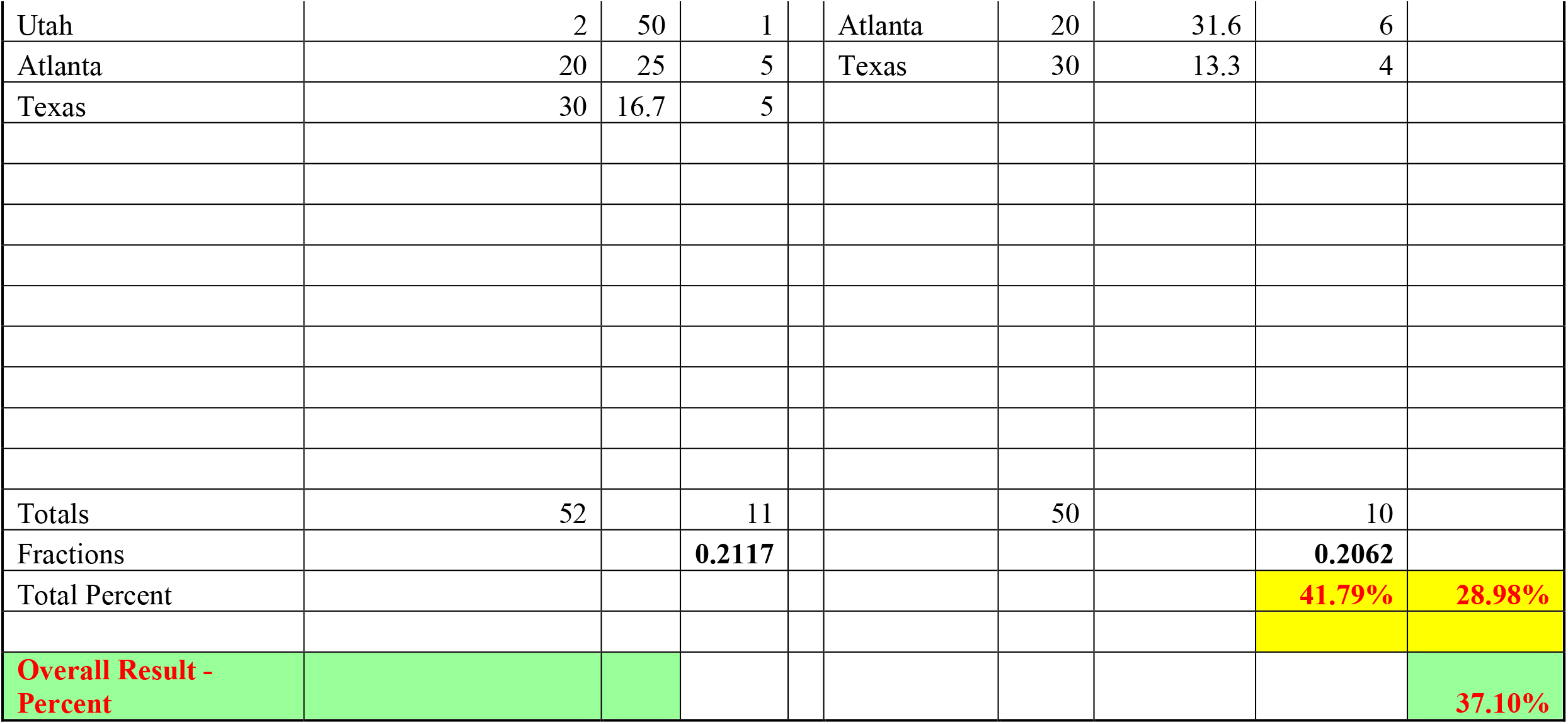
ICBDSR Data Re-Visited.

**Supplementary Table 9.:** WARDA Data on ETOPFA Rates.

| **Year** | **Live.Births** | **ETOPFA** | **Total.T21** | **WA.T21.TOP.Rate** | **TopRt** | **FMaxTR** |
| --- | --- | --- | --- | --- | --- | --- |
| 1980 | 1.0987 | 0.0000 | 1.0987 | 0.0000 | 0.0000 | 0.0000 |
| 1981 | 0.8537 | 0.1254 | 0.9791 | 0.1275 | 0.1275 | 0.1645 |
| 1982 | 1.1136 | 0.1184 | 1.2320 | 0.0941 | 0.0941 | 0.1215 |
| 1983 | 1.0850 | 0.1115 | 1.1965 | 0.0923 | 0.0923 | 0.1191 |
| 1984 | 0.9987 | 0.1910 | 1.1897 | 0.1581 | 0.1581 | 0.2041 |
| 1985 | 1.1504 | 0.3283 | 1.4787 | 0.2183 | 0.2183 | 0.2818 |
| 1986 | 1.1651 | 0.4079 | 1.5729 | 0.2496 | 0.2496 | 0.3222 |
| 1987 | 1.0283 | 0.3144 | 1.3427 | 0.2131 | 0.2131 | 0.2750 |
| 1988 | 1.0863 | 0.5670 | 1.6533 | 0.3323 | 0.3323 | 0.4289 |
| 1989 | 0.9783 | 0.2932 | 1.2716 | 0.2276 | 0.2276 | 0.2938 |
| 1990 | 1.1156 | 0.3368 | 1.4523 | 0.2135 | 0.2135 | 0.2756 |
| 1991 | 1.0437 | 0.4235 | 1.4673 | 0.2722 | 0.2722 | 0.3514 |
| 1992 | 1.2603 | 0.5247 | 1.7851 | 0.2834 | 0.2834 | 0.3658 |
| 1993 | 1.4625 | 0.5250 | 1.9875 | 0.2521 | 0.2521 | 0.3253 |
| 1994 | 0.6695 | 0.8570 | 1.5265 | 0.5557 | 0.5557 | 0.7173 |
| 1995 | 1.0231 | 1.1168 | 2.1399 | 0.5080 | 0.5080 | 0.6557 |
| 1996 | 1.0882 | 0.8358 | 1.9241 | 0.4141 | 0.4141 | 0.5345 |
| 1997 | 0.8433 | 1.0092 | 1.8525 | 0.5322 | 0.5322 | 0.6869 |
| 1998 | 0.7498 | 0.9517 | 1.7015 | 0.5301 | 0.5301 | 0.6842 |
| 1999 | 0.8149 | 1.0241 | 1.8390 | 0.5399 | 0.5399 | 0.6969 |
| 2000 | 1.2767 | 1.7022 | 2.9789 | 0.5606 | 0.5606 | 0.7236 |
| 2001 | 0.7289 | 1.3779 | 2.1068 | 0.6454 | 0.6454 | 0.8330 |
| 2002 | 1.1402 | 1.5080 | 2.6482 | 0.5430 | 0.5430 | 0.7008 |
| 2003 | 0.8232 | 1.4073 | 2.2304 | 0.6058 | 0.6058 | 0.7819 |
| 2004 | 1.0974 | 1.7032 | 2.8006 | 0.5800 | 0.5800 | 0.7487 |
| 2005 | 1.0400 | 1.8621 | 2.9021 | 0.6341 | 0.6341 | 0.8184 |
| 2006 | 0.8383 | 1.6893 | 2.5276 | 0.6539 | 0.6539 | 0.8440 |
| 2007 | 1.0044 | 1.6823 | 2.6868 | 0.6008 | 0.6008 | 0.7755 |
| 2008 | 0.7523 | 1.5095 | 2.2618 | 0.6224 | 0.6224 | 0.8033 |
| 2009 | 0.9761 | 1.9857 | 2.9618 | 0.6489 | 0.6489 | 0.8375 |
| 2010 | 0.9582 | 2.2352 | 3.1935 | 0.6997 | 0.6997 | 0.9031 |
| 2011 | 0.6527 | 2.2291 | 2.8819 | 0.7815 | 0.7815 | 1.0087 |
| 2012 | 0.7383 | 2.1802 | 2.9185 | 0.7531 | 0.7531 | 0.9720 |
| 2013 | 0.7200 | 2.0825 | 2.8024 | 0.7461 | 0.7461 | 0.9630 |
| 2014 | 0.5122 | 1.8075 | 2.3198 | 0.7748 | 0.7748 | 1.0000 |
| 2015 | 0.5122 | 1.8075 | 2.3198 | 0.7748 | 0.7748 | 1.0000 |

**Supplementary Table 10.:**
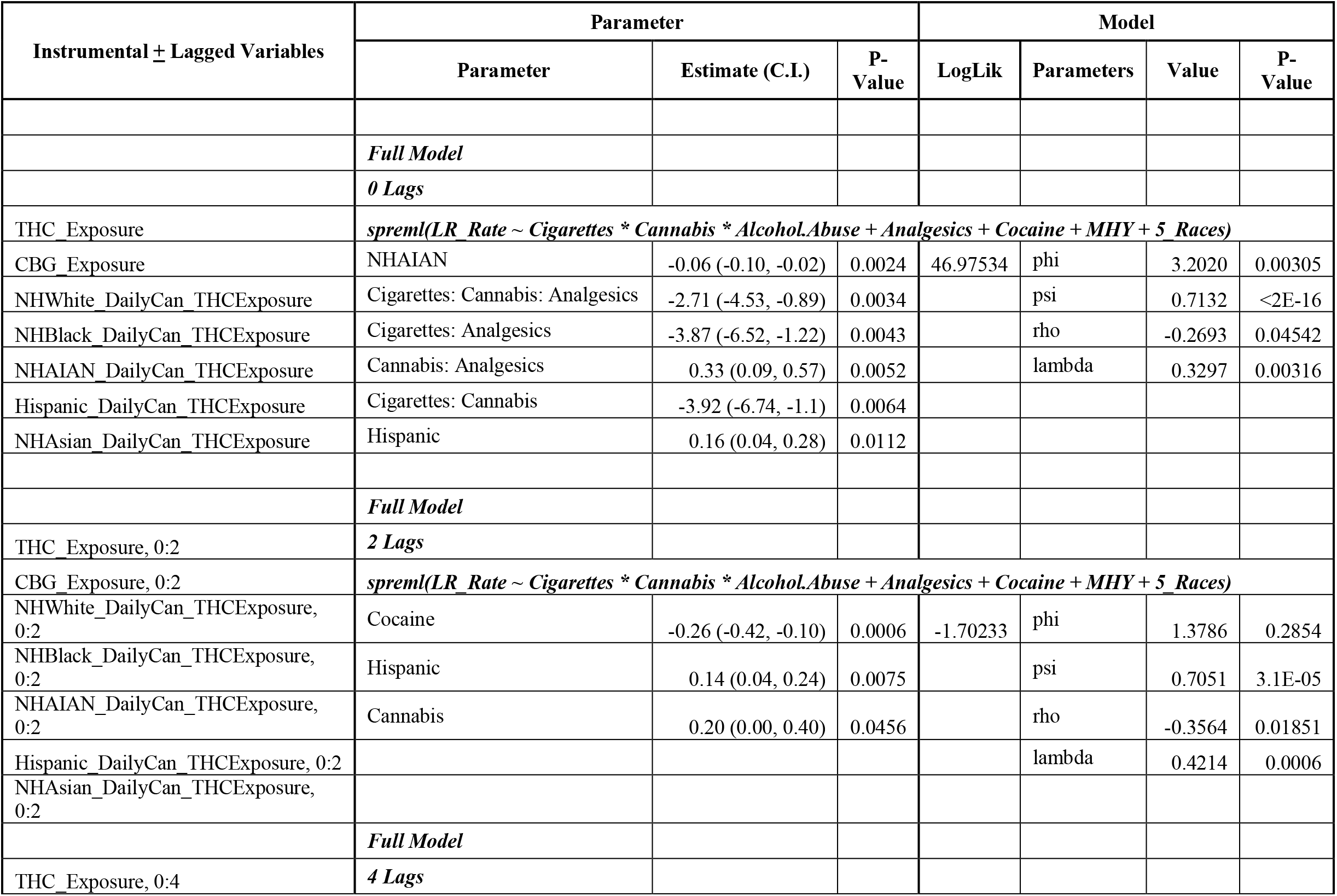
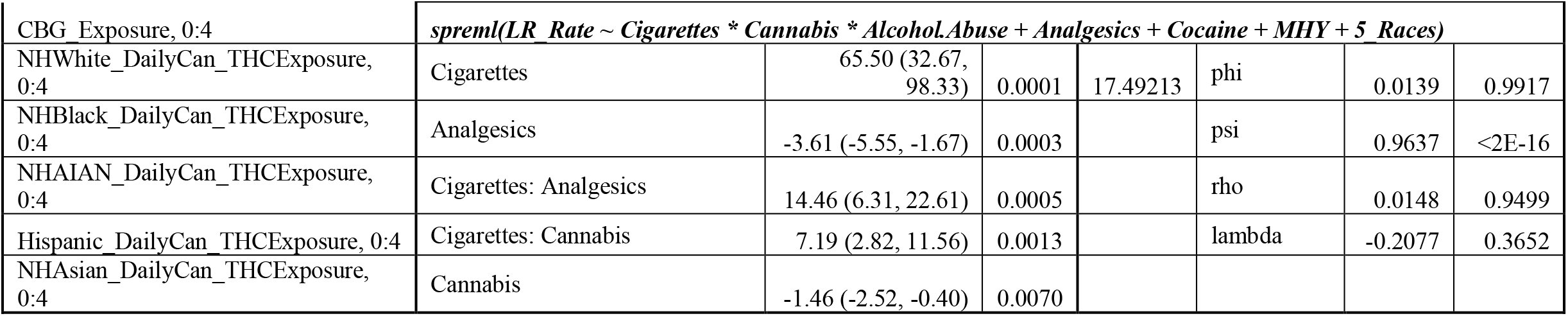
Spatial Regression with International Adjustment Factor for Silent Cases – 63%.

| Instrumental ± Lagged Variables | Parameter | | | Model | | | |
| --- | --- | --- | --- | --- | --- | --- | --- |
|  | Parameter | Estimate (C.I.) | P-Value | LogLik | Parameters | Value | P-Value |
|  | *Full Model* |  |  |  |  |  |  |
|  | *0 Lags* |  |  |  |  |  |  |
| THC Exposure | *spreml(LR_Rate ~ Cigarettes \* Cannabis \* Alcohol.Abuse + Analgesics + Cocaine + MHY + 5 Races)* |  |  |  |  |  |  |
| CBG Exposure | NHAIAN | -0.06 (-0.10, -0.02) | 0.0024 | 46.97534 | phi | 3.2020 | 0.00305 |
| NHWhite_DailyCan_THCExposure | Cigarettes: Cannabis: Analgesics | -2.71 (-4.53, -0.89) | 0.0034 |  | psi | 0.7132 | <2E-16 |
| NHBlack_DailyCan_THCExposure | Cigarettes: Analgesics | -3.87 (-6.52, -1.22) | 0.0043 |  | rho | -0.2693 | 0.04542 |
| NHAIAN_DailyCan_THCExposure | Cannabis: Analgesics | 0.33 (0.09, 0.57) | 0.0052 |  | lambda | 0.3297 | 0.00316 |
| Hispanic_DailyCan_THCExposure | Cigarettes: Cannabis | -3.92 (-6.74, -1.1) | 0.0064 |  |  |  |  |
| NHAsian_DailyCan_THCExposure | Hispanic | 0.16 (0.04, 0.28) | 0.0112 |  |  |  |  |
|  | *Full Model* |  |  |  |  |  |  |
| THC Exposure, 0:2 | *2 Lags* |  |  |  |  |  |  |
| CBG Exposure, 0:2 | *spreml(LR_Rate ~ Cigarettes \* Cannabis \* Alcohol.Abuse + Analgesics + Cocaine + MHY + 5 Races)* |  |  |  |  |  |  |
| NHWhite_DailyCan_THCExposure, 0:2 | Cocaine | -0.26 (-0.42, -0.10) | 0.0006 | -1.70233 | phi | 1.3786 | 0.2854 |
| NHBlack_DailyCan_THCExposure, 0:2 | Hispanic | 0.14 (0.04, 0.24) | 0.0075 |  | psi | 0.7051 | 3.1E-05 |
| NHAIAN_DailyCan_THCExposure, 0:2 | Cannabis | 0.20 (0.00, 0.40) | 0.0456 |  | rho | -0.3564 | 0.01851 |
| Hispanic_DailyCan_THCExposure, 0:2 |  |  |  |  | lambda | 0.4214 | 0.0006 |
| NHAsian_DailyCan_THCExposure, 0:2 |  |  |  |  |  |  |  |
|  | *Full Model* |  |  |  |  |  |  |
| THC Exposure, 0:4 | *4 Lags* |  |  |  |  |  |  |

| CBG Exposure, 0:4 | *spreml(LR Rate ~ Cigarettes \* Cannabis \* Alcohol.Abuse + Analgesics + Cocaine + MHY + 5 Races)* |  |  |  |  |  |  |
| --- | --- | --- | --- | --- | --- | --- | --- |
| NHWhite_DailyCan_THCExposure, 0:4 | Cigarettes | 65.50 (32.67, 98.33) | 0.0001 | 17.49213 | phi | 0.0139 | 0.9917 |
| NHBlack_DailyCan_THCExposure, 0:4 | Analgesics | -3.61 (-5.55, -1.67) | 0.0003 |  | psi | 0.9637 | <2E-16 |
| NHAIAN_DailyCan_THCExposure, 0:4 | Cigarettes: Analgesics | 14.46 (6.31, 22.61) | 0.0005 |  | rho | 0.0148 | 0.9499 |
| Hispanic_DailyCan_THCExposure, 0:4 | Cigarettes: Cannabis | 7.19 (2.82, 11.56) | 0.0013 |  | lambda | -0.2077 | 0.3652 |
| NHAsian_DailyCan_THCExposure, 0:4 | Cannabis | -1.46 (-2.52, -0.40) | 0.0070 |  |  |  |  |

**Supplementary Table 11.:**
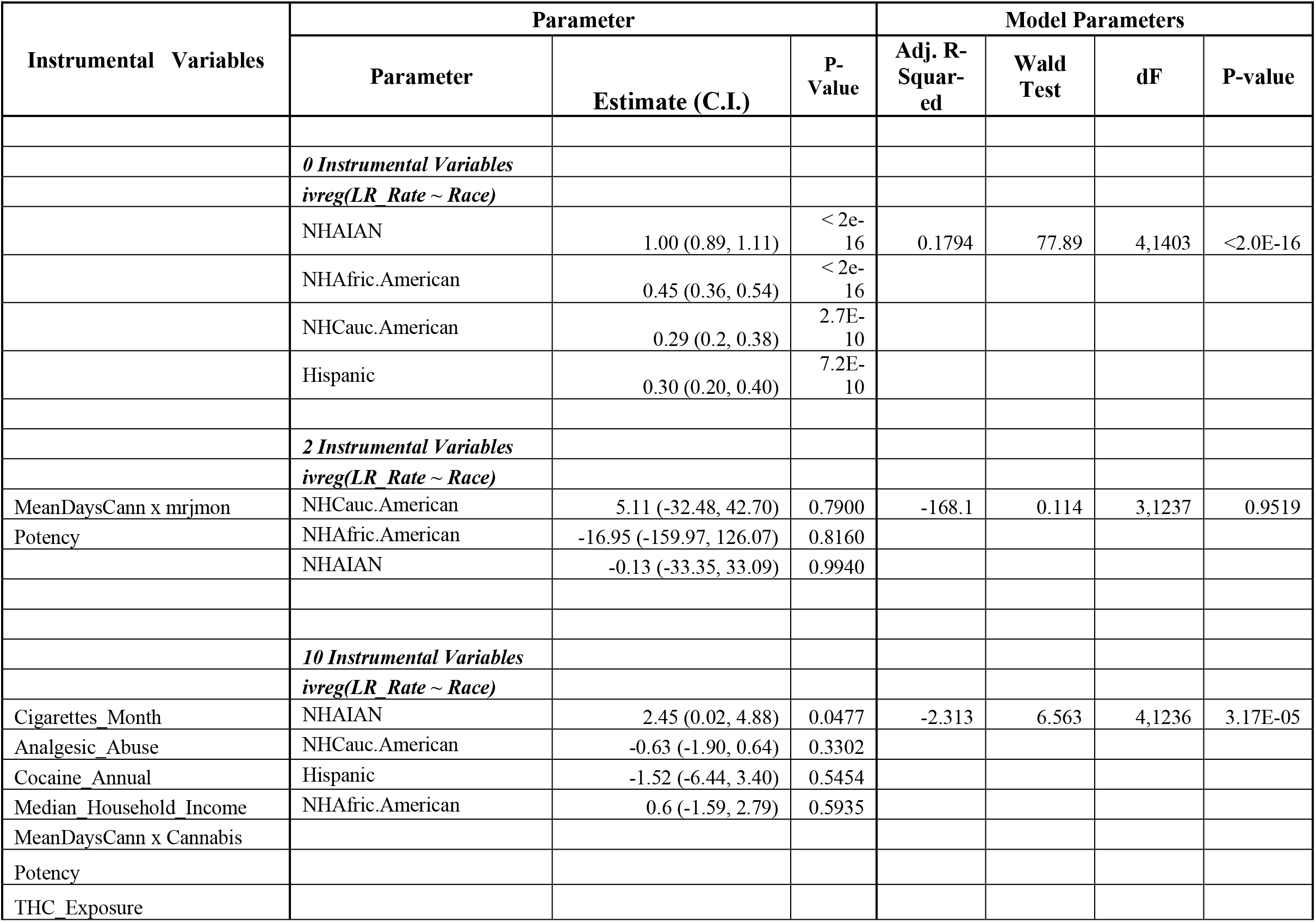
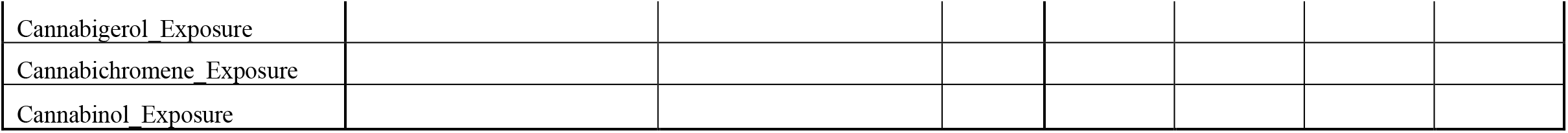
Two-Step Instrumental Regression by Race.

| Instrumental Variables | Parameter |  |  | Model Parameters |  |  |  |
| --- | --- | --- | --- | --- | --- | --- | --- |
|  | Parameter | Estimate (C.I.) | P-Value | Adj. R-Squared | Wald Test | dF | P-value |
|  | *0 Instrumental Variables* |  |  |  |  |  |  |
|  | *ivreg(LR_Rate ~ Race)* |  |  |  |  |  |  |
|  | NHAIAN | 1.00 (0.89, 1.11) | < 2e-16 | 0.1794 | 77.89 | 4,1403 | <2.0E-16 |
|  | NHAfric.American | 0.45 (0.36, 0.54) | < 2e-16 |  |  |  |  |
|  | NHCauc.American | 0.29 (0.2, 0.38) | 2.7E-10 |  |  |  |  |
|  | Hispanic | 0.30 (0.20, 0.40) | 7.2E-10 |  |  |  |  |
|  | *2 Instrumental Variables* |  |  |  |  |  |  |
|  | *ivreg(LR_Rate ~ Race)* |  |  |  |  |  |  |
| MeanDaysCann x mrjmon | NHCauc.American | 5.11 (-32.48, 42.70) | 0.7900 | -168.1 | 0.114 | 3,1237 | 0.9519 |
| Potency | NHAfric.American | -16.95 (-159.97, 126.07) | 0.8160 |  |  |  |  |
|  | NHAIAN | -0.13 (-33.35, 33.09) | 0.9940 |  |  |  |  |
|  | *10 Instrumental Variables* |  |  |  |  |  |  |
|  | *ivreg(LR_Rate ~ Race)* |  |  |  |  |  |  |
| Cigarettes Month | NHAIAN | 2.45 (0.02, 4.88) | 0.0477 | -2.313 | 6.563 | 4,1236 | 3.17E-05 |
| Analgesic Abuse | NHCauc.American | -0.63 (-1.90, 0.64) | 0.3302 |  |  |  |  |
| Cocaine Annual | Hispanic | -1.52 (-6.44, 3.40) | 0.5454 |  |  |  |  |
| Median Household Income | NHAfric.American | 0.6 (-1.59, 2.79) | 0.5935 |  |  |  |  |
| MeanDaysCann x Cannabis |  |  |  |  |  |  |  |
| Potency |  |  |  |  |  |  |  |
| THC Exposure |  |  |  |  |  |  |  |
| Cannabigerol Exposure |  |  |  |  |  |  |  |
| Cannabichromene Exposure |  |  |  |  |  |  |  |
| Cannabinol Exposure |  |  |  |  |  |  |  |

**Supplementary Table 12.:** Numbers of Cases and Controls in States with Legal Cannabis vs. Other Paradigms.

| Dichotomized Status | No. Limb Reductions | No. Limb Reductions 37% Silent Factor | No. Limb Reductions 63% Silent Factor | Total Births | Normal Births | Normal Births 37% Silent Factor | Normal Births 63% Silent Factor |
| --- | --- | --- | --- | --- | --- | --- | --- |
| Other | 14,398 | 21,106 | 31,954 | 33,645,176 | 33,630,778 | 33,624,070 | 33,613,222 |
| Legal | 215 | 338 | 566 | 429,020 | 428,805 | 428,682 | 428,454 |

**Supplementary Table 13:**
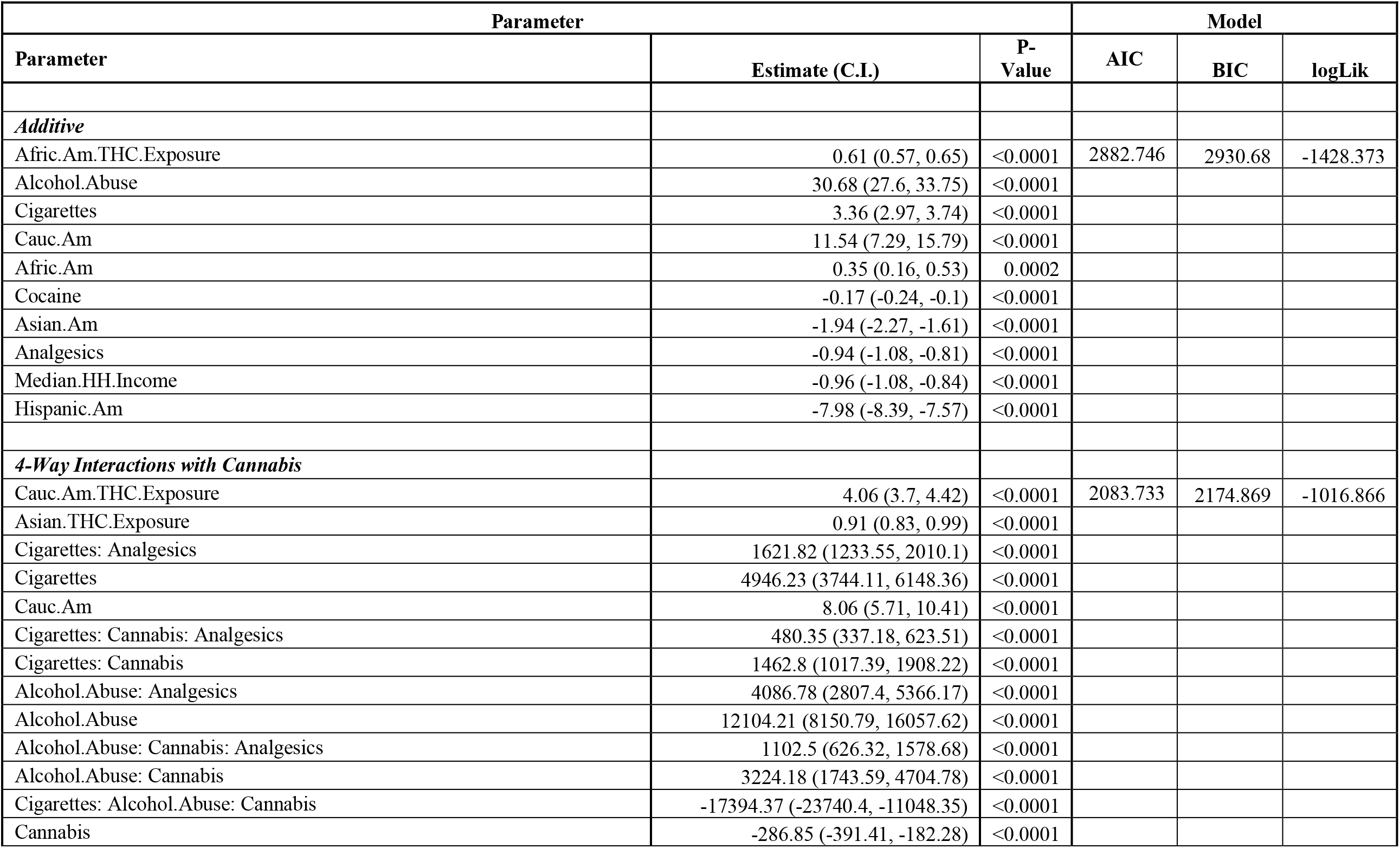
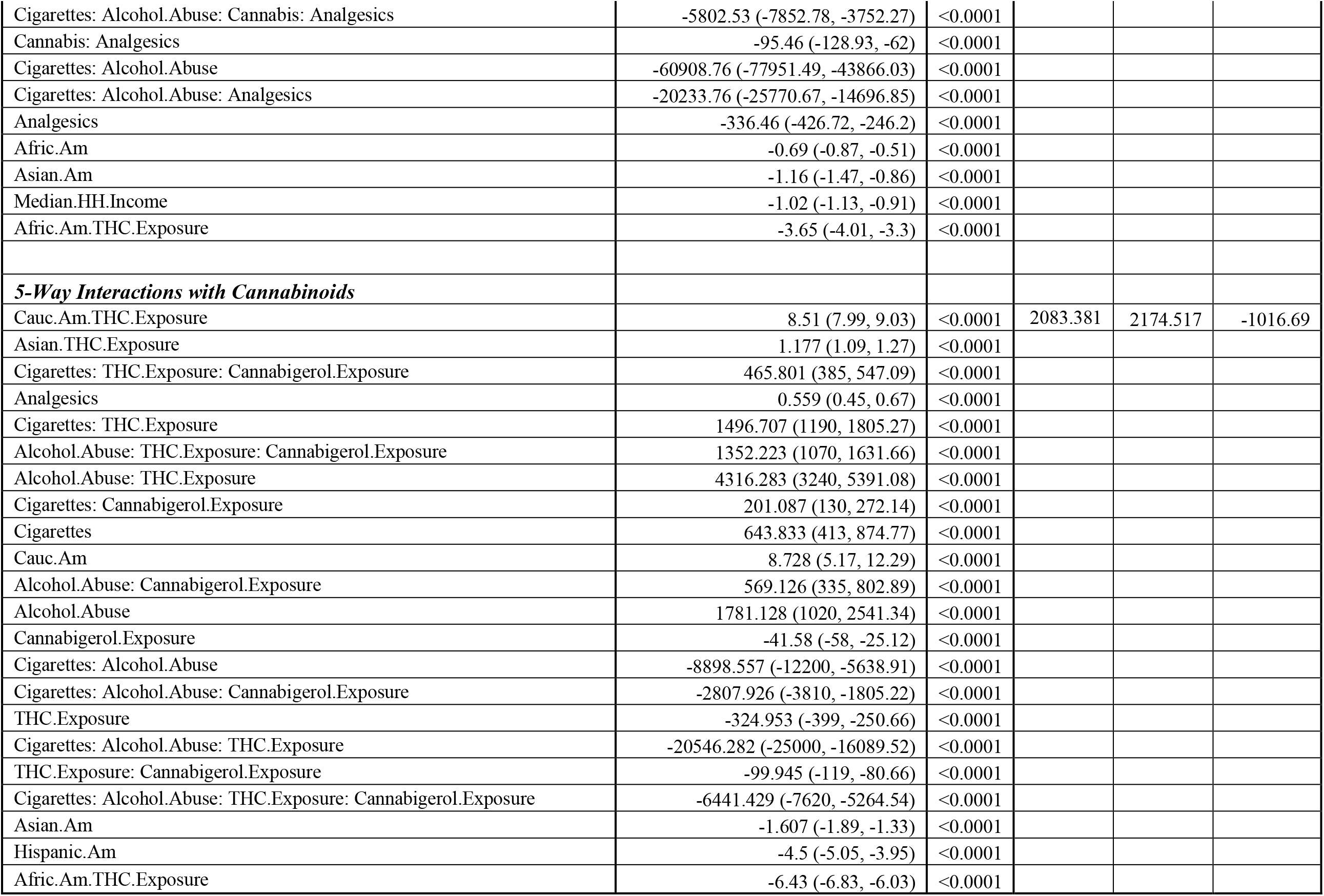
Inverse Probability-Weighted Mixed Effects Regression – Final Models.

| Parameter |  |  | Model |  |  |
| --- | --- | --- | --- | --- | --- |
| Parameter | Estimate (C.I.) | P-Value | AIC | BIC | logLik |
| *Additive* |  |  |  |  |  |
| Afric.Am.THc.Exposure | 0.61 (0.57, 0.65) | <0.0001 | 2882.746 | 2930.68 | -1428.373 |
| Alcohol.Abuse | 30.68 (27.6, 33.75) | <0.0001 |  |  |  |
| Cigarettes | 3.36 (2.97, 3.74) | <0.0001 |  |  |  |
| Cauc.Am | 11.54 (7.29, 15.79) | <0.0001 |  |  |  |
| Afric.Am | 0.35 (0.16, 0.53) | 0.0002 |  |  |  |
| Cocaine | -0.17 (-0.24, -0.1) | <0.0001 |  |  |  |
| Asian.Am | -1.94 (-2.27, -1.61) | <0.0001 |  |  |  |
| Analgesics | -0.94 (-1.08, -0.81) | <0.0001 |  |  |  |
| Median.HH.Income | -0.96 (-1.08, -0.84) | <0.0001 |  |  |  |
| Hispanic.Am | -7.98 (-8.39, -7.57) | <0.0001 |  |  |  |
| *4-Way Interactions with Cannabis* |  |  |  |  |  |
| Cauc.Am.THc.Exposure | 4.06 (3.7, 4.42) | <0.0001 | 2083.733 | 2174.869 | -1016.866 |
| Asian.THc.Exposure | 0.91 (0.83, 0.99) | <0.0001 |  |  |  |
| Cigarettes: Analgesics | 1621.82 (1233.55, 2010.1) | <0.0001 |  |  |  |
| Cigarettes | 4946.23 (3744.11, 6148.36) | <0.0001 |  |  |  |
| Cauc.Am | 8.06 (5.71, 10.41) | <0.0001 |  |  |  |
| Cigarettes: Cannabis: Analgesics | 480.35 (337.18, 623.51) | <0.0001 |  |  |  |
| Cigarettes: Cannabis | 1462.8 (1017.39, 1908.22) | <0.0001 |  |  |  |
| Alcohol.Abuse: Analgesics | 4086.78 (2807.4, 5366.17) | <0.0001 |  |  |  |
| Alcohol.Abuse | 12104.21 (8150.79, 16057.62) | <0.0001 |  |  |  |
| Alcohol.Abuse: Cannabis: Analgesics | 1102.5 (626.32, 1578.68) | <0.0001 |  |  |  |
| Alcohol.Abuse: Cannabis | 3224.18 (1743.59, 4704.78) | <0.0001 |  |  |  |
| Cigarettes: Alcohol.Abuse: Cannabis | -17394.37 (-23740.4, -11048.35) | <0.0001 |  |  |  |
| Cannabis | -286.85 (-391.41, -182.28) | <0.0001 |  |  |  |
| Cigarettes: Alcohol.Abuse: Cannabis: Analgesics | -5802.53 (-7852.78, -3752.27) | <0.0001 |  |  |  |
| Cannabis: Analgesics | -95.46 (-128.93, -62) | <0.0001 |  |  |  |
| Cigarettes: Alcohol.Abuse | -60908.76 (-77951.49, -43866.03) | <0.0001 |  |  |  |
| Cigarettes: Alcohol.Abuse: Analgesics | -20233.76 (-25770.67, -14696.85) | <0.0001 |  |  |  |
| Analgesics | -336.46 (-426.72, -246.2) | <0.0001 |  |  |  |
| Afric.Am | -0.69 (-0.87, -0.51) | <0.0001 |  |  |  |
| Asian.Am | -1.16 (-1.47, -0.86) | <0.0001 |  |  |  |
| Median.HH.Income | -1.02 (-1.13, -0.91) | <0.0001 |  |  |  |
| Afric.Am. THC.Exposure | -3.65 (-4.01, -3.3) | <0.0001 |  |  |  |
| **5-Way Interactions with Cannabinoids** |  |  |  |  |  |
| Cauc.Am. THC.Exposure | 8.51 (7.99, 9.03) | <0.0001 | 2083.381 | 2174.517 | -1016.69 |
| Asian. THC.Exposure | 1.177 (1.09, 1.27) | <0.0001 |  |  |  |
| Cigarettes: THC.Exposure: Cannabigerol.Exposure | 465.801 (385, 547.09) | <0.0001 |  |  |  |
| Analgesics | 0.559 (0.45, 0.67) | <0.0001 |  |  |  |
| Cigarettes: THC.Exposure | 1496.707 (1190, 1805.27) | <0.0001 |  |  |  |
| Alcohol.Abuse: THC.Exposure: Cannabigerol.Exposure | 1352.223 (1070, 1631.66) | <0.0001 |  |  |  |
| Alcohol.Abuse: THC.Exposure | 4316.283 (3240, 5391.08) | <0.0001 |  |  |  |
| Cigarettes: Cannabigerol.Exposure | 201.087 (130, 272.14) | <0.0001 |  |  |  |
| Cigarettes | 643.833 (413, 874.77) | <0.0001 |  |  |  |
| Cauc.Am | 8.728 (5.17, 12.29) | <0.0001 |  |  |  |
| Alcohol.Abuse: Cannabigerol.Exposure | 569.126 (335, 802.89) | <0.0001 |  |  |  |
| Alcohol.Abuse | 1781.128 (1020, 2541.34) | <0.0001 |  |  |  |
| Cannabigerol.Exposure | -41.58 (-58, -25.12) | <0.0001 |  |  |  |
| Cigarettes: Alcohol.Abuse | -8898.557 (-12200, -5638.91) | <0.0001 |  |  |  |
| Cigarettes: Alcohol.Abuse: Cannabigerol.Exposure | -2807.926 (-3810, -1805.22) | <0.0001 |  |  |  |
| THC.Exposure | -324.953 (-399, -250.66) | <0.0001 |  |  |  |
| Cigarettes: Alcohol.Abuse: THC.Exposure | -20546.282 (-25000, -16089.52) | <0.0001 |  |  |  |
| THC.Exposure: Cannabigerol.Exposure | -99.945 (-119, -80.66) | <0.0001 |  |  |  |
| Cigarettes: Alcohol.Abuse: THC.Exposure: Cannabigerol.Exposure | -6441.429 (-7620, -5264.54) | <0.0001 |  |  |  |
| Asian.Am | -1.607 (-1.89, -1.33) | <0.0001 |  |  |  |
| Hispanic.Am | -4.5 (-5.05, -3.95) | <0.0001 |  |  |  |
| Afric.Am. THC.Exposure | -6.43 (-6.83, -6.03) | <0.0001 |  |  |  |

## Supplementary Figures

**Supplementary Figure 1.:**
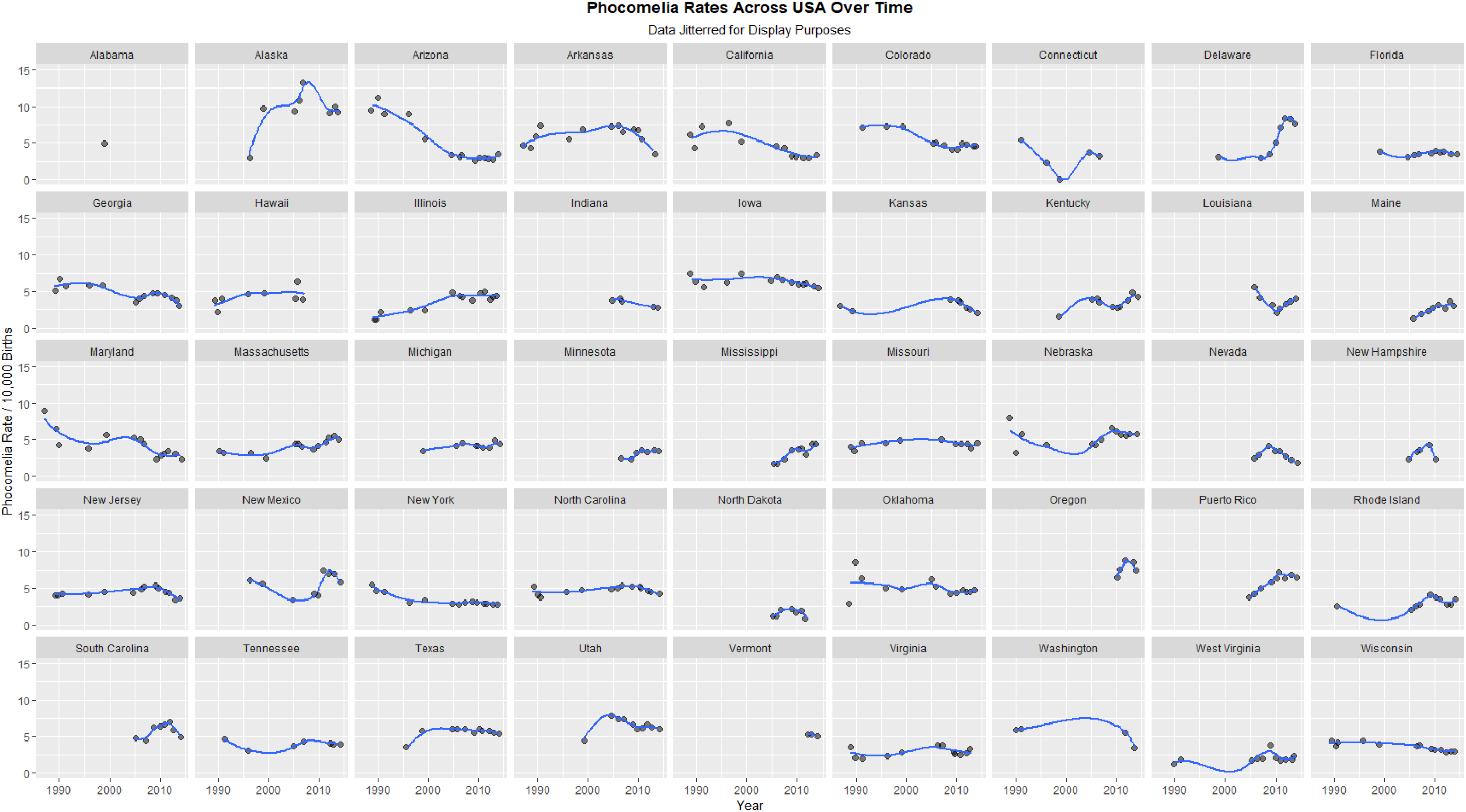
Limb Reduction Rate by State – Facetted Plot.

**Supplementary Figure 2.:**
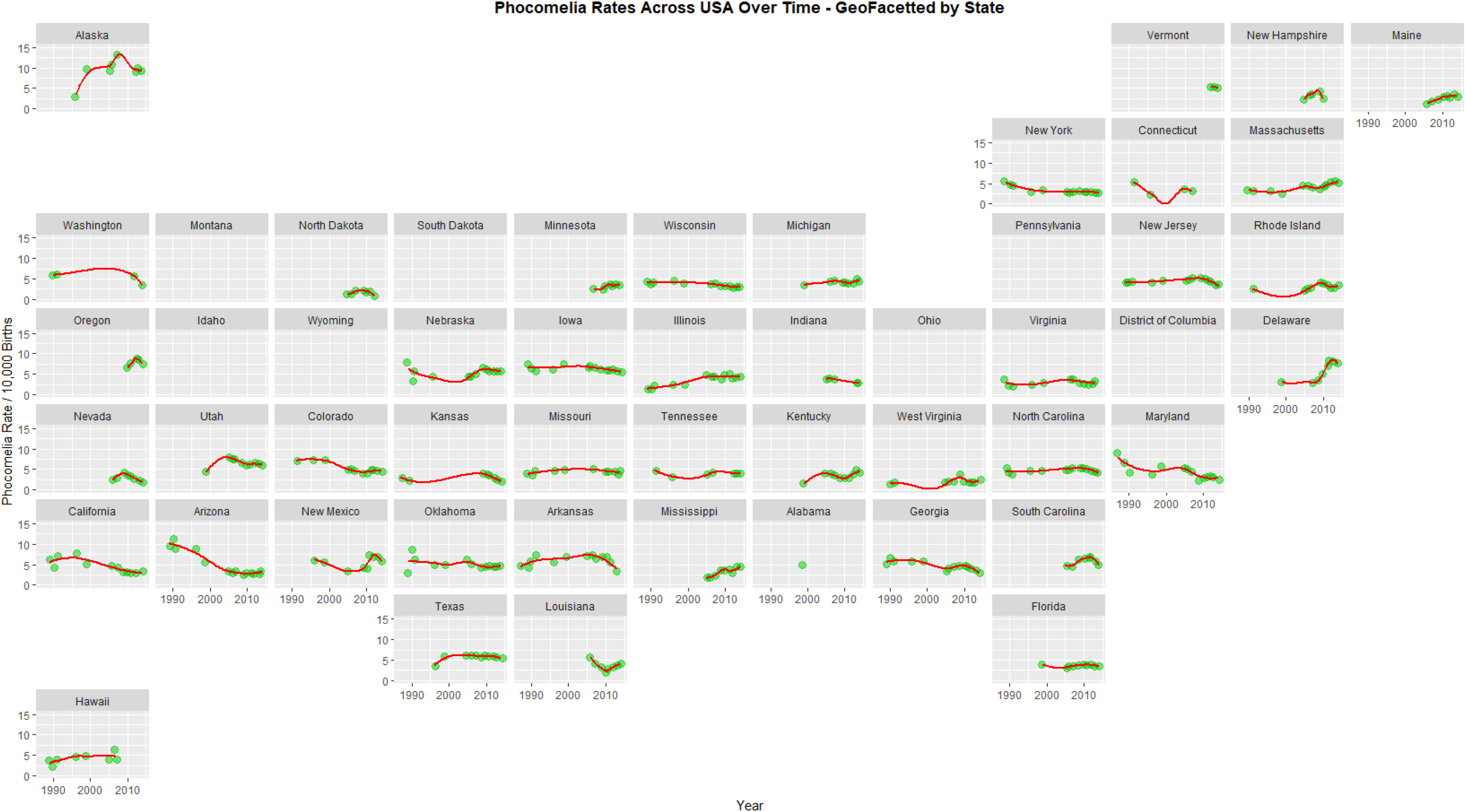
Limb Reduction Rate by State – Geo-facetted Plot.

**Supplementary Figure 3.:**
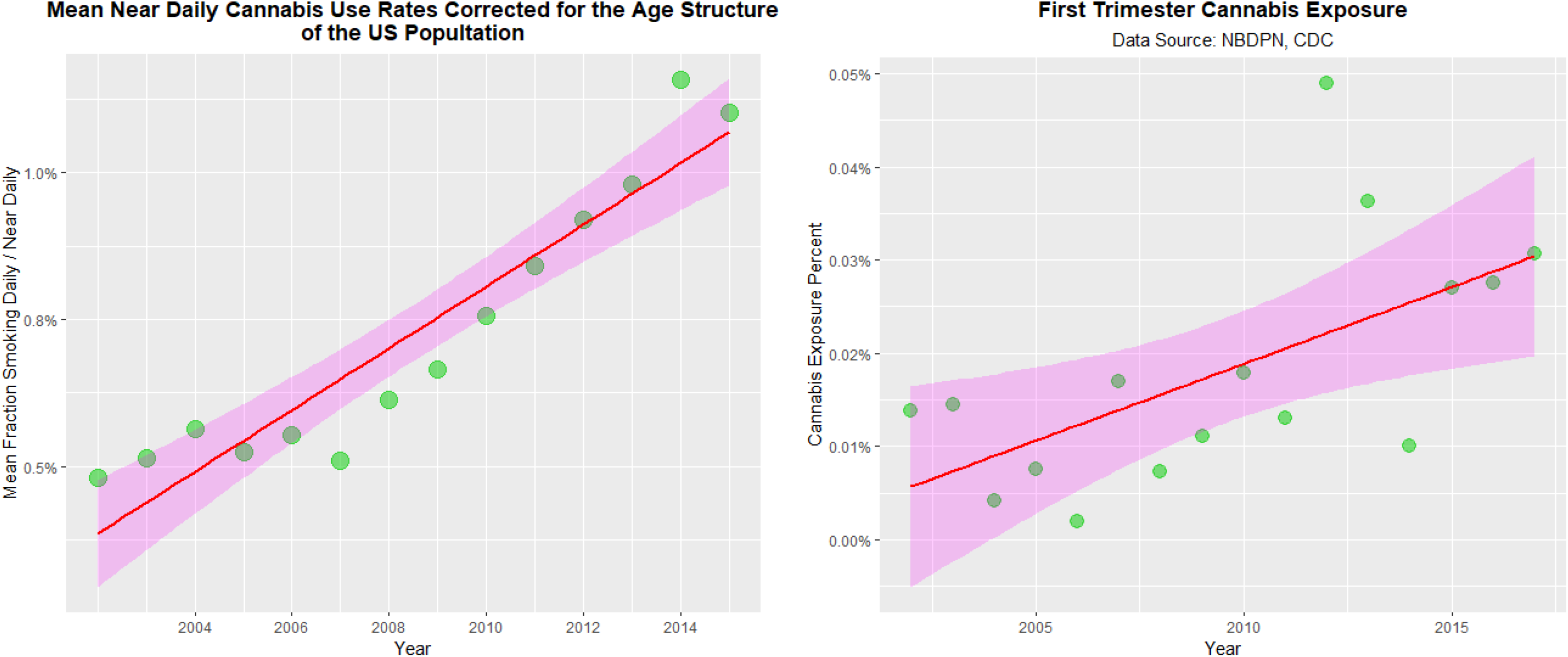
Daily Cannabis Use – General and in Pregnancy.

**Supplementary Figure 4.:**
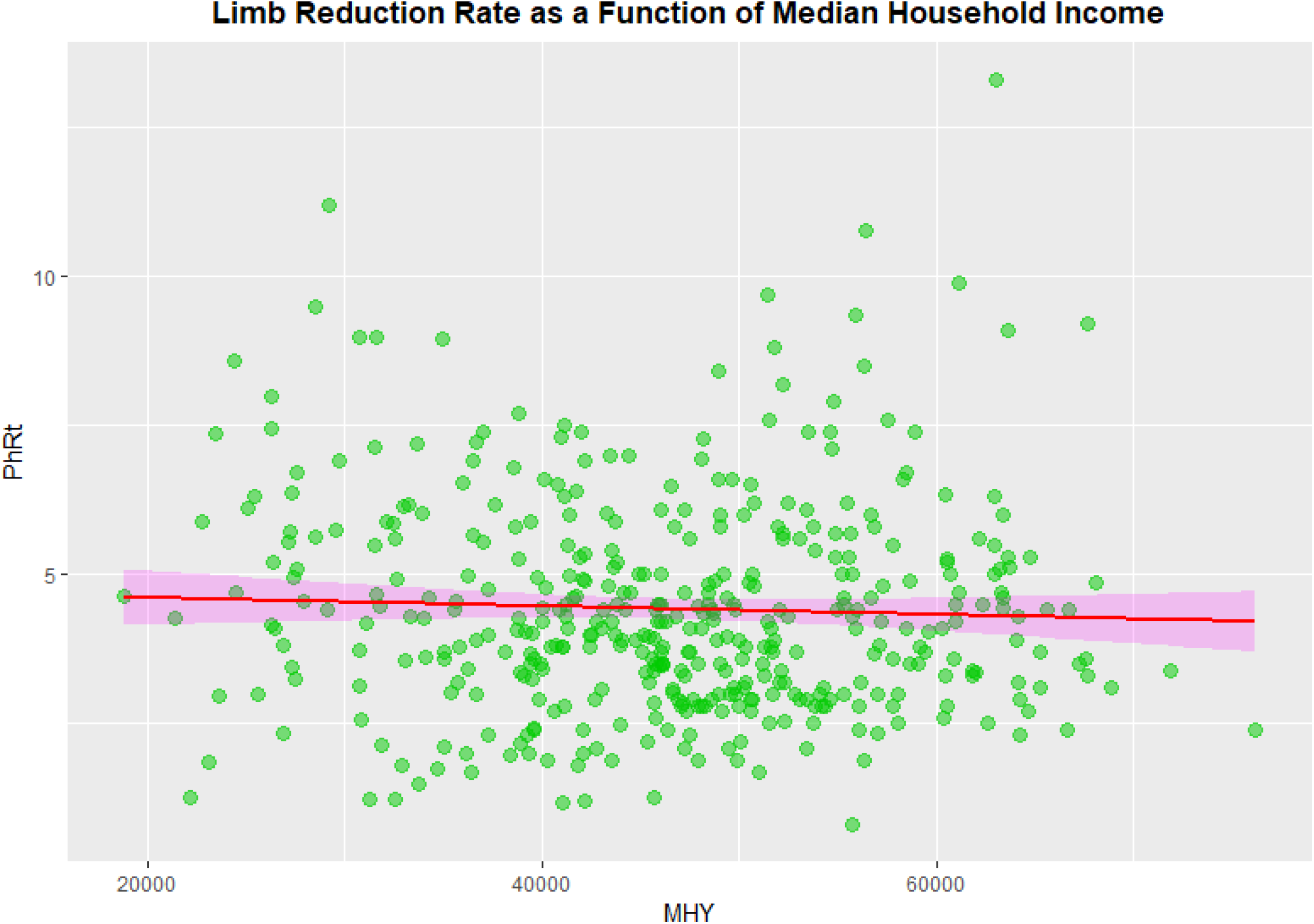
Limb Reduction Rate Median Household Income (MHY)

**Supplementary Figure 5.:**
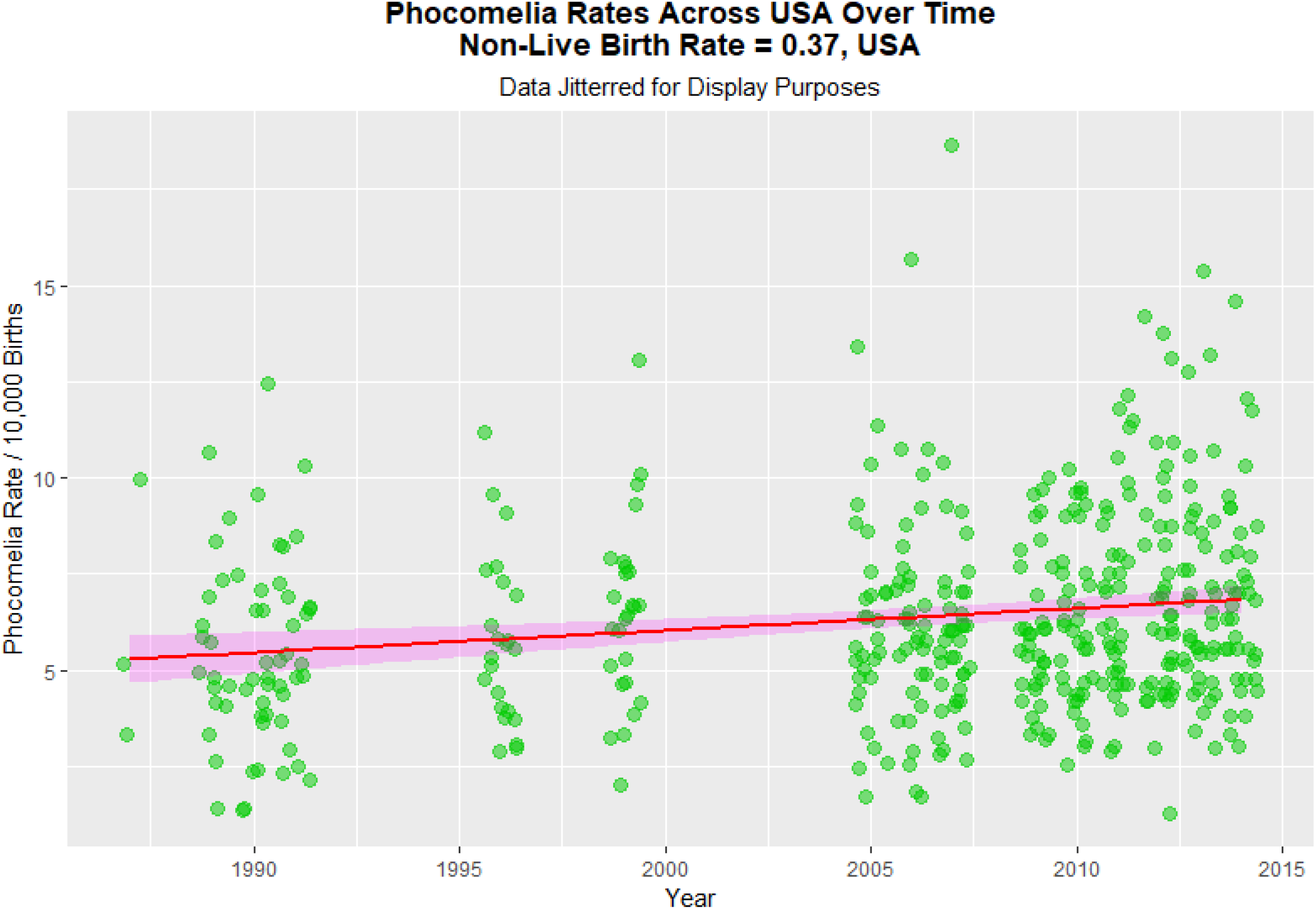
LR Non-Live Birth Rate – Scaled with USA Silent Factor, 37%.

**Supplementary Figure 6:**
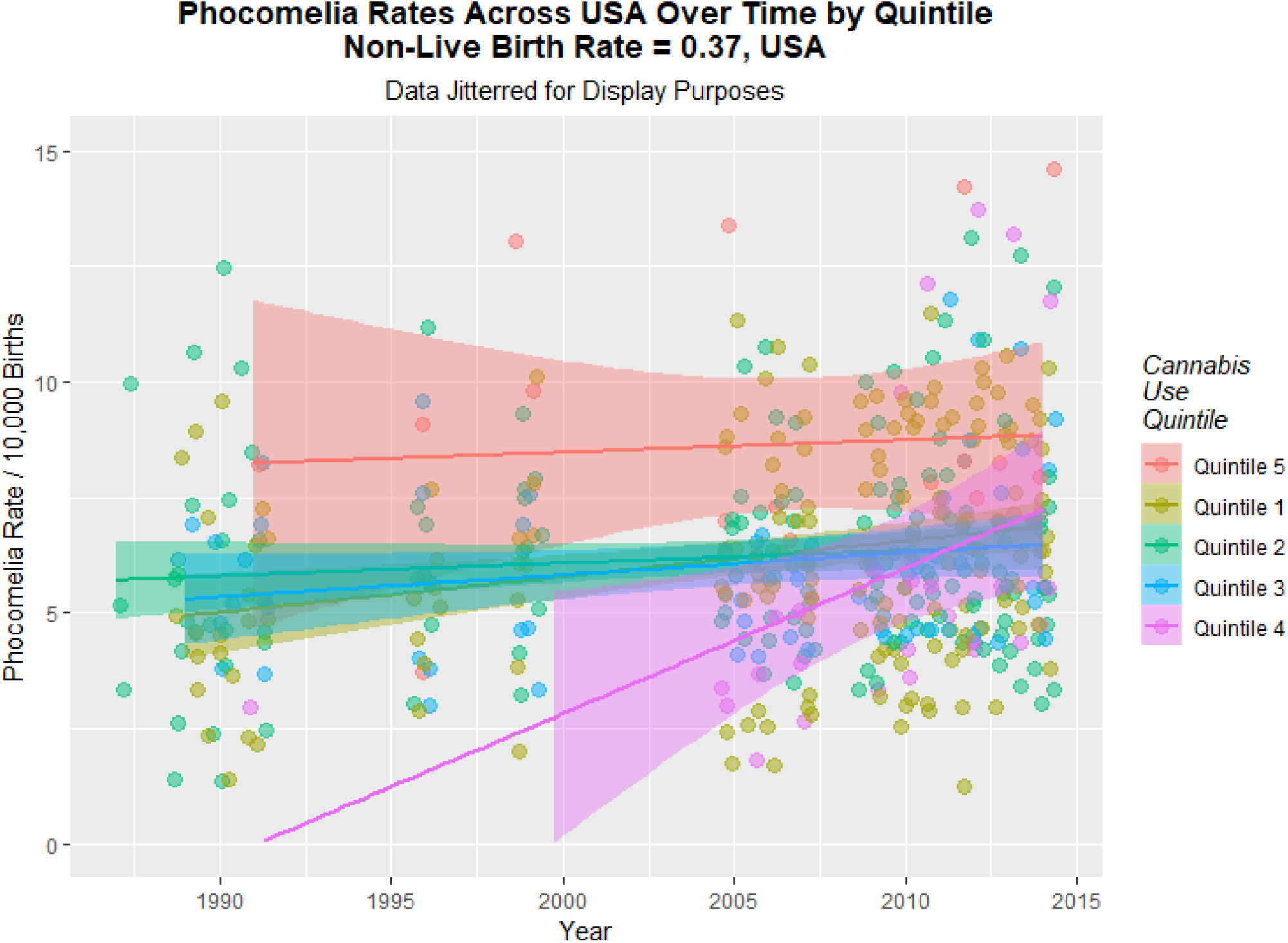
LRR Over Time by Quintile Corrected for USA Silent Factor.

**Supplementary Figure 7:**
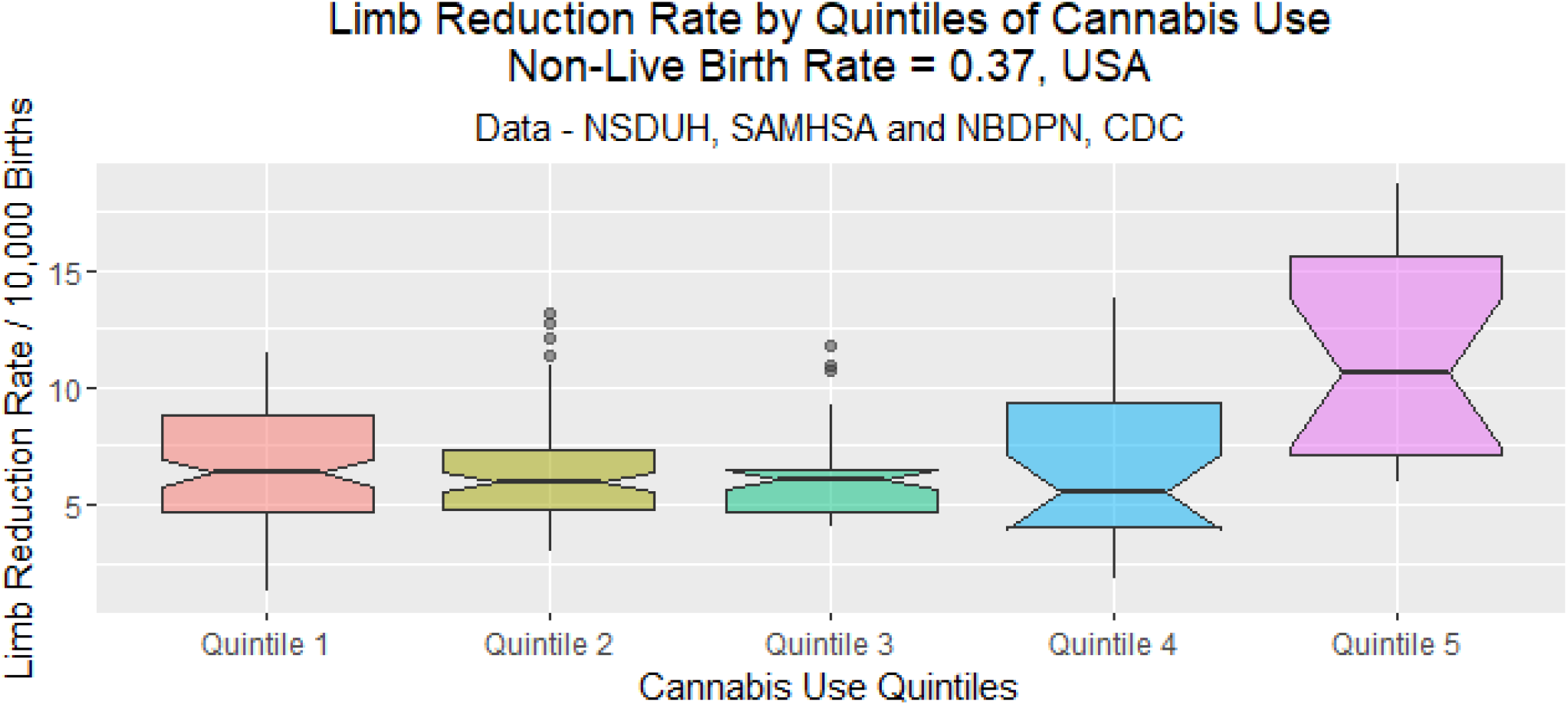
LRR Boxplots by Quintile Corrected for USA Silent Factor.

**Supplementary Figure 8.:**
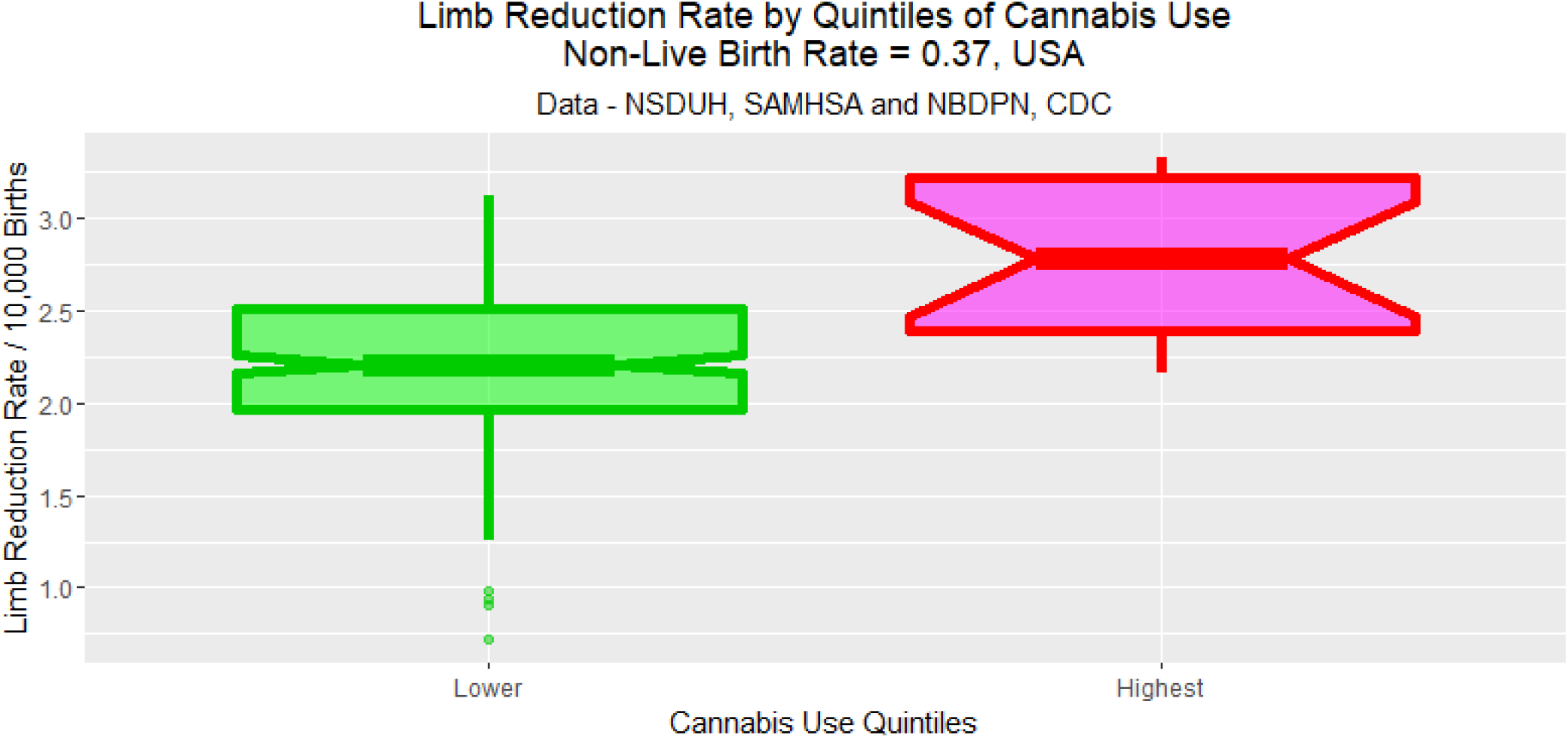
LRR Boxplots by Dichotomized Quintile Corrected for USA Silent Factor.

**Supplementary Figure 9.:**
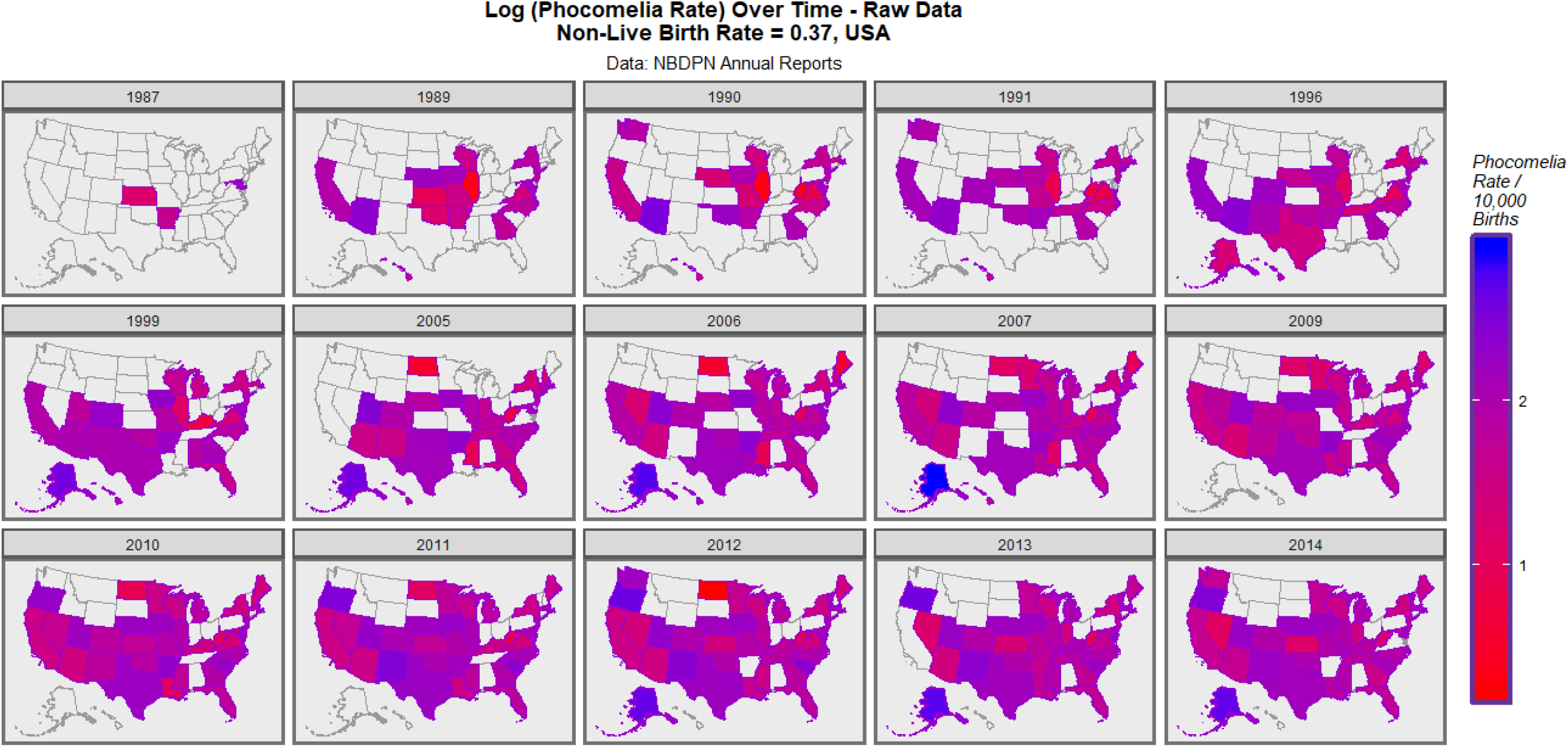
Map-Graphs of LRR Corrected for USA Silent Factor – Raw Data.

**Supplementary Figure 10.:**
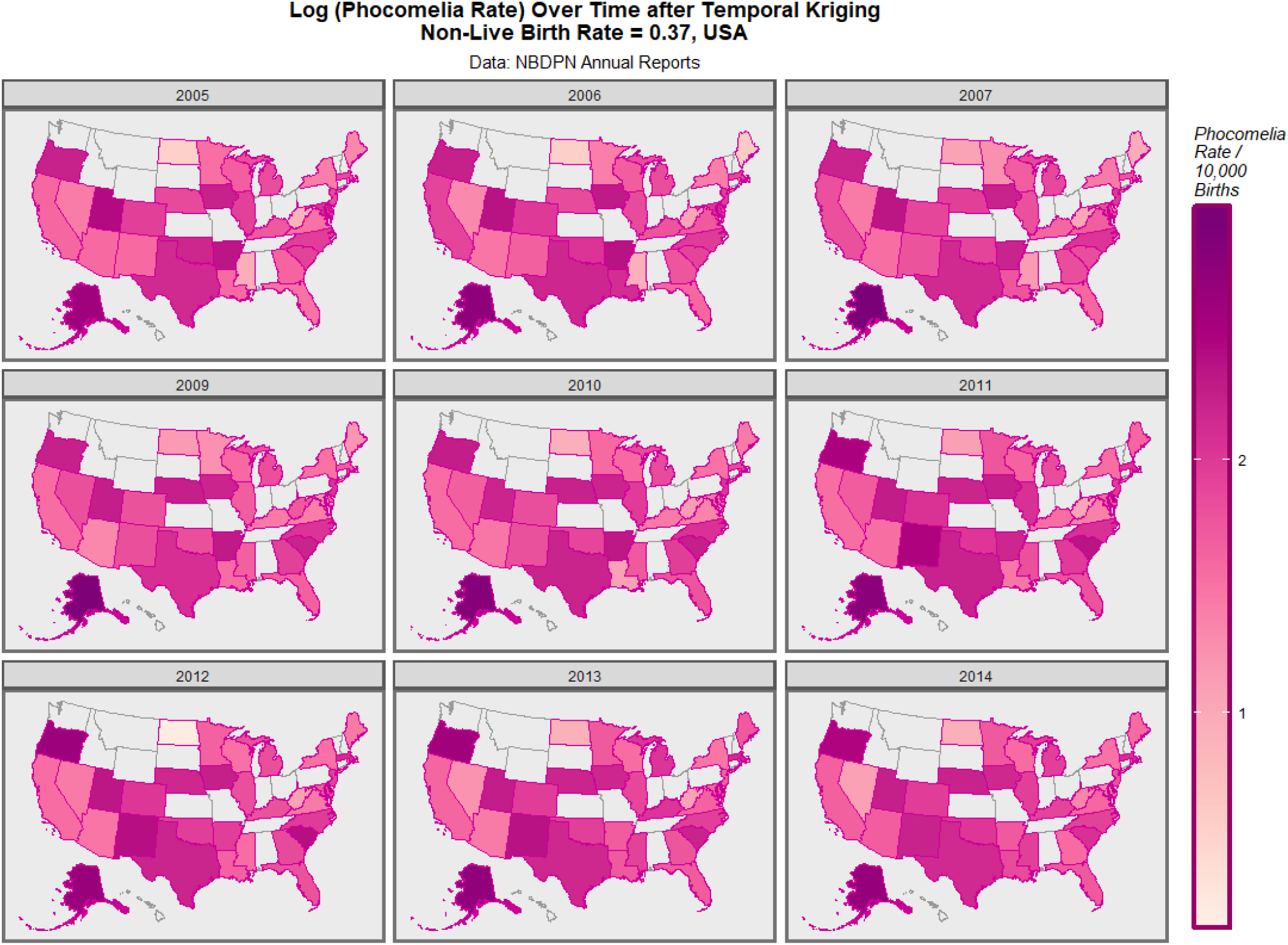
Map-Graphs of LRR Corrected for USA Silent Factor – Kriged Data.

**Supplementary Figure 11.:**
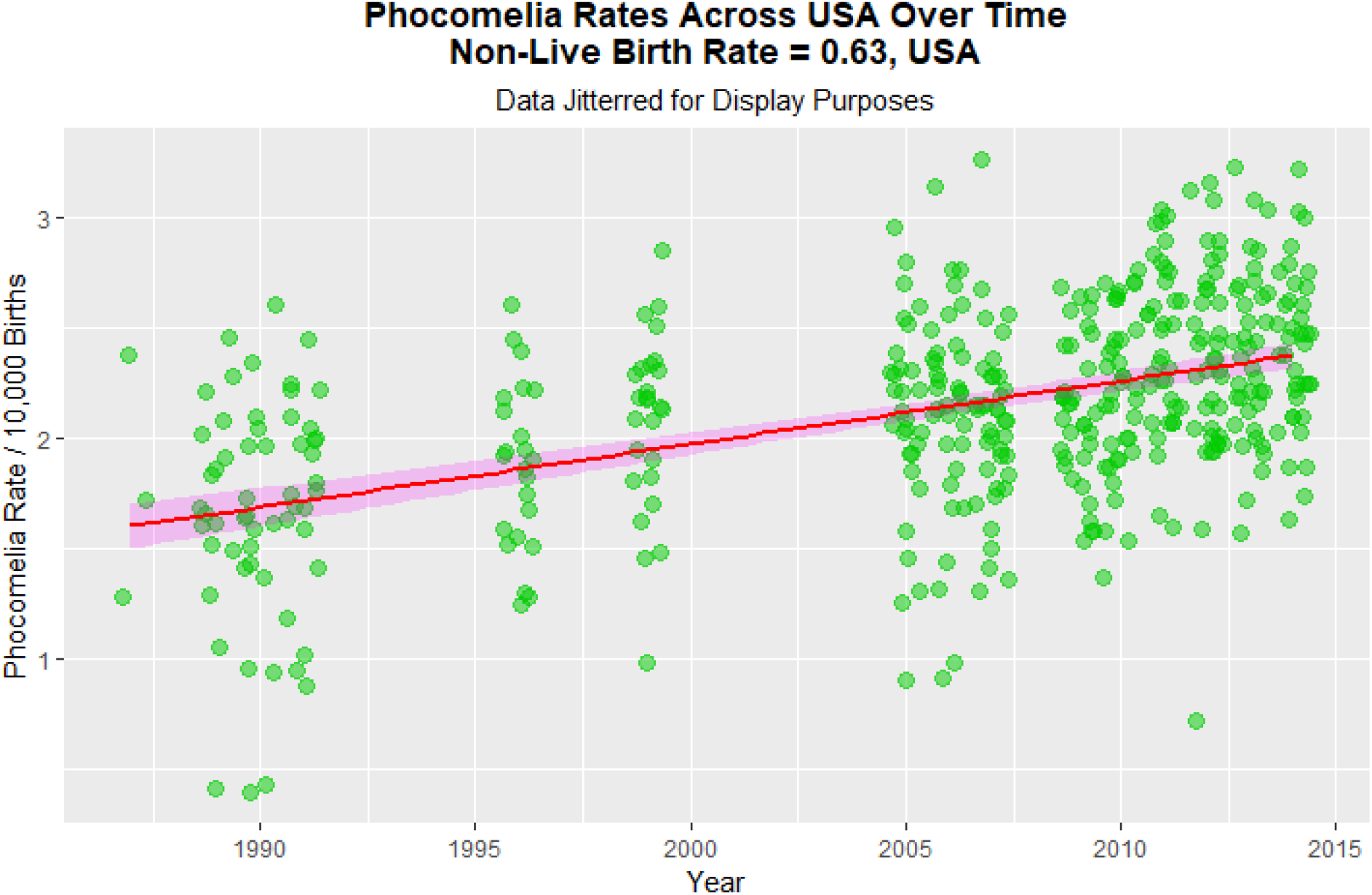
LR Non-Live Birth Rate Over Time – Scaled with International Silent Factor, 63%.

**Supplementary Figure 12:**
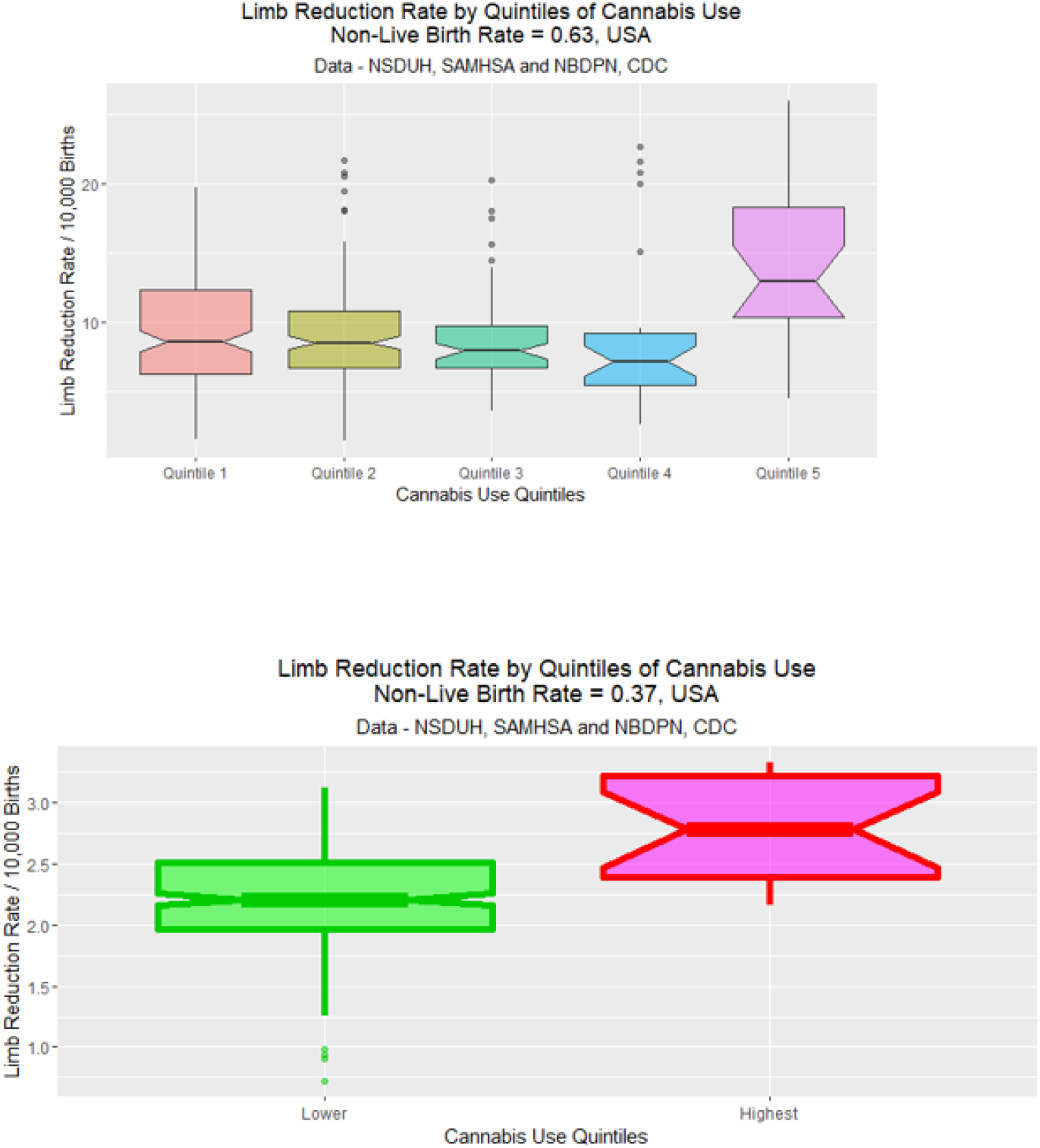
LR Non-Live Birth Rate Boxplots – Scaled with International Silent Factor, 63%.

**Supplementary Figure 13.:**
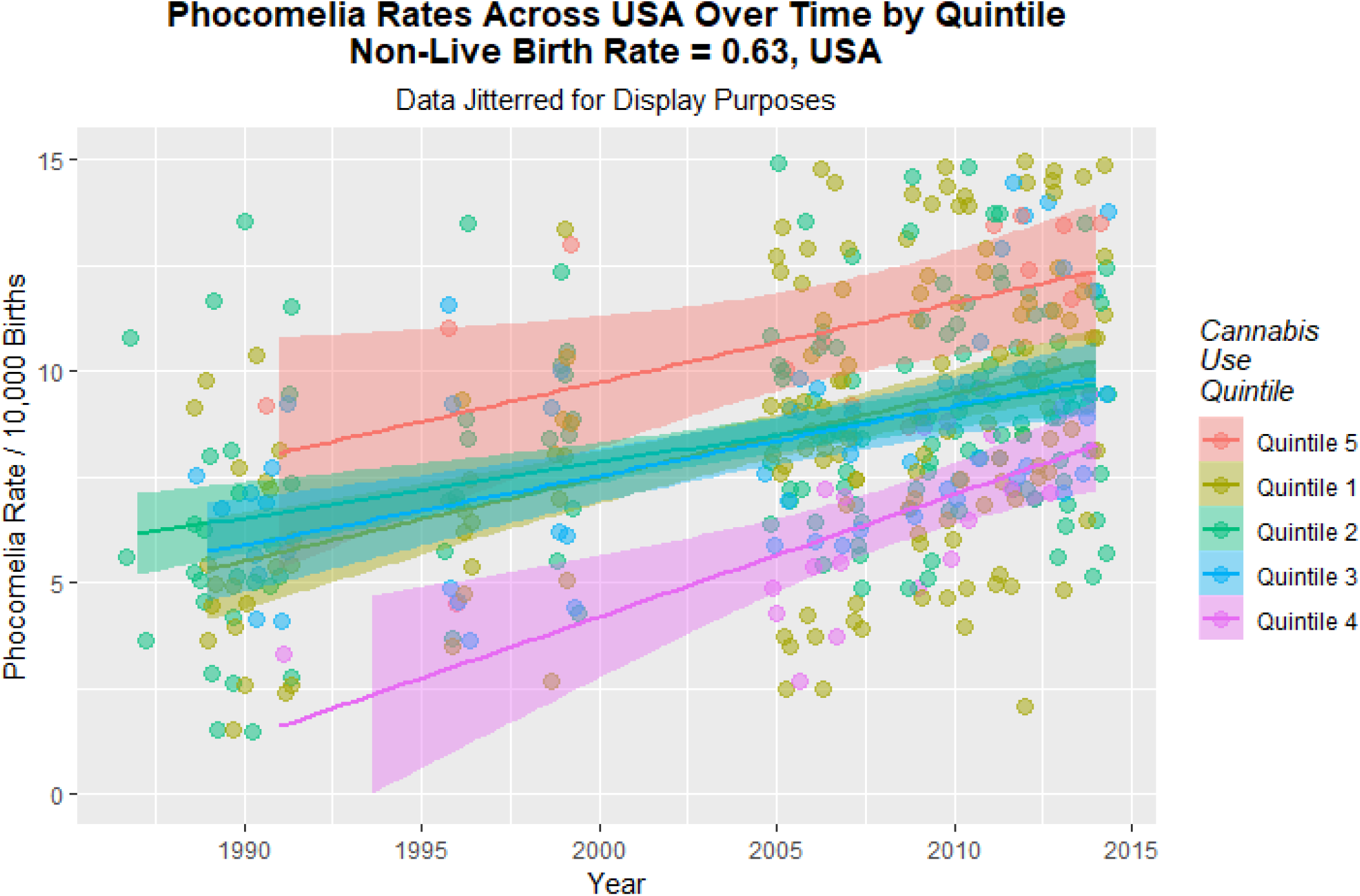
LR Non-Live Birth Rate Over Time by Quintiles – Scaled with International Silent Factor.

**Supplementary Figure 14.:**
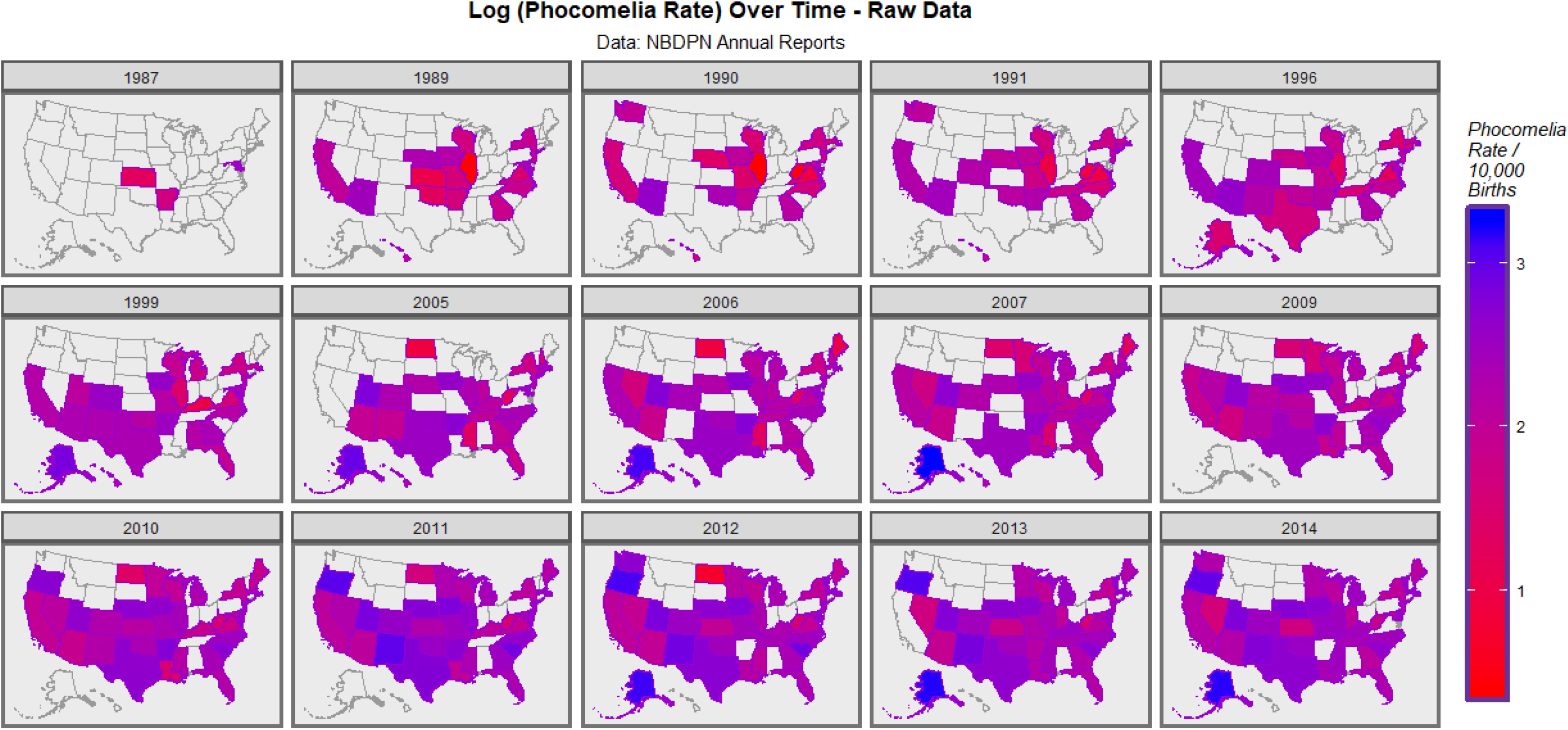
Map-Graphs of LRR Over Time – Adjusted with International Silent Factor – Raw Data.

**Supplementary Figure 15.:**
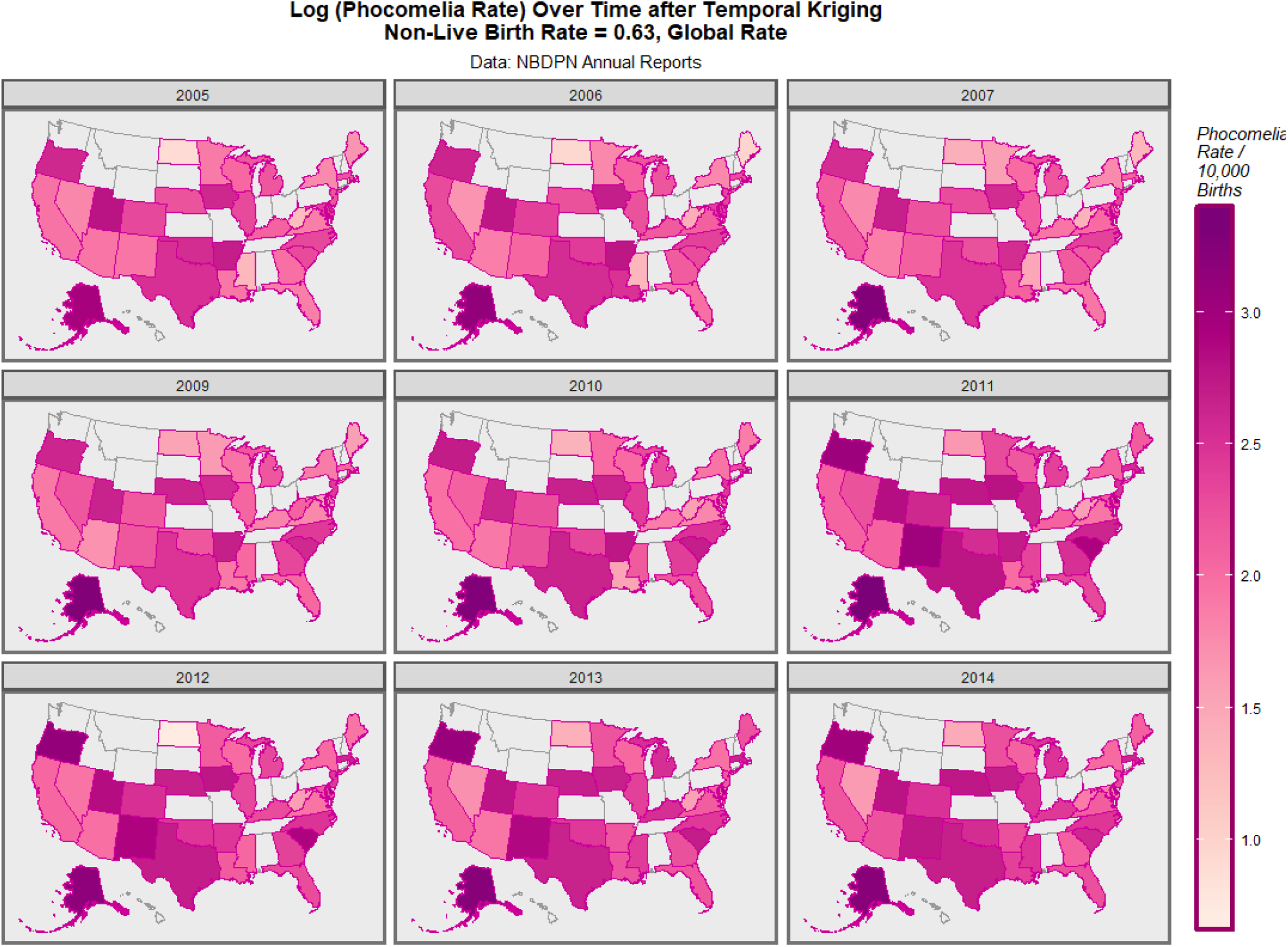
Map-Graphs of LRR Over Time – Adjusted with International Silent Factor – Kriged Data.

**Supplementary Figure 16.:**
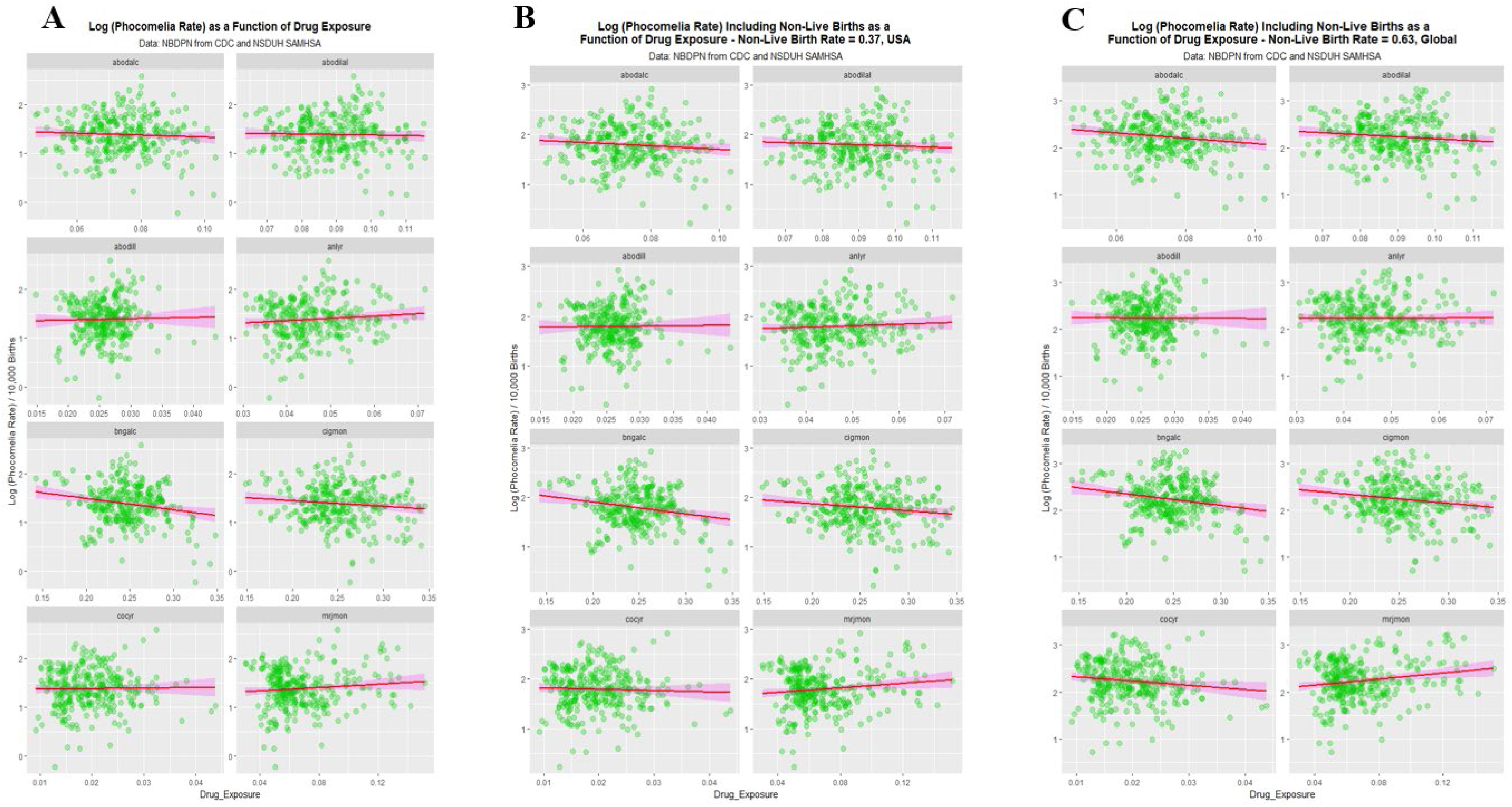
Drug Exposure Plots for Hidden Factors of 0%, 37% and 63%.

**Supplementary Figure 17.:**
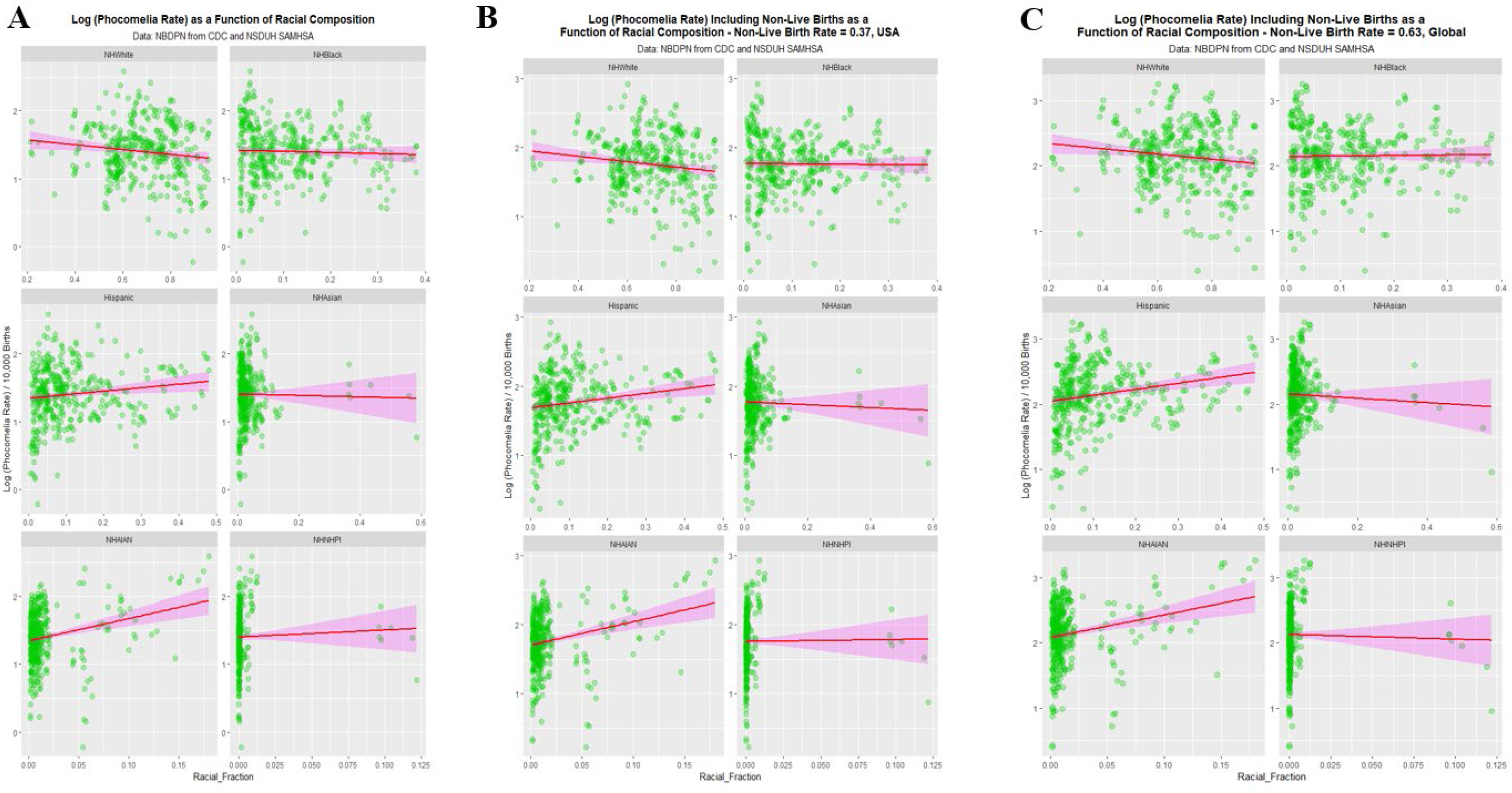
Ethnic Composition Plots for Hidden Factors of 0%, 37% and 63%.

**Supplementary Figure 18.:**
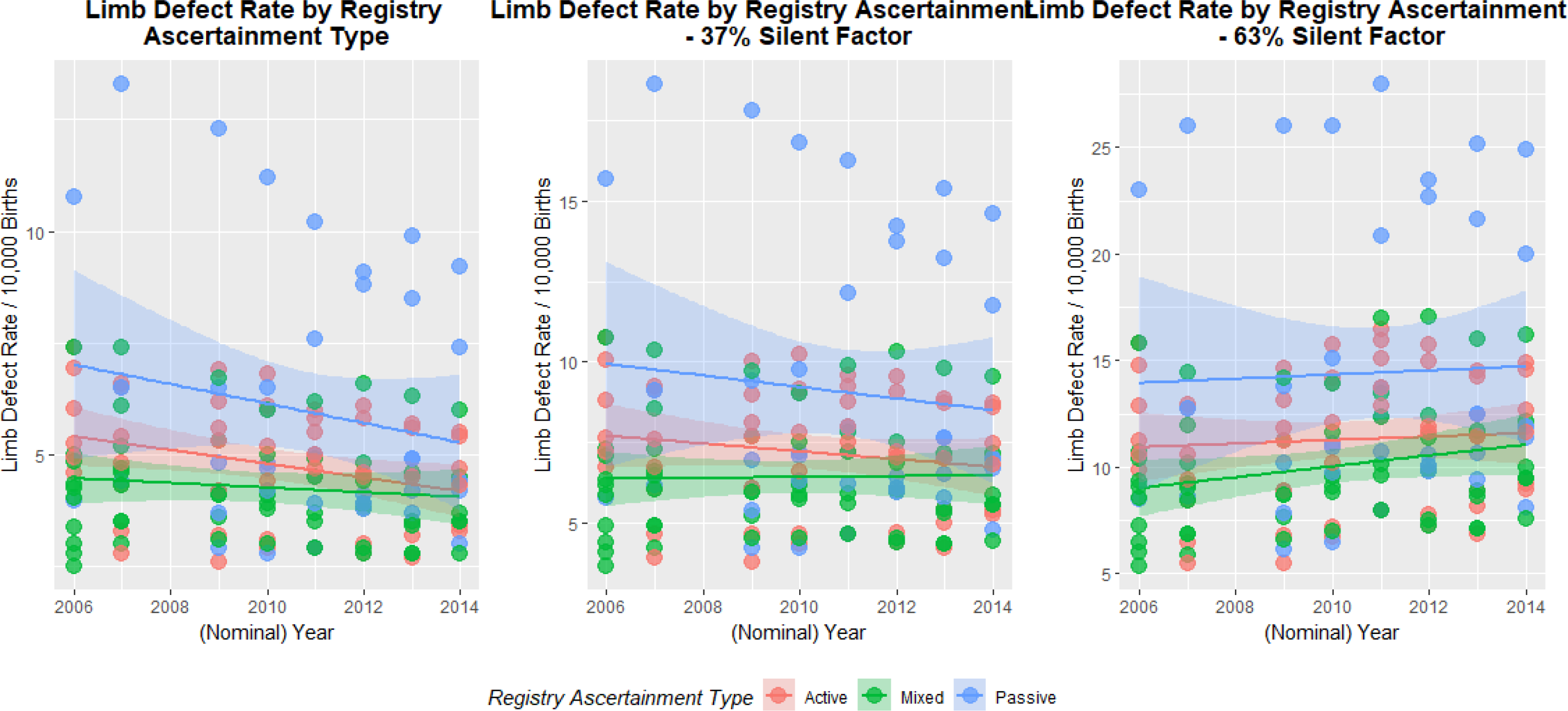
Effects of Registry Ascertainment and Case Finding Practices.

